# Novel Digital Markers of Sleep Dynamics: A Causal Inference Approach Revealing Age and Gender Phenotypes in Obstructive Sleep Apnea

**DOI:** 10.1101/2024.10.23.24315965

**Authors:** Michal Bechny, Akifumi Kishi, Luigi Fiorillo, Julia van der Meer, Markus Schmidt, Claudio Bassetti, Athina Tzovara, Francesca Faraci

## Abstract

Despite evidence that sleep-disorders alter sleep-stage dynamics, clinical practice resists including these parameters in PSG-reports. Leveraging the matrix of sleep-stage transition proportions, we propose (i) a general framework to quantify sleep-dynamics, (ii) several novel markers of their alterations, and (iii) demonstrate our approach using Obstructive Sleep Apnea (OSA), the most prevalent sleep-disorder. Using causal inference techniques, we address confounding in an observational clinical database and estimate markers personalized by age, gender, and OSA-severity. Importantly, our approach adjusts for five categories of sleep-wake-related comorbidities, a factor overlooked in existing research but present in 48.6% of OSA-subjects in our high-quality dataset. Key markers, such as NREM-REM-oscillations and sleep-stage-specific fragmentations, were increased across all OSA-severities and demographic groups. Additionally, we identified distinct gender-phenotypes, suggesting that females may be more vulnerable to awakenings and REM-sleep-disruptions. External validation of the transition markers on the SHHS database confirmed their robustness in detecting sleep-disordered-breathing (average AUROC = 66.4%). With advancements in automated sleep-scoring and wearable devices, our approach holds promise for developing low-cost screening tools for sleep-, neurodegenerative-, and psychiatric-disorders exhibiting altered sleep patterns.

## Introduction

The clinical sleep study (polysomnography, PSG) involves comprehensive overnight monitoring of body biosignals, including encephalogram (EEG), electrocardiogram (ECG), electromyogram (EMG), and others. Medical personnel evaluate the PSG following guidelines of the American Academy of Sleep Medicine (AASM)^1^, focusing on the detection of breathing arrests, movement events, and notably, categorizing stages of sleep. Sleep scoring - conventionally done manually for each 30-second window (epoch) of the biosignals recorded - differentiates between five sleep-wake stages: wakefulness (W), rapid-eye-movement (REM) sleep, and three other non-REM (N1, N2, N3) sleep-states. Such a structured sleep-scoring (hypnogram) forms a basis for the PSG report, providing information on basic markers (e.g., sleep efficiency, % of sleep-stages, REM latency) that relate to sleep quality and may also indicate certain sleep disorders^2–4^.

Sleep and its markers have a complex relationship with individuals’ age and may vary by gender^5^. Several meta-analyses have made considerable efforts to establish normative values of sleep markers in healthy individuals^6,7^. However, the validity of certain estimates might be questionable due to inappropriate statistical evaluations of the individual studies whose results were pooled^8^. For instance, REM latency, as a time-to-event phenomenon subject to censoring, is best quantified using survival techniques rather than mean comparisons. Similarly, the % of sleep-stages, which are interdependent, should be assessed by compositional methods. Proper techniques enabling unbiased estimation are however rarely applied. Quantification of normative ranges and changes in sleep markers in diseased subjects is even more challenging. The observational study design of PSG databases, typically including non-randomized symptomatic subjects, introduces a high degree of confounding^9^. This results in an imbalanced prevalence of individuals with different clinical statuses and distributional shifts in their demographic characteristics. These factors make it difficult to separate the effects of natural ageing from the effects of particular disorders on sleep parameters. The unaddressed confounding, difficulty in assessing data of patients who often suffer from several sleep disorders simultaneously, and the use of not always appropriate statistical approaches are major challenges that increase the risk of biased conclusions even in the analysis of well-established PSG markers.

While differences in sleep-stage dynamics are evident for certain sleep disorders, such as increased sleep fragmentation in Obstructive Sleep Apnea (OSA)^10,11^, or a short REM latency in narcoleptic patients^12^, the clinical PSG report has, so far, included only a limited number of dynamics-related markers. This includes sleep and REM latencies and the absolute counts of sleep-stage transitions or awakenings^1^. While latencies target the first (tens of) minutes of the night, the numbers of transitions/awakenings are proportional to sleep duration and may not sufficiently capture more complex patterns of sleep dynamics that may be specific to individual sleep disorders. Despite the clinical utility of studying sleep dynamics, there is resistance to incorporating its parameters into the PSG report, primarily due to the lack of a uniform methodology that provides a valid and intuitive framework for their evaluation by medical professionals. Recognizing these limitations, significant research has been conducted to comprehensively explore sleep dynamics in various modalities. These studies, which date back to the 1980s, exhibit different levels of heterogeneity in terms of subject demographics, clinical diagnoses, and the methodologies employed^13^. Two main investigative directions have emerged: (i) *focusing on the transitions between sleep stages*, and (ii) *focusing on the duration of sleep stages*. The perspectives of these two seemingly distinct but strongly interrelated areas are discussed in the following two separate paragraphs, highlighting the contribution of the most impactful studies.

Research on sleep-stage transitions has evolved rapidly, beginning with one of the earliest mathematical models by Kemp (1986), who quantified transition intensities in 23 healthy males aged 18-30^14^. Yassouridis (1999) followed by exploring the relationship between transition intensities and plasma cortisol levels in 30 males aged 20-30^15^. Several studies identified associations between transition rates and clinical symptoms. For instance, Burns (2008) observed increased sleep fragmentation and transitions into N3 in 15 females with fibromyalgia syndrome (mean ± standard deviation (SD) age of 42.5 ± 12.9), contrasting with age- and gender-matched controls^16^. Laffan (2010) found a significant association between transition rates and self-reported sleep quality in a large cohort from the Sleep Heart Health Study (SHHS) database, consisting of 5684 participants (47.2% males, all aged over 40)^17^. The existing research extends to specific conditions such as chronic fatigue syndrome, where Kishi (2008) reported abnormal REM transitions in 22 female patients (aged 42 ± 8) in comparison to healthy controls of similar demographics^18^. Further exploring clinical implications, Kim (2009) found differences in sleep-stage dynamics between nights with and without CPAP therapy in 113 OSA subjects (aged 54.0 ± 11.7, 16 females)^19^. Wei (2017) documented increased N2-to-W/N1 transitions in 46 insomnia patients (aged 50.3 ± 13.6, 8 males) compared to age- and gender-matched controls, indicating altered sleep patterns^20^. In addition, Schlemmer (2015) analyzed first- and second-order sleep-stage transitions across 4 groups of subjects (young vs old, healthy vs disorder), highlighting the varied impacts of ageing and pathological conditions^21^. Yet, the disordered subjects represented a pool of various sleep and psychological conditions, and the findings cannot be attributed to a specific diagnosis. Recently, Wachter (2020) utilized MANOVA adjusted for age, gender, and BMI, to evaluate differences in the 25 most common second-order transitions in different severities of OSA compared to healthy subjects, demonstrating associations with demographic and clinical factors^22^. The significant findings primarily related to wake and light-sleep (N1, N2) oscillations, when comparing severe-OSA and healthy. An innovative yet not diagnosis-oriented approach by Yetton (2018) applied a Bayesian network to model transitions as well as stage durations in 3202 - according to exclusion criteria - healthy subjects (mean age of 62.5, 60% males). The prediction-oriented results demonstrated the highest accuracy (62.3%) in the identification of the current stage based on the previous 2 stages, the duration of the last stage, and no consideration of age, gender, or BMI^23^.

Another perspective in understanding sleep dynamics focuses on the quantification of sleep stage durations, providing insights into the temporal characteristics of individual sleep-wake periods. Lo (2002) initiated this research direction by examining sleep-wake dynamics in 20 healthy subjects (aged 23-57, 9 males), revealing different characteristics between sleep and wake periods’ duration and advocating for their modelling using power law distributions^24^. Building on this, Penzel (2003) applied power-law models to quantify sleep-stage durations in both healthy and disordered subjects, identifying reduced duration and hence more fragmented sleep in sleep-apnea subjects^25^ (with no specific demographics details provided). Following that, Norman (2006) exploited survival techniques and revealed decreased sleep continuity when comparing 10 mild and 10 moderate/severe subjects with sleep-disordered-breathing (SDB) against 10 normal subjects^26^. The analysis did not consider subjects’ age, which was significantly higher in disordered subjects. Chervin (2009) compared sleep architecture in 48 children (aged 5-12.9) with sleep-disordered breathing to healthy controls, finding a significant decrease in the duration of N2 and REM^27^. Bianchi (2010) employed multi-exponential fitting to analyze sleep-stage durations across 376 predefined controls (aged 68.2 ± 6.3, 35.6% males), in comparison to 496 mild-OSA (aged 63.8 ± 0.3, 60% males), and 338 severe-OSA (aged 63.7 ± 10.5, 70.7% males) subjects from the SHHS database^28^. They report accelerated decay rates in W, NREM, and REM among OSA subjects, suggesting a larger sleep fragmentation and shorter stage bouts. Notably, despite considerable age and gender differences within its sample (35.6% vs 70.7% males in healthy vs severe-OSA), the study did not adjust for them. Klerman (2013) investigated durations of sleep-wake states in healthy subjects and identified an age-related decline of NREM-sleep continuity^29^. A comparison of sleep-stage duration by Kishi (2020) in sleep bruxism (SB) patients (aged 23.3 ± 1.1, 6 males) and matched controls showed that despite no differences in the prevalence of sleep-stages (except for N1), the SB subjects differed in several parameters describing their dynamics, particularly related to an increased REM fragmentation and hence reduced duration of REM-bouts^30^.

By analysing sleep-stage transitions^14–23^ or by characterizing their duration^24–30^, all of these studies highlight the importance and clinical utility of analysing sleep dynamics across a wide range of disorders. Although most of the studies focus on one of these two aspects, it is important to point out that their nature is functionally linked as the lower transition probability relates to an increased bout duration^31,32^. The existing research works have variously addressed the complexities of confounding and the selection of appropriate statistical models. The majority of studies concurred on the need to control for age and gender or limit the demographic ranges to ensure a homogeneous group of study participants. In existing studies, this is achieved by using stratified analysis with (M)ANOVA (e.g.,^21,22,28^), regression adjustment (e.g.,^17^), or selecting matched individuals (e.g.,^16,20,30^). The simplicity of the first two approaches, typically comparing the effect of exposure (such as OSA) on the outcome (e.g., sleep dynamics) against unexposed healthy controls, is offset by its susceptibility to confounding bias^33^. Analyzing non-randomized observational PSG databases, which typically include older, symptomatic individuals, complicates the separation of confounder effects (of age, gender) from the exposure (disorder). In contrast, while the matching approach helps a lot to reduce the bias^34^, it is generally applied within smaller subject cohorts. This limitation arises from the challenges of finding individuals with matched characteristics within typically imbalanced clinical databases of limited sizes.

Our study introduces a comprehensive framework for quantifying sleep dynamics, demonstrated on OSA but applicable to other (sleep) disorders. OSA, the most prevalent sleep disorder affecting up to 17% of the general adult population^35^, serves as a use-case to showcase the framework’s versatility. Building on existing research and addressing its limitations, our framework—depicted in Figure 1 and explained in-depth in *Methods*—fulfils several key objectives:

**Figure 1.**
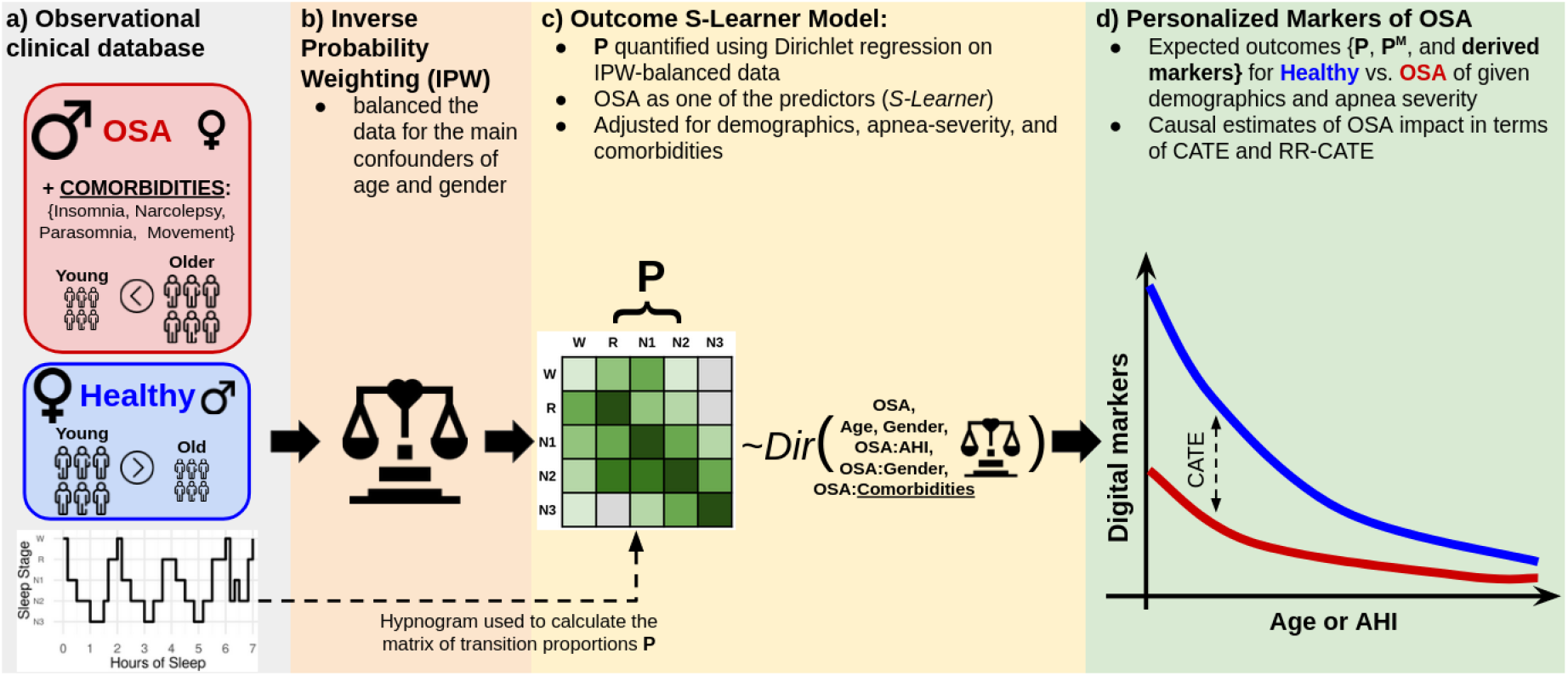
Graphical overview of the implemented approach for quantifying sleep-stage dynamics. Part a): The study utilized observational data, including hypnograms of subjects with a conclusive diagnosis of either Obstructive Sleep Apnea (OSA) or healthy status. The illustration highlights differences in the overall prevalence of OSA (OSA-affected > healthy) concerning gender (male predominance in OSA), age (higher OSA prevalence in older subjects), and comorbidities (not present in healthy subjects). Part b): Inverse Probability Weighting (IPW) is applied to balance the data for the primary confounders of age and gender, having distributional overlap between OSA and healthy subjects. Part c: A sleep fingerprint matrix **P** of sleep-stage transition proportions is modelled using Dirichlet regression within a causal S-Learner framework to capture the effects of OSA, its severity (Apnea-Hypopnea Index, AHI), age, gender, and comorbidities. Part d): The framework quantifies digital markers of OSA (raw **P, P***^M^* as the normalized Markovian **P**, and derived quantities such as sleep fragmentation), personalized for subjects’ demographics, OSA severity, and comorbidities, and presented in terms of Conditional Average Treatment Effect (CATE) and Risk-Ratio CATE (RR-CATE).

- **Data acquisition**, Figure 1a: Leveraging a high-quality, heterogeneous observational clinical database, we identified OSA and healthy subjects (aged 6-91 years) based on clinical gold-standard of *conclusive* diagnosis. Consistent with the literature (e.g.,^17,21,22,28,35^), we identified age and gender as the primary confounders. The subjects’ sleep was summarized through AASM-scored hypnograms, forming the basis for proposing and deriving novel digital markers of sleep and its dynamics. The information about sleep comorbidities was also considered, to adjust our framework for additional possible confounders. This importance of the need for commorbidity-adjustement can be underscored by the fact that 48.6% of OSA subjects in our dataset had at least one sleep-wake comorbidity among their conclusive diagnoses.
- **Balancing confounders**, Figure 1b: To address confounding of age and gender, exhibiting distributional overlap between OSA and healthy subjects, we applied *Inverse Probability Weighting (IPW)* (c.f.,^36–38^) that ensured balanced comparisons between the OSA and healthy groups, regarding the main confounding factors. In short, IPW aims to mathematically re-weight the original dataset, as it was matched regarding the confounders considered (such as age).
- **Sleep dynamics modeling**, Figure 1c: Utilizing hypnograms, we propose a novel *“sleep fingerprint”*, a matrix **P** of sleep-stage transition proportions. As first ones in the field, respecting the interdependencies between individual dimensions of transition proportions **P** (that sum-up to 100%), we quantified them jointly using Dirichlet regression^39^, a method well-suited for the compositional nature of **P**, within a causal S-Learner framework^40^ applied to IPW-balanced data. The idea of causal S-Learner is to extrapolate outcomes for *“conditioned”* (OSA) vs control (healthy) subjects for arbitrary values of predictors. This approach enables the estimation of changes in sleep (dynamics) across different ages, OSA-severities (AHI), and the previously understudied interplay of OSA with gender and sleep-wake-related comorbidities.
- **Digital marker quantification**, Figure 1d: Finally, by exploiting the estimated model (1.c), we quantify not only the estimated effects of OSA on **P** but also derive several novel digital markers. These markers capture the disorder’s impact on sleep, sleep-stage dynamics and also durations, personalized for arbitrary values of predictors (such as age, gender, apnea-severity), and are presented in terms of *Conditional Average Treatment Effect* (CATE) and *Risk-Ratio CATE* (RR-CATE)^41^, standing for absolute and relative comparisons of expected outcomes (such as specific stage-transitions) for OSA and healthy, respectively.

Our framework integrates the two main branches of sleep dynamics research—quantification of sleep-stage transitions and durations—by demonstrating their interconnectedness and enabling their simultaneous quantification. Our study is the first in the field to rigorously account for the interactions between OSA, gender, and a wide range of comorbidities, providing a deeper understanding and less biased estimates of how OSA impacts sleep across various ages, genders, and apnea-severity levels. As demonstrated in our results, the quantified effects and markers of OSA can be leveraged to: (i) *explain*—by establishing normative values for sleep parameters tailored to different demographic profiles and OSA severity; and also (ii) *predict*—by training models capable of identifying OSA subjects based solely on observed demographics and sleep-stage dynamics. The results are publicly accessible through an interactive online app, fostering a broader scientific exploration and discussion.

## Results

The main findings of our study are presented in the four subsections:

- *Modelling of sleep-stage transition matrix*, following Figure 1a-c, presents the estimation of causal S-learner quantifying the matrix of sleep-stage transition proportions **P**, and the impact of predictors, on IPW-balanced data.
- *Personalized digital markers of sleep dynamics and the effects of OSA*, following Figure 1d, introduces principal findings on OSA-markers based on: 1. *raw matrix* **P** exploring the overall prevalence of individual transitions; 2. *derived markers* capturing certain clinical properties by summing up relevant dimensions of **P**; and 3. *derived Markovian matrix* **P**^*M*^ investigating sleep-stage-specific transition mechanisms related to stage durations. The personalization of markers refers to the estimation of the OSA impact for various levels of age, genders, and apnea-severity, helping to understand how OSA alters sleep and its dynamics across different subpopulations.
- *Predictive Performance of* **P** *Markers on External Data* evaluates the utility of each of the 25 possible sleep-stage transition proportions in identifying subjects with moderate sleep-disordered breathing (AHI > 15). A logistic regression model was trained on the study dataset (BSDB) and applied to the large open-access dataset (SHHS), using only age, gender, and the specific transition proportion as predictors.
- The last part introduces our app, which allows interactive exploration of results beyond the scope of the ones presented within this paper (e.g., interactions of OSA with arbitrary comorbidities, evaluation of extreme OSA with AHI > 50, etc).

### Modelling of sleep-stage transition matrix

#### Propensity score model and IPW balancing

To balance the Berner Sleep Data Base (BSDB) study dataset for the main confounders of gender and age, we used the Inverse Probability Weighting (IPW) strategy, c.f., Figure 1a-b. Propensity scores introduced in Eq. 24 were used to calculate weights according to Eq. 25. The estimates of propensity scores were based on the logistic regression model from Eq. 29. The choice of gender and age as the inputs for the IPW was driven by the evidence of existing studies that control for them^17,22^and clinical evidence that OSA is more prevalent in males and at older ages^35^. In the BSDB exploited, both OSA and healthy subjects can be observed across the entire range of age and genders, thus satisfying the assumption of overlap and positivity^37^. After re-weighting the dataset, the characteristics of age and gender were balanced, which was evidenced by a t-test based on IPW-reweighted means and standard deviations that failed to reject (p-val > 0.05) the null hypothesis of equality of variable means between the OSA and healthy subjects. The weights were subsequently used within the outcome model, enforcing the balanced impact of age and gender, across OSA and healthy subjects.

#### Outcome model

The proportions of the 25 possible sleep-stage transitions in **P** were modeled using Dirichlet regression (c.f., Figure 1c) applied to IPW-balanced data. The model specification followed Eq. 30, and the inclusion of the OSA indicator as one of its predictors exploited the causal S-learner framework, enabling a straightforward quantification of (age, gender, apnea-severity)-heterogeneous OSA-effects in terms of Conditional Average Treatment Effect (CATE) and Risk-Ratio CATE (RR-CATE) (c.f., Eq. 27-28). Simplistically, the CATE and RR-CATE refer to absolute and relative differences between conditioned (i.e., OSA-affected) and control (i.e., healthy) subjects, respectively. The model estimation followed the implementation of Dirichlet regression in R^39^. To assess uncertainty, both in the model coefficients and derived effects, the nonparametric bootstrap with 200 repetitions was used to calculate 95% confidence intervals (CI) based on 2.5% and 97.5% bootstrapped quantiles.

A summary of estimated regression coefficients together with CI for each predictor and transition proportion is provided in Supplementary Table 1. The estimates indicate a significant influence of both demographics (age and gender), OSA, and its severity (AHI) on sleep-stage dynamics, as at least one of them had a significant impact on each of the transition proportions. The significant interactions of OSA with gender point to the presence of possible gender-specific OSA phenotypes. The adjustment for comorbidities appears to be essential as the comorbidity indicators influenced most of the transitions.

**Table 1.** AUROC with 95% CI for predicting moderate sleep-disordered breathing (AHI > 15) using individual sleep-stage transition proportions. Results are shown for SHHS1 (Sleep Heart Health Study baseline) and SHHS2 (follow-up), of N = number of subjects after exclusion criteria. Mean ± SD (standard deviation) summarizes performance across all transitions. Asterisk (*) denotes significant predictive power with AUROC (> 50%).

| Transition (Predictor) | SHHS1 (N = 5,734) | SHHS2 (N = 2,621) |
| --- | --- | --- |
| W → W | 67.84* (66.47, 69.21) | 66.51* (64.46, 68.55) |
| W → N1 | 68.71* (67.36, 70.07) | 67.97* (65.95, 69.99) |
| W → N2 | 67.04* (65.66, 68.42) | 66.03* (63.98, 68.09) |
| W → N3 | 66.18* (64.78, 67.57) | 65.62* (63.55, 67.69) |
| W → R | 68.42* (67.06, 69.78) | 68.47* (66.46, 70.48) |
| N1 → W | 69.28* (67.93, 70.62) | 68.39* (66.38, 70.39) |
| N1 → N1 | 66.8* (65.42, 68.18) | 65.57* (63.51, 67.63) |
| N1 → N2 | 66.74* (65.35, 68.12) | 66.27* (64.22, 68.33) |
| N1 → N3 | 67.2* (65.83, 68.58) | 66.23* (64.18, 68.28) |
| N1 → R | 67.54* (66.17, 68.91) | 66.86* (64.82, 68.9) |
| N2 → W | 65.18* (63.78, 66.58) | 63.79* (61.7, 65.89) |
| N2 → N1 | 65.73* (64.34, 67.13) | 65.04* (62.97, 67.11) |
| N2 → N2 | 59.82* (58.37, 61.28) | 59.89* (57.74, 62.05) |
| N2 → N3 | 67.43* (66.06, 68.8) | 66.46* (64.41, 68.51) |
| N2 → R | 66.06* (64.67, 67.45) | 65.16* (63.09, 67.23) |
| N3 → W | 65.82* (64.42, 67.23) | 65.12* (63.04, 67.2) |
| N3 → N1 | 67.19* (65.82, 68.57) | 66.27* (64.22, 68.32) |
| N3 → N2 | 67.45* (66.07, 68.82) | 66.49* (64.44, 68.54) |
| N3 → N3 | 67.89* (66.52, 69.26) | 67.61* (65.58, 69.64) |
| N3 → R | 66.94* (65.56, 68.32) | 65.57* (63.51, 67.64) |
| R → W | 64.87* (63.47, 66.28) | 62.9* (60.79, 65.01) |
| R → N1 | 67.45* (66.08, 68.82) | 67.15* (65.11, 69.18) |
| R → N2 | 66.41* (65.02, 67.8) | 66.01* (63.95, 68.06) |
| R → N3 | 67.03* (65.65, 68.41) | 66.19* (64.14, 68.24) |
| R → R | 68.18* (66.82, 69.54) | 67.88* (65.86, 69.89) |
| Mean ± SD | 66.77 ± 1.79 | 65.98 ± 1.82 |

Given the complex relationship of the marginal effect on the outcome (i.e., transition %’s) with individual coefficients and the actual predictors’ value (c.f., Eq. 18), we detail results in the intuitive scales of expected percentages, differences (CATE, Eq. 27), and risk-ratios (RR-CATE, Eq. 28), below.

### Personalized digital markers of sleep dynamics and the effects of OSA

The estimated outcome model enables various scenarios of comparisons of OSA vs healthy, including the raw matrix **P**, derived markers (e.g., % of sleep-stages), and Markovian transition matrix **P**^*M*^, c.f., Figure 1d. All this, for arbitrary values of predictors, provides a wide range of results that can inspire new investigative directions. Since all of our results refer to (possibly derived) transition probabilities (%), we present them in *RR-CATE (CATE)%* format, indicating the amount of *relative (absolute)%* changes, respectively.

Utilizing our model (Eq. 30) can extrapolate OSA-effects for arbitrary values of predictors, we showcased the results for three scenarios according to OSA severity, **O1: mild (AHI = 5), O2: moderate (AHI = 15)**, and **O3: severe (AHI = 30)**; three ages: **A1: young (30 years), A2: middle-aged (50 years), A3: older (70 years)**; and for **females (F)** and **males (M), *without comorbidities***. When selecting the most prominent effect in a group, we choose the one according to RR-CATE.

#### *Matrix* P *of sleep-stage transition proportions*

The heatmap in Figure 2 shows whether individual transition proportions in **P** (Eq. 1) were significantly altered due to specific OSA conditions across different ages and genders. All these aggregated findings are based on detailed results depicted as supplementary heatmap figures supplemented with respective estimates and CI. Supplementary Figures 1 and 4 depict expected **P** for different ages and OSA-severities for F and M, respectively. Based on that, Supplementary Figures 2 and 5 present CATE comparisons between different levels of OSA and healthy individuals of the same demographics, and Supplementary Figures 3 and 6 depict the respective RR-CATE.

**Figure 2.**
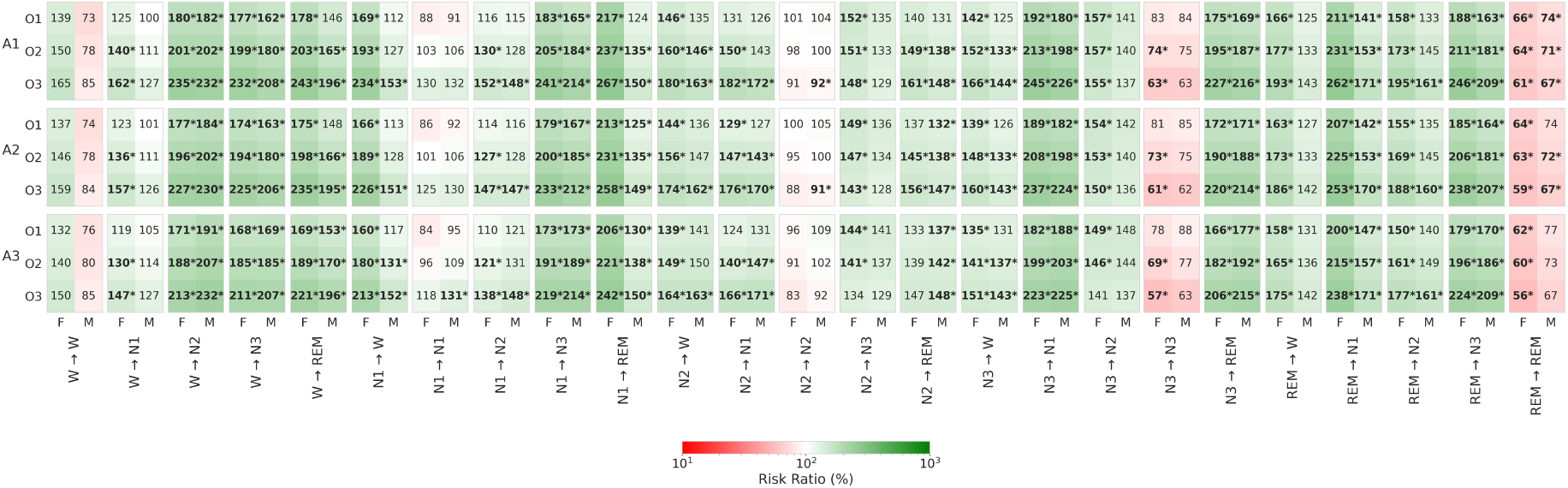
Heatmap of Risk-Ratio Conditional-Average-Treatment-Effects (RR-CATE) of OSA (compared to a matched healthy subject) on individual dimensions of sleep-fingerprint matrix **P** of sleep-stage transition proportions, per gender (F, M), age (A1, A2, A3), and OSA-severity (O1, O2, O3). Decreased (i.e., RR < 100%) and increased (i.e., RR > 100%) risk-ratios are depicted with red and green shaded backgrounds, respectively. Significant effects are in bold and highlighted with a star (*).

**Figure 3.**
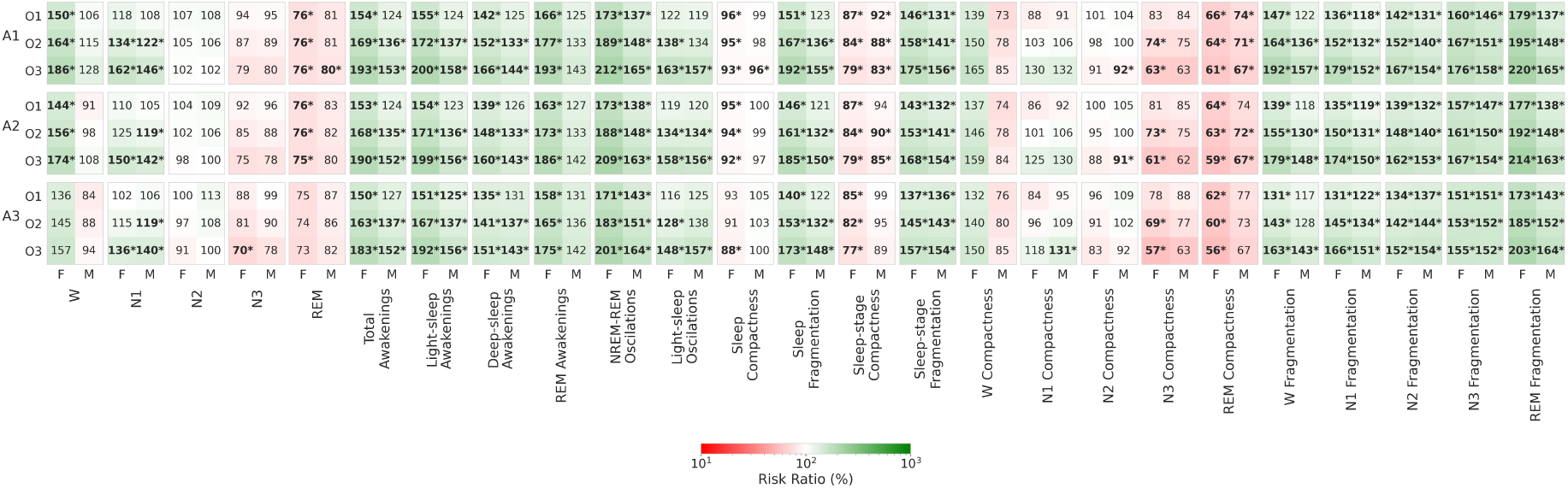
Heatmap of Risk-Ratio Conditional-Average-Treatment-Effects (RR-CATE) of OSA (compared to a matched healthy subject) on PSG-markers derived from matrix **P** of sleep-stage transition proportions, per gender (F, M), age (A1, A2, A3), and OSA-severity (O1, O2, O3). Decreased (i.e., RR < 100%) and increased (i.e., RR > 100%) risk-ratios are depicted with red and green shaded backgrounds, respectively. Significant effects are in bold and highlighted with a star (*).

**Figure 4.**
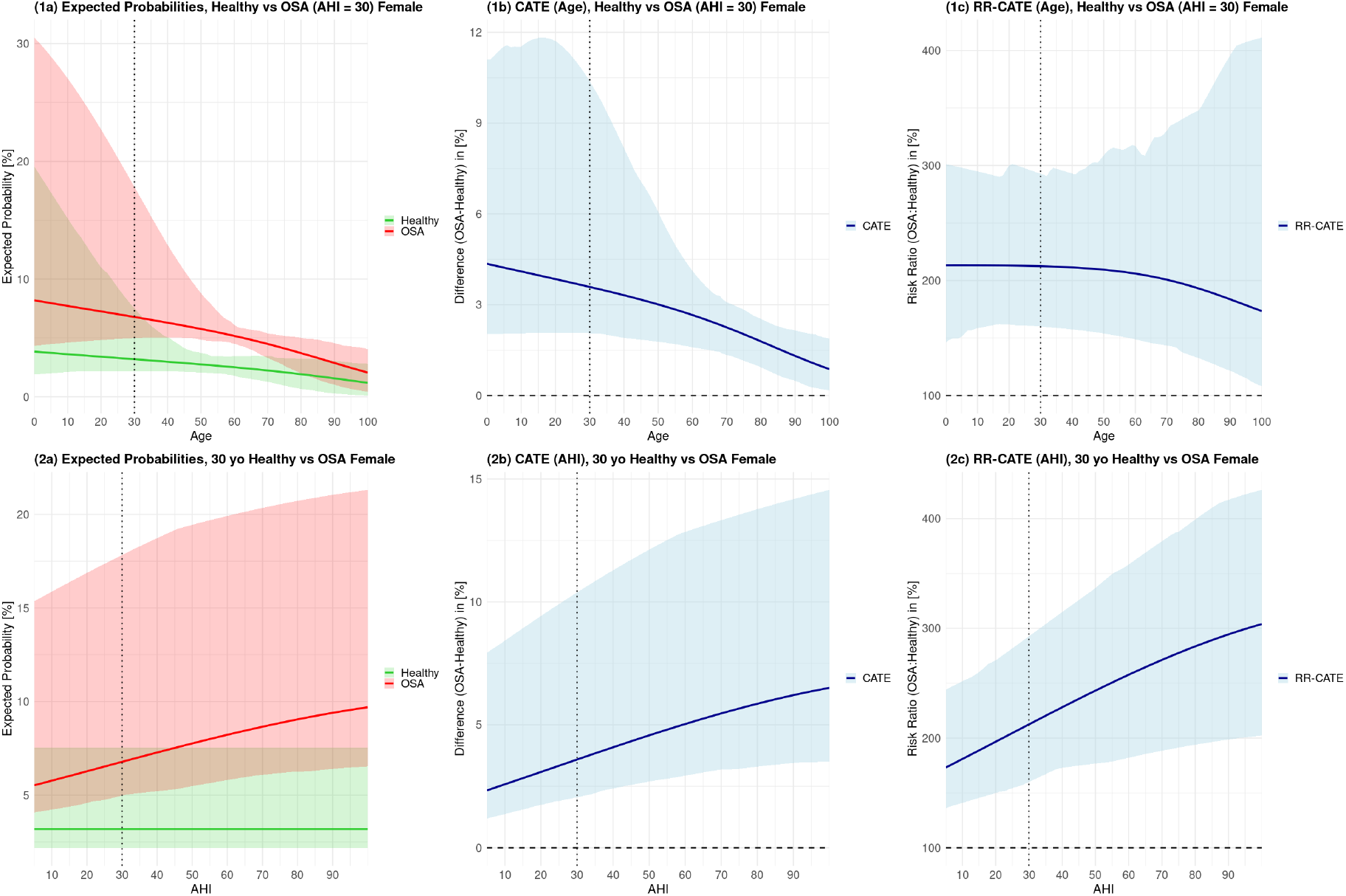
Effects of age and OSA-severities on NREM-REM oscillations, *P*(NREM ? REM), in females. The left plots (1a, 2a) depict expected probabilities for varying age with fixed AHI = 30, and for varying AHI with fixed age = 30. Based on that, the central (1b, 2b) and right (1c, 2c) plots depict age- and AHI-related CATE and RR-CATE.

**Figure 5.**
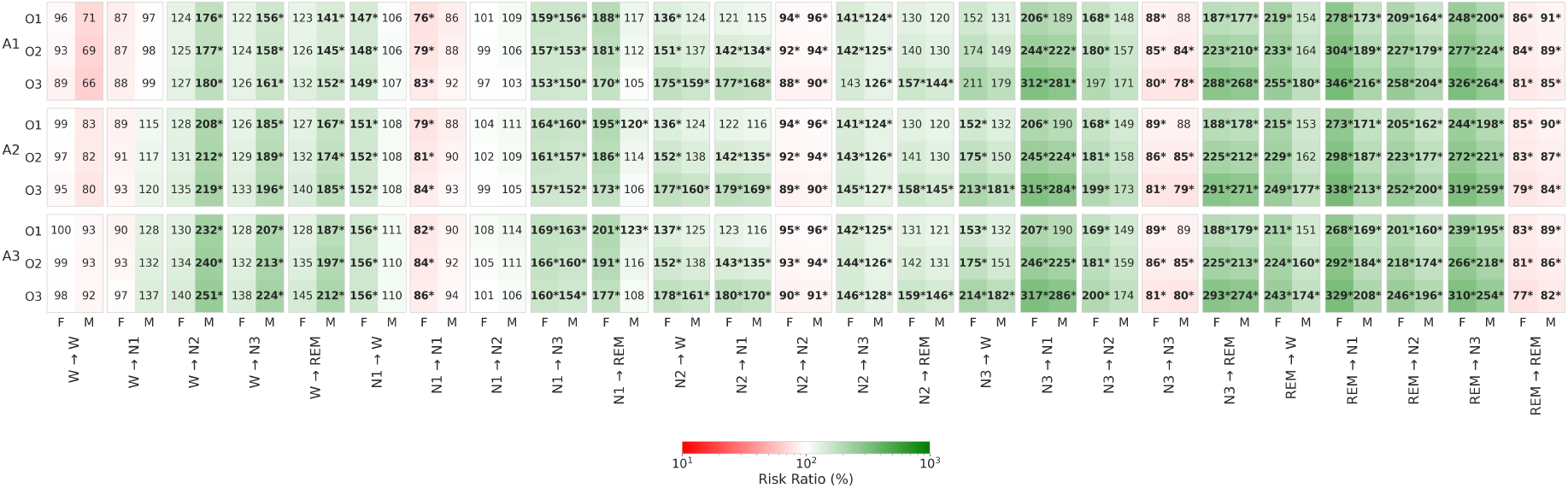
Heatmap of Risk-Ratio Conditional-Average-Treatment-Effects (RR-CATE) of OSA (compared to a matched healthy subject) on individual dimensions of row-normalized Markovian transition matrix **P**^*M*^, per gender (F, M), age (A1, A2, A3), and OSA-severity (O1, O2, O3). Decreased (i.e., RR < 100%) and increased (i.e., RR > 100%) risk-ratios are depicted with red and green shaded backgrounds, respectively. Significant effects are in bold and highlighted with a star (*).

Notably, except for N2 → N3 and N3 → N2 of A3-F, each significant effect identified for O1 or O2 of both genders was followed with significant effect in the corresponding more severe OSA group. This follows the intuition, that the sleep-stage dynamics and hence also **P** change gradually with increasing prevalence of apnea events (i.e., AHI). The exemption of older F is justified by a significantly lower % of N3, 70.04 (-5.6)% in A3-O3 (c.f., supplementary Table 4).

As the entire **P** sums up to 100%, each decrease in a certain proportion is compensated with an increase in one or more other ones. For F, a major decrease is observed in REM → REM, with RR-CATE of about 60% across all ages and OSA severities, and the most prominent drop, 55.55 (-4.85)%, in older. This suggests significant REM sleep instability, which could impact cognitive health^42^. The O2- and O3-F also show significantly decreased N3 → N3, as low as 57.08 (-6.97)% in A3, indicating disrupted deep-sleep continuity, which may affect physical restoration and memory consolidation^43^. For A1-M, REM → REM decreased for all OSA severities, down to 67.5 (-6.19)%, and for A2-(O2,O3), 66.93 (-4.73)%, with the largest declines always in O3. The decreases in all A3-M-OSA groups were not significant, likely due to a larger variance in estimates caused by the limited number of healthy older M in the data. Contrary to F, a decrease in N3 → N3 was not significant in M, but a significant decrease in N2 → N2 was noted for (A1, A2) O3-OSA, as low as 91.09 (-3.22)%.

For both genders of all ages and OSA severities, several significantly increased transition proportions were identified, distinguishing them from healthy subjects. The most pronounced effects were found in A1-O3-F. The increased W → (N2, N3) transitions, up to 234.6 (0.4)%, indicate more frequent arousals attributable to apneic events and subsequent attempts to quickly regain restorative sleep. Increased transitions from N1→N3, up to 241.0 (0.4)%, suggest a compensatory mechanism where the body attempts to achieve the restorative effects of deep sleep, bypassing intermediate stages due to frequent sleep disruptions. The increase in N3→(N1, REM) transitions, up to 245.5 (0.3)%, indicates frequent deep sleep disruptions, causing a regression to lighter sleep or irregular shifts to REM sleep. Lastly, elevated REM→(N1, N3) transitions, up to 261.6 (0.6)%, reflect REM stage instability, with more frequent abrupt changes in sleep depth.

Interestingly, all OSA-F showed a significant increase in awakenings from all sleep stages, (N1, N2, N3, REM) →W. For M, there was no increase in REM → W in any OSA group, and increases in (N1, N2, N3) → W were observed only for O2 and O3. This suggests that in comparison to M, the OSA-F may experience more fragmented sleep due to frequent awakenings from all stages, potentially leading to greater daytime sleepiness.

#### *PSG markers derived from* P

The heatmap in Figure 3 aggregates the OSA-effects identified for different PSG markers (c.f., Eq. 2-13) derived from **P**. Detailed results concerning expected probabilities (%) of their occurrence following Eq. 18-19, CATE, and RR-CATE for individual age and OSA categories are provided in Supplementary Tables 2-4 for F, and Tables 5-7 for M, respectively.

**Table 2.** Comparison of demographics, sleep metrics, and prevalence of sleep comorbidities among healthy and (mild, moderate, severe) OSA subjects in the BSDB dataset. Variables denoted with ^*^ are binary, summarized as count (percentage), N (%), and significantly different pairs are listed, following a significant chi-squared independence test and pairwise posthoc proportions test. Healthy subjects were excluded from comorbidities comparisons as they had no comorbidities. Variables denoted with ^†^ are continuous, summarized as mean (standard deviation), *µ*(*σ*), and significant pairs are listed following a significant Kruskal-Wallis test and pairwise Wilcoxon posthoc test. All posthoc pairwise comparisons were performed with Bonferonni corrections at the significance level of 0.05.

|  | H: Healthy | O1: Mild OSA | O2: Moderate OSA | O3: Severe OSA | Significant Pairs |
| --- | --- | --- | --- | --- | --- |
| **DEMOGRAPHICS:** |  |  |  |  |  |
| Subjects [count] | 62 | 238 | 164 | 158 |  |
| *Males | 25 (40.3) | 166 (69.7) | 117 (71.3) | 127 (80.4) | O1H, O2H, O3H |
| *Females | 37 (59.7) | 72 (30.3) | 47 (28.7) | 31 (19.6) | O1H, O2H, O3H |
| †Age | 34.9 (18) | 50.6 (14.9) | 53.8 (14.7) | 58 (11.9) | O1H, O2H, O3H, O3O1 |
| **SLEEP METRICS:** |  |  |  |  |  |
| †TST [minutes] | 370.3 (62.9) | 345.5 (74.3) | 344.1 (76.4) | 321.4 (64.2) | O2H, O3H, O3O1, O3O2 |
| †Efficiency [%] | 88.4 (6.6) | 83.4 (11.7) | 81.3 (12.2) | 78.9 (12.5) | O2H, O3H, O3O1 |
| †Sleep latency [minutes] | 16.5 (15.4) | 12.9 (19) | 16.6 (24) | 15.7 (22) | O1H |
| †REM latency [minutes] | 113.4 (50.7) | 138.9 (80.1) | 124.4 (72.6) | 148.6 (86.3) | - |
| †Hourly transitions [$\frac{N}{\text{hour}}$] | 16.2 (4.9) | 20.6 (5.6) | 22.2 (6.1) | 25.9 (7.9) | O1H, O2H, O3H, O2O1, O3O1, O3O2 |
| †Hourly awakenings [$\frac{N}{\text{hour}}$] | 2.4 (1.2) | 3.2 (1.7) | 3.3 (1.7) | 4 (2.8) | O1H, O2H, O3H, O3O1 |
| †W [%] | 11.6 (6.6) | 16.6 (11.7) | 18.7 (12.2) | 21.1 (12.5) | O1H, O2H, O3H, O3O1 |
| †N1 [%] | 9.4 (6.3) | 13.6 (7.6) | 16.3 (9.1) | 24.1 (13.3) | O1H, O2H, O3H, O2O1, O3O1, O3O2 |
| †N2 [%] | 40.6 (10.3) | 39.9 (10.3) | 36.1 (10.7) | 34.1 (13.2) | O2H, O3H, O2O1, O3O1 |
| †N3 [%] | 21.7 (8.7) | 16.9 (9.7) | 16.4 (11.1) | 10.2 (7.9) | O1H, O2H, O3H, O3O1, O3O2 |
| †REM [%] | 16.7 (7.1) | 13.1 (6.7) | 12.5 (6.4) | 10.4 (6.5) | O1H, O2H, O3H, O3O1, O3O2 |
| **SLEEP COMORBIDITIES:** |  |  |  |  |  |
| *No comorbidity | 62 (100) | 94 (39.5) | 85 (51.8) | 109 (69) | O3O1, O3O2 |
| *Single comorbidity | 0 (0) | 52 (21.8) | 37 (22.6) | 30 (19) | - |
| *Multiple comorbidities | 0 (0) | 92 (38.7) | 42 (25.6) | 19 (12) | O2O1, O3O1, O3O2 |
| *Insomnias | 0 (0) | 44 (18.5) | 22 (13.4) | 15 (9.5) | - |
| *Narcolepsy type 1 | 0 (0) | 11 (4.6) | 7 (4.3) | 4 (2.5) | - |
| *Other hypersomnias | 0 (0) | 88 (37) | 37 (22.6) | 15 (9.5) | O2O1, O3O1, O3O2 |
| *Parasomnias | 0 (0) | 28 (11.8) | 25 (15.2) | 22 (13.9) | - |
| *Movement-related | 0 (0) | 23 (9.7) | 13 (7.9) | 12 (7.6) | - |
| *Circadian-rhythm-related | 0 (0) | 2 (0.8) | 3 (1.8) | 1 (0.6) | - |

Regarding the percentagess of individual sleep-stages, the main effect of OSA shared between both genders of all ages is the increase in N1 in O3, with the largest increase of 161.94 (5.53)% in A1-F. The increase affected also all O2-M, up to 122.36 (2.57)% in A1, and A1-O2-F, 134.41 (3.07)%. F seem to have more affected sleep macro-architecture by OSA than M, as for all OSA-severities of (A1, A2)-F an additional increase in W%, up to 185.63% (3.57%) in A1-O3, suggesting a reduced sleep-efficiency, and decreased REM%, as low as 74.9 (-4.25)% in A2-O3, was identified. Except for reduced REM% in A1-O3-M, 79.54 (-4.46)%, these changes were identified only in F.

In addition to increased N3- and REM-awakening from Eq. 5-6 already discussed above, increased aggregates of total-awakenings (Eq. 3), up to 192.55 (2.89)%, and of light-sleep-awakenings (Eq. 4), up to 200.35 (2.01)%, were observed in all age and OSA categories with exception of O1-M, with largest effects in A1-O3-F.

A particularly sensitive marker of OSA for all severities appear to be NREM-and-REM oscillations (Eq. 7), which were identified as significantly increased across all groups, peaking at 212.48 (3.59)% in A1-O3-F. This marker is elaborated in detail in Figure 4 showcasing the expected outcome for F. The upper plots (1a-c) depict the expected probability (%), CATE, and RR-CATE and corresponding CIs for varying age (and fixed AHI), whereas the bottom plots (2a-c) for varying AHI (and fixed age). One can observe, that the effect of OSA remains significant over the entire range of both, age and AHI. The magnitude of the difference tends to decrease with age (c.f., 1b-c), from CATE of about 4.5% in children to 1.5% in older age, likely due to generally shorter sleep with decreasing REM% and lower number of sleep cycles. The effect’s size increases rapidly with AHI (c.f., 2b-c), which typically increases with age. The outcomes for M are illustrated in supplementary Figure 13.

Another two highly sensitive derived markers of OSA include sleep- and sleep-stage-fragmentation from Eq. 10 and 12, referring to probabilities of transitions between wakefulness and sleep, and switching from one non-W stage to the other, respectively. The effect of the sleep-fragmentation was significant across all groups except O1-M and peaked at 192.33 (5.66)% for A1-O3-F. The sleep-stage-fragmentation was increased in all groups, peaking at 174.94 (10.42)% in A1-O3-F. The sleep-stage-fragmentation marker is in-depth elaborated in supplementary Figures 14 and 15, for F and M, respectively.

The increased fragmentation is reflected in decreased sleep- and sleep-stage-compactness from Eq. 9 and 11, referring to staying in not-interrupted sleep and sleep-stage, respectively. Reduced sleep-compactness, down to 88.42 (-9.65)% in A3-O3-F, seems specific to F, suggesting their more frequent apnea-related arousals than M. The sleep-stage-compactness was reduced in all categories of F, down to 76.77 (-16.54)% in A3-O3. This decrease, however, was not present for A3-M and A2-O1-M.

The reduced stage-specific-compactness metrics (e.g., REM → REM) were already elaborated in the section on **P**-specific transition %’s. Yet, the stage-specific-fragmentation markers (Eq. 13) show significant alterations due to OSA across almost all demographic groups. The only gender-specific difference can be observed in wake-fragmentation, which is increased in all cases of F (likely due to more frequent awakenings experienced), up to 192.1 (2.77)% in A1-O3, but not for O1- and A3-O2-M. The fragmentation related to non-REM (N1, N2, N3) stages increased in all OSA and demographics groups, ranging from 118.29 (1.18)% in N1-fragmentation in A1-O1-M to 178.61% (4.05%) in A1-O3-F. The most pronounced effects were visible in REM-fragmentation, up to 219.51 (2.32)% in A1-O3-F, referring to more than twice as many transitions leaving REM sleep.

#### *Markovian transition matrix* P^M^ *derived from* P

Finally, we present the main findings based on **P**^*M*^, derived from **P** through row normalization as shown in Eq. 14. While **P** quantifies the overall probabilities (%) of the 25 sleep-stage transitions, **P**^*M*^ conditions on the presence of a specific stage, summing to 100% per row. Therefore, whereas **P** evaluates overall chances of observing specific transitions in the hypnogram during the night (e.g., 36.4% of N2 → N2 in healthy A1-F), the **P**^*M*^ evaluates the distribution of the next sleep stage given the current stage (e.g., 84.3% to stay in N2 in healthy A1-F), offering another perspective on the underlying mechanisms of sleep dynamics. The heatmap in Figure 5 depicts how individual transitions of **P**^*M*^ (Eq. 14) altered due to specific OSA conditions across different ages and genders. Detailed results on expected transition probabilities of **P**^*M*^, CATE, and RR-CATE for comparisons of OSA vs healthy are provided in heatmap Supplementary Figures 7-9 and 10-12 for F and M, respectively.

##### W-transitions

Despite increased occurrences of **P**-transitions from W in F, the respective **P**^*M*^ -dynamic was not significantly altered, indicating that the mechanism of the W-transitions remains similar to healthy subjects, but those transitions tend to occur more often. This suggests that for OSA-F, the overall increased W% is the main trigger of the W-related transitions in **P**. Conversely, M exhibit increased W → (N2, N3, REM) transitions, up to 250.5 (1.3)% in A3-O3 for W → N2, across all ages and OSA severities, suggesting an increased sleep pressure due to its disruption induced by apneic events.

##### N1-transitions

Both genders showed increased N1 → N3, up to 169.4 (0.9)% in A3-O1-F. Only F experience increased N1 →W, up to 156.5 (4.7)% in A3-O1, and decreased N1→ N1, as low as 76.5 (-10.3)% in A1-O1. Increased N1 → REM transitions were present in all F, up to 201.1 (3.4)% in A3-O1, but only in some of the O1-M, up to 122.7 (1.1)% in A3.

##### N2-transitions

All groups have decreased N2 → N2, down to 88.4 (-9.8)% in A1-O3-F, and, except for A1-O3-F, significantly increased N2 → N3 transitions, up to 145.6 (2.2)% in A3-O3-F. All F groups have increased N2 → W transitions, up to 177.6 (1.6)% in A3-O3-F, which is present also in all O3-M. N2 → N1 increased for all O2 and O3 groups, up to 179.8 (4.2)% in A3-O3-F, and N2 → R increased for all O3.

##### N3-transitions

Across all groups, the N3 dynamic had significantly increased transitions into REM, peaking up to 293.1 (2.1)% in A3-O3-F, pointing to almost three times higher occurrence of these atypical transitions in OSA. Additionally, decreased N3 → N3, as low as 77.9 (-18.0)% in A1-O3-M, and increased N3 → N1, up to 316.8% (2.5%) in A3-O3-F, were noted for all except O1-M. Transitions N3 → W increased in all (A2, A3)-F, up to 214.1% (2.6%) in A3, and only in O3-M of the same demographics.

##### REM-transitions

The most prominent effects of OSA are visible in changed REM dynamics. The decrease in REM → REM in both genders of all ages, down to 77.1 (-20.4)% in A3-O3-F, is compensated by increased transitions into all NREM-stages, up to 345.8 (5.9)% in REM → N1 for A1-O3-F. The increased REM → W is specific for all F, up to 254.8 (5.3)% in A1-O3-F. For M, these transitions are decreased partially for all O3 and A3-O2, up to 180.0% (2.8%) in A3-O3.

##### Stage-survival

Finally, following Eq. 15, the diagonal elements of **P**^*M*^ (i.e., probabilities of W → W, N1 → N1, etc.) simplistically approximate the average expected duration of individual sleep stages, bridging transition dynamics with investigations modelling the sleep-bout durations. Here, naturally, significantly decreased probabilities of staying in a given stage introduced above are equivalent to significantly decreased stage durations.

### Predictive Performance of P Markers on External Data

The results of the previous sections focused on quantifying the effects of OSA, specifically explaining how OSA impacts sleep dynamics and its markers. To illustrate the informativeness of these markers, we developed a simple logistic regression model for each of the 25 transition proportions in **P**. The binary outcome variable was defined as moderate sleep-disordered breathing, indicated by AHI > 15, and the predictions were based on three predictors: age, gender, and the percentage of a specific transition. The inclusion of age and gender was motivated by the observed heterogeneity of OSA effects with respect to these factors. Each of these logistic regression models, trained on the study dataset (BSDB), was used to make predictions on the SHHS1 and SHHS2 subsets of SHHS, which contain observational data on subjects who underwent baseline PSG (SHHS1) and follow-up PSG several years later (SHHS2, N = 2,621).

The results in Table 1 indicate that each transition proportion included in the simple predictive model demonstrated significant predictive power, as all AUROCs and their confidence intervals were much greater than 50%. The average AUROC performance across proportions was 66.77% for SHHS1 and 65.98% for SHHS2, with standard deviations of 1.79% and 1.82%, respectively, highlighting practical equivalence in generalizations between SHHS1 and SHHS2. This robustness is particularly notable given that (i) we used a simple logistic regression model that assumes monotonic effects of individual predictors and no interactions between them, (ii) we predicted moderate sleep-disordered-breathing (AHI > 15) using only age, gender, and the percentage of a single transition, (iii) the models were trained on a relatively small dataset of 622 patients, and (iv) the generalization was performed from the clinical population of BSDB to the broader public represented by SHHS. All AUROCs are very similar, which can be explained by the fact that all transitions in **P** are interdependent and numerically share related information.

### Interactive R Shiny app

The above-presented results focused on three categories age (30, 50, 70 years), OSA severity (mild, moderate, severe), and both genders, considering a case without sleep-comorbidities. For a deeper exploration of our findings, the volume of which is beyond the scope of this paper, we created a freely accessible app (https://mystatsapps.shinyapps.io/Causal_Sleep_Dynamics/) that interactively displays results for arbitrary values of predictors. As an input, the user specifies the transition(s) of interest by clicking out some of the 25 (5 × 5) dimensions, age, OSA severity (AHI), and the presence of comorbidities (as indicated in Eq 30). Additionally, the user chooses whether CATE and RR-CATE should be displayed for age or AHI (= `CATE-variable`).

As an output, the app displays a total of six panels. The most important one, `Effects of OSA`, displays expected probabilities (%) of selected transitions for healthy vs OSA together with corresponding CATE and RR-CATE. All these outputs are supplemented by 95% CI and are depicted for selected age (range 0-100 years) or AHI (range 5-100), and both genders.

The `Percentual Transition Matrix` and `Markovian Transition Matrix` tabs show the expected matrix of sleep-stage transitions **P** and the derived row-normalized **P**^*M*^ for healthy and OSA subjects of both genders and specified characteristics. In addition, each tab shows matrices of CATE and RR-CATE depicted as heatmaps supplemented with 95% CI.

The `Dirichlet Regression Coefficients` tab summarizes regression coefficients as presented in Supplementary Table 1. The dimensions of specified transitions of interest from the input are highlighted.

The `Marginal Effects of All Predictors` tab approximate the Eq. 18 by calculating the difference in the outcome by a row-indicated change in the predictors’ value. The marginal effects that are supplemented with 95% CI are shown concerning four baselines (healthy, OSA) × (female, male), of specified characteristics from the input. Due to the complex relationship of marginal effect with all Dirichlet dimensions its value changes with the values of predictors (c.f., Eq. 18). Hence, their understanding can be particularly useful in understanding the interplay between different levels of demographics, OSA severity, and particularly their interactions with comorbidities, that have been so far understudied.

Finally, the `Sleep Stage Survival` tab depicts survival curves of individual sleep stages, based on diagonal elements on **P**^*M*^ and Eq. 15. Notably, as this quantity is based on the whole-night **P**^*M*^, survival curves illustrate the overall average duration of individual stages.

## Discussion

Sleep is a complex phenomenon whose finest mechanisms are yet to be fully deciphered. Scoring sleep into a hypnogram of five sleep-wake stages translates it into a simplified, human-readable code, enabling the calculation of PSG markers and their interpretation by clinical personnel. Currently, likely due to fragmented or less intuitive methodologies, the established markers from clinical PSG reports provide only negligible information on sleep dynamics^1,44^. Yet, existing studies provide strong evidence that more granular characteristics of sleep-stage transitions^14–23^ or sleep-stage duration/survival^24–30^ can be specific for various sleep conditions and age. For clinical, economic, and ethical reasons, most of the related research has in common that PSG data were collected in a non-randomised way and were analysed retrospectively, hence subjected to considerable confounding^33^. A minority of studies investigating sleep dynamics addressed confounding either by analyzing subjects with restricted demographic ranges (e.g.,^14,15,24^), or by selecting typically age- and gender-matched controls (e.g.,^16,18,20,30^). This may limit the findings’ generalizability or underfit the age- and gender-specific phenotypes.

By exploiting techniques of causal inference (IPW-balancing from Eq. 25; S-Learner from Eq. 30), our study presents a novel and highly flexible approach to jointly quantify (i) sleep-stage dynamics, (ii) effect of disorder, and (iii) derive several established as well as novel digital markers of sleep. We demonstrate our approach to OSA, the most prevalent sleep condition and a significant risk factor, evidenced to impact sleep macro-strucure and dynamics^19,22,25–28^.

Working with the observational BSDB database, we initially balanced the dataset using IPW-reweighting and addressed the confounding of age and gender, whose distributions differed between healthy and OSA-affected subjects. Ignoring this, it would be challenging to separate the effects of demographics (e.g., of ageing) from OSA, since its prevalence and severity increase with age^28^. To quantify sleep-stage dynamics, we proposed to exploit the matrix **P** (Eq. 1), consisting of 25 (5 × 5) interdependent transition proportions. Thanks to the flexibility of **P** to quantify all, the dynamics, derived markers, and Markovian **P**^*M*^, we suggest considering it as a simple *digital sleep-fingerprint*. All dimensions of **P** were modelled jointly as an outcome of Dirichlet regression (Eq. 17, 30), respecting their compositional nature (summing to 100%) and allowing their straightforward aggregation to derive many established and novel PSG markers (c.f., Eq. 3-13). In contrast, analyzing dependent outcomes, e.g., % of sleep stages and their transitions, separately, such as using (M)ANOVA^22^, would lead to biases and disregard constraints on value ranges and cumulative sums. Considering predictors of age and gender allowed outcome model’s (Eq. 30) adaptation to nonlinear changes in sleep due to ageing and quantification of possible gender phenotypes^2,4,5^. Most importantly, the inclusion of the OSA indicator followed the causal S-learner framework^40^, allowing direct quantification of OSA effects in terms of CATE and RR-CATE (c.f., Eq. 27-28) by comparing expected outcomes for healthy individuals of given demographics with hypothetically matched OSA-subject of specified OSA-severity (AHI). Our modelling approach avoids discretization of age and AHI, and hence allows quantification of personalized (up to OSA-severity and demographics) effects/markers, closely aligning the needs of precision medicine. Even so, it is important to recall that the BSDB dataset contained only 2 cases of paediatric OSA (age < 18 years) and therefore, the conclusions should be taken with care when generalizing them to the pediatric OSA-population. Uniquely, the richness of BSDB allowed us to account for interactions between OSA and several other sleep comorbidities - a clinically well-known and relevant fact (c.f., ^45–49^), so far either overlooked (e.g.,^19,25^), being admitted but not handled (c.f., ^28^), or leading to analysis of subjects with no sleep-comorbidities (e.g.,^22,26^). With 48.6% of OSA subjects in our observational dataset having at least one additional sleep comorbidity, addressing these interactions is crucial for reducing bias and accurately estimating the impact of OSA from other conditions.

The estimated outcome model provides three main dimensions of our results. First, the quantification of sleep fingerprint **P** provides information on the % of time spent in individual transitions and compactness of sleep-stages. Several transitions were significantly increased by OSA for all demographics and AHI-severity groups: W → (N2, N3), N1 →N3, N3 → (N1, REM), and REM → (N1, N3), all peaking with RR-CATE >200%. Despite their rare presence in healthy subjects, our findings suggest they may be a sensitive marker of OSA. In addition, all OSA-F had significantly increased (N1, N2, N3, REM) → W, W → REM, N1 → REM, REM → (W, N2), and decreased REM → REM, suggesting their higher vulnerability to awakenings and REM-disruptions in comparison to M, for whom these effects were observed only partially. This finding may also be linked to more likely REM-OSA in F^50^. Secondly, by aggregating dimensions of **P**, one can derive standard PSG markers (e.q., % of sleep-stages), and many novel proposed ones, that may be specific to particular conditions. For all demographic and AHI groups, OSA significantly increased NREM-REM oscillations (c.f., Eq. 7), overall sleep-stage fragmentation (c.f., Eq. 12), and (N1, N2, N3, REM)-specific fragmentations (c.f., Eq. 13). In addition, all, sleep-, light-sleep, and deep-sleep-awakenings (c.f., Eq. 3-5), were increased for all moderate and severe-OSA groups. Finally, row-normalizing **P** yields the Markovian **P**^*M*^, which quantifies the probabilistic distribution of the next phase given the current state, thus investigating deeper dynamic mechanisms. For all age and AHI groups, OSA increased N1 → N3, N3 → REM, REM → (N1, N2, N3), and decreased REM → REM and N2 → N2. All moderate and severe OSA had also increased N3→ N1 and decreased N3 →N3. For all OSA-M, an additional increase in W → (N2, N3, REM) and for all OSA-F increase in N1→ (W, REM), (N2, REM) →W and decreased N1→ N1 was observed. Furthermore, we demonstrated that **P**^*M*^ can also be used to model sleep-stage survival (Eq. 15), bridging the two principal directions of sleep dynamics research: sleep-transitions^14–23^ and sleep-stage bout duration quantification^24–30^. The merit of the stage survival analysis includes the evaluation of the functional form of the distribution. We can learn their statistical property which provides insights into the underlying mechanism.

To underscore the diagnostic utility of our findings, we evaluated the predictive power of individual transition proportions in **P** on external data from SHHS, containing a broad population of subjects from the general public. For each transition, we developed a simple logistic regression model using age, gender, and the specific transition percentage as predictors, and assessed its ability to identify moderate sleep-disordered breathing (AHI > 15). Results showed significant predictive utility across all 25 transitions, with all AUROC values exceeding 50% (range of 59.82-69.28), and their average of 66.77% for SHHS1 and 65.98% for SHHS2, with respective standard deviations of 1.79% and 1.82%. This robust performance highlights the generalizability of the derived markers from the BSDB dataset to a broader population while confirming the informativeness of individual transitions as predictors of sleep-disordered breathing. Higher predictive performance can be expected when including additional predictors not reflected in **P** (e.g., total-sleep-time, sleep-latency), their interactions with specific proportion, using all proportions jointly, or using a more complex predictive model than logistic regression.

In summary, our findings from different perspectives confirm that OSA is associated with reduced continuity of N2, N3, and REM sleep, reflected by increased sleep fragmentation^19,22,25–28^. By exploiting the matrices **P** and **P**^*M*^, we identified OSA-specific transitions contributing to this fragmentation, particularly atypical transitions from light to deep sleep and oscillations between N3 and REM. These transitions, though rare in healthy individuals, may serve as sensitive markers of OSA, possibly reflecting compensatory mechanisms where the body attempts to acquire back the restorative states, after their frequent disruption by apneic events. Additionally, we proposed several intuitive markers that aggregate dimensions of **P**, demonstrating their potential to distinguish between disordered and healthy subjects. The results of our work are also available as an interactive app, allowing in-depth exploration of results and proposed markers for arbitrary demographics, OSA-severity, and their interactions with other sleep-comorbidities.

Our approach to support diagnostics, has broader applicability beyond the OSA use-case, as sleep dynamics and their markers can be specific to other sleep disorders, such as narcolepsy, insomnia, periodic limb movement disorder, and others. With the rise of telemedicine and increasing use of wearables, investigating sleep dynamics and its markers could become a valuable screening tool for assessing the risk of psychiatric (e.g., depression, schizophrenia, etc.) and neurodegenerative disorders (e.g., Parkinson’s disorder, Alzheimer’s disease, etc.), which are evidenced to be associated with disrupted sleep^51–53^. Even though the consumer devices provide - compared to clinical PSG - lower quality signals and hypnograms, adaptation and re-estimation of our approach to these data has still great potential to provide valuable insights. Furthermore, with advances in automatic sleep-scoring tools that offer hypnodensity beyond the standard hypnogram^54^, our framework could enhance the understanding of sleep micro-events and more granular sleep dynamics, when hypnograms on less than 30-second windows would be used as data for our model. Our future work will extend our approach to address several of its limitations. Following ideas of^21^, we aim to extend it to the second-order sleep-stage transitions that would require quantifying a 125 (= 5 x 25) dimensional transition cube. Next, we plan to account for time spent asleep and investigate dynamics at different times of the night. Currently, we have focused on transitions aggregated over the entire sleep period, but recognizing the non-stationary nature of sleep offers opportunities for identifying even more specific markers. This would also concern the quantification of sleep-stage survival or duration, which our current work approximated by an overall night expectation. Next, we plan to consider whether the subject’s apnea events are REM- or NREM-dominant, which may reveal additional phenotypes. And lastly, we plan to investigate in greater detail the interaction of OSA with comorbidities, which can already be explored in our app.

## Methods

This section details the study data, introduces the matrix of sleep-stage transition proportions as a foundational digital marker, and explores its properties alongside several novel sleep markers. Additionally, we outline the technical framework, which leverages causal inference tools to minimize bias in the conclusions of this observational study, and present a use case examining the effects of OSA.

### Data

#### Berner Sleep Data Base (BSDB)

For the primary evaluations (such as estimating the effects) of our study, we exploited the clinical *Berner Sleep Data Base* (BSDB) from Inselspital, University Hospital Bern. We considered a subset of 62 healthy subjects (aged 0-71 years) with excluded existing clinical conditions undergoing PSG as controls in several historical studies, and a total of 560 individuals having OSA (aged 2-81 years, including 2 pediatric cases aged < 18 years) as one of their conclusive diagnoses, made by physicians considering all test-based diagnoses (e.g., actigraphy- or PSG-based), clinical anamnesis, and the context. The PSG signals were recorded at 200 Hz and scored manually according to the AASM rules^1^. To align older recordings scored by Rechtschaffen and Kales^55^ rules with AASM standard, N3 and N4 stages were merged into N3. To prevent bias due to possibly longer sleep-onset in the unfamiliar clinical setting, a part of the PSG recording and hypnogram before the first sleep was cut off. Further, recordings with total sleep time <180 minutes, >5% of the time with lights-on, no sleep-stage transitions, and subjects with breath control or ventilation therapy introduced, or undergoing split night PSG evaluations were excluded. For the basic statistical description of BSDB in Table 2, we considered 3 groups of OSA subjects: mild (O1) with AHI ∈ [5, 15), moderate (O2) with AHI ∈ [15, 30), and severe (O3) with AHI ≥ 30.

Most sleep metrics and demographics differ significantly between healthy individuals and OSA groups, as well as across different OSA severity levels. There is a clear trend of increasing age and % of males from healthy to more severe OSA, which is also associated with changes in sleep architecture, such as decreased sleep efficiency and reduced N3 and REM %. Separating the effects of these demographic shifts from the effects of OSA is a key challenge, addressed using a causal inference below.

#### Sleep Heart Health Study (SHHS)

The Sleep Heart Health Study (SHHS) is a large, multi-centre cohort study designed to investigate the relationship between sleep-disordered breathing and cardiovascular outcomes^56,57^. SHHS1 includes baseline polysomnography (PSG) data collected from 5,804 unique subjects aged 39–90 years, while SHHS2 provides follow-up PSG data for 2,651 subjects aged 44–90 years. Following the same criteria as in BSDB, we included only subjects with total sleep time (TST) > 180 minutes. After this selection, SHHS1 retained 5,734 subjects (mean age 63.1 ± 11.2 years, 47.6% male), and SHHS2 included 2,621 subjects (mean age 67.5 ± 10.3 years, 46.1% male).

SHHS1 and SHHS2 were utilized to independently evaluate the predictive power of individual sleep-stage transition proportions, forming the foundation for deriving novel sleep markers, in identifying subjects with sleep-disordered breathing. These analyses provide robust external validation of the effectiveness of these transition proportions in the predictive task, which underscores their clinical relevance.

For both BSDB and SHHS datasets, the definition of the Apnea-Hypopnea Index (AHI) used aligns with the National Sleep Research Resource (NSRR) harmonization^57^: AHI = (All apneas + hypopneas with ≥30% nasal cannula [or alternative sensor] reduction and ≥3% oxygen desaturation or with arousal) per hour of sleep, which follows clinical guidelines^1^.

### Matrix P of sleep-stage transition proportions: a basic sleep marker

Our framework proposes the use of a flexible digital marker—a sleep fingerprint—that, based on the observed sleep stages of a subject, enables the derivation of both established and novel PSG parameters, quantifying various sleep characteristics that may be specific to different sleep conditions. The basis for achieving this is the hypnogram, which represents the sequence of sleep-wake stages (W, N1, N2, N3, REM) throughout the night. While sleep dynamics in clinical PSG reports are currently limited to the total counts of transitions and awakenings, this can be easily extended by the 5 x 5 matrix of sleep-stage transition proportions **P**. Let us denote the total number of epochs in the patient’s hypnogram (starting from sleep-onset) as *N*^*E*^, and the number of transitions from stage *i* to *j* as *N*^*ij*^. Each cell *p*_*ij*_ of **P** can then be expressed as:

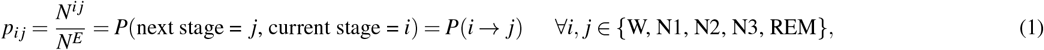

indicating the empirical probability (proportion, %) of observing a transition from stage *i* to *j* (*i* → *j*), relative to all the transitions observed in the hypnogram. In the following, we highlight three main dimensions of the clinical relevance of **P**.

#### P *recovers the majority of clinically established PSG markers*

For example, summing up the column transition proportions of **P** yields the overall percentage of sleep stages:

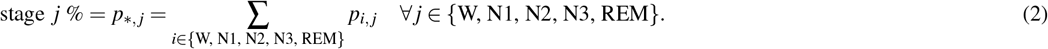

In addition, other clinically commonly used PSG markers can be easily derived by considering relevant proportions and the *Total Sleep Time* (TST),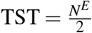, in minutes. For example, *Sleep Efficiency* (SE), quantifying the percentage of sleep after its onset, can be calculated as SE = ∑ _*j*∈{N1,N2,N3,REM}_ *p*_*,*j*_ = 1 − *p*_*,*W*_ . The *Wake After Sleep Onset* (WASO) minutes can be computed as 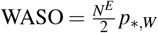. The *Number of Awakenings* (NoA) can be determined by NoA = *N*^*E*^∑ *i*∈{N1, N2, N3, REM}, *p*_*i*_,_*W*_ . Finally, the *Number of Transitions* (NoT) is given by NoT = *N*^*E*^ ∑*i*∈{W, N1, N2, N3, REM}(1 − *pi*,*i*).

#### P *allows derivation of novel PSG markers*

The aggregation of **P**-dimensions offers a great flexibility to derive several novel and highly intuitive digital markers of sleep and its dynamics. Considering a set of sleep-states, 𝒮 = {N1, N2, N3, REM}, we propose and in results also evaluate the following.

*Total Awakenings*, the probability of transitioning from any sleep-state (𝒮) to wakefulness:

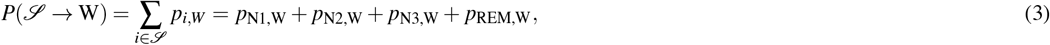

*Light-sleep Awakenings*, the probability of transitioning from light sleep (N1, N2) to wakefulness:

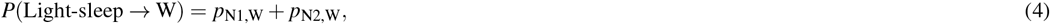

*Deep-sleep Awakenings*, the probability of transitioning from deep sleep (N3) to wakefulness:

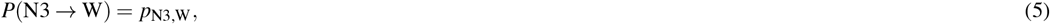

*REM Awakenings*, the probability of transitioning from REM sleep to wakefulness:

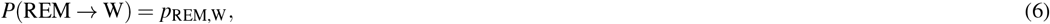

*NREM-REM Oscillations*, sum of probabilities for transitions between NREM sleep stages and REM sleep:

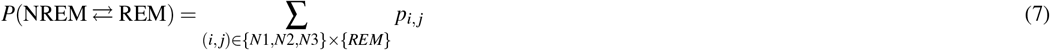

*Light-sleep Oscillations*, sum of probabilities for transitions between the light sleep stages (N1 a, N2):

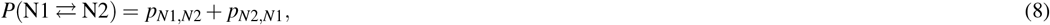

*Sleep Compactness*, the total probability of staying within any (non-wake) sleep stages:

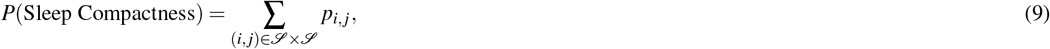

*Sleep Fragmentation*, the total probability of switching between wakefulness and sleep states:

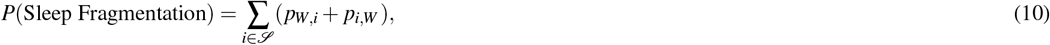

*Sleep-stage Compactness*, the sum of probabilities of staying within the same (non-wake) sleep stages:

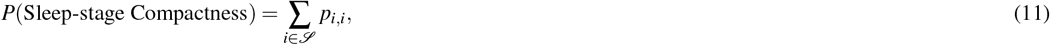

*Sleep-stage Fragmentation*, the probability of transitioning from one (non-wake) sleep stage to a different one:

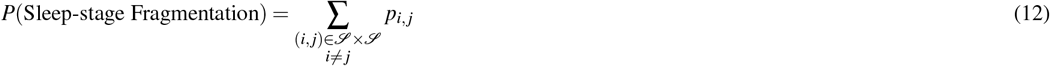

*Stage-specific Compactness and Fragmentation*, for each sleep stage *i*, the probability of staying in the same stage and the probability of switching to any other sleep stage, respectivelly:

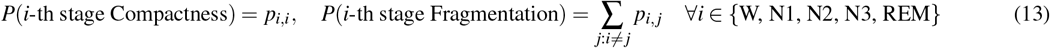

Each metric from Eq. 3-13 expands the standard clinical PSG markers and focuses on a specific sleep pattern. Their quantification requires no additional effort once the subject has undergone the PSG study and the hypnogram is available.

#### P bridges stage-transitions and durations-oriented sleep dynamics research

Normalizing **P** so that each row sums to 1 (100%) yields a standard transition matrix, often utilized in Markovian models. We denote this matrix as **P**^*M*^, where *M* indicates it is Markovian. Each cell,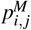, corresponds to the conditional probability of transitioning to stage *j* after being in stage *i*:

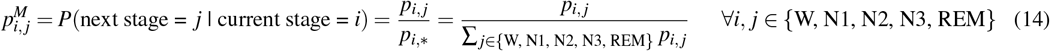

The key difference is that while **P** provides an overall view of the plausibility of individual transitions, **P**^*M*^ operates under the assumption that a given state has occurred and problematically evaluates the chances of (not-)switching the sleep-stage in the next epoch. Both **P** and **P**^*M*^ are interconnected and offering two perspectives on sleep-stage dynamics. Notably, the diagonal elements of **P**^*M*^ enable straightforward quantification of the sleep-stage durations, as they are exponentially distributed, 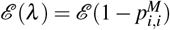, with the expected duration (ED) of each stage (over entire night):

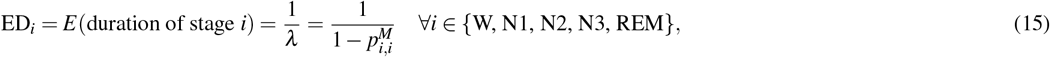

known as the mean sojourn time. Due to the scoring of sleep in 30-second windows, these durations are measured in epochs.

### Causal framework to quantify sleep-stage transition matrix P and effects of a disorder

The preceding sections have highlighted the utility of investigating the matrix **P** as a sleep-fingerprint, showing its relation to several clinically established PSG markers and its connection between stage-transition and stage-duration sleep dynamics research. Moreover, we introduced several novel markers derived from **P**. To quantify **P** and the derived markers, the next sections will present an approach that combines Dirichlet regression, well-suited for the compositional data of **P**, with elements of causal inference to address confounding. The key challenge in modeling **P** lies in respecting the compositional nature of the data, where the total of all percentages must sum to 100%. Ignoring this constraint, such as analyzing particular proportions separately with ANOVA, can lead to significant bias and counterintuitive outcomes. This issue is evident in some meta-analyses where, for example, aggregated percentages of sleep stages do not sum to 100%, as seen in Table 2 of ^7^. This challenge must be addressed when modeling the proportions of sleep-stage transitions in **P**, which involve 25 compositional dimensions. Ensuring the outcomes are intuitive and correct is crucial for enabling their interpretation by medical professionals.

#### Dirichlet regression: model formulation and properties

The Dirichlet distribution is well-suited for modeling compositional data, such as percentages or the elements of **P**. For a random variable *Y* = (*Y*_1_,*Y*_2_, …,*Y*_*D*_) representing proportions over *D* dimensions, the probability density function of the Dirichlet distribution is parameterized by a vector of positive reals *α* = (*α*_1_, …, *α*_*D*_) and given by:

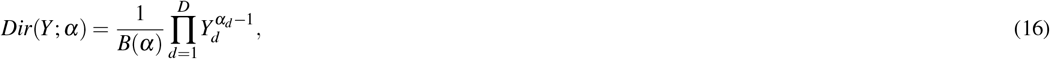

where *B*(*α*) is the multivariate beta function ensuring normalization^39^. In Dirichlet regression, the logarithms of *α* are modeled as functions of covariates, adapting the distribution’s characteristics based on predictor values:

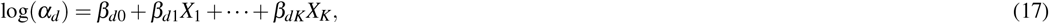

where *X* = (*X*_1_, …, *X*_*K*_) is a set of *K* covariates and *β*_*d*_ = (*β*_*d*0_, …, *β*_*dK*_) a vector of regression coefficients for the *d*-th dimension. The expectation of each component *Y*_*d*_, *E*[*Y*_*d*_], and the marginal effect of *X*_*j*_ on 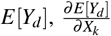, are directly influenced by all elements of *X* and *α*, reflecting the interdependencies of compositional data:

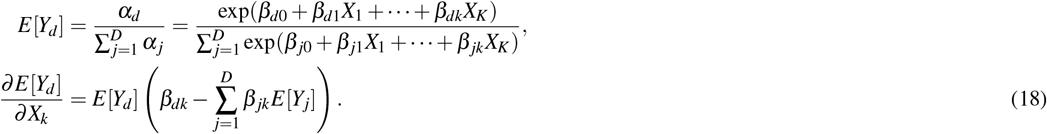

A convenient property of the Dirichlet distribution is its ability to aggregate over several dimensions, allowing flexible quantification of measures based on the elements’ summation. For example, aggregating dimensions *i* and *j* yields:

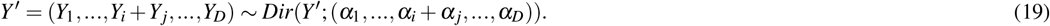

Thus, Dirichlet regression is suitable for modelling **P**, and its aggregation property facilitates straightforward quantification of all markers derived from it (c.f., Eq. 2-13).

#### Causal elements

In contrast to randomized experiments, the analysis of observational data, such as those from PSG databases, is susceptible to confounding, due to varying distributions of characteristics (e.g., age), between treated/exposed/conditioned and healthy-control subjects. Our study, which aims to quantify changes in sleep parameters resulting from a sleep disorder, adopts the principles and standard notation of causal inference^41^. We define the *treatment/exposure/condition* variable *T* as an indicator of whether a subject suffers from a particular disorder of interest (*T* = 1), or is a healthy control (*T* = 0). **In line with the language of causal inference, the treatment within our study corresponds to the presence of OSA**. The *outcome* (*Y*) represents the sleep parameter investigated, such as **P**, while subject characteristics and potential confounders are denoted as *X* .

### Potential outcomes framework and causal estimands

The potential outcomes framework asserts to each individual two hypothetical outcomes: *Y* (1), under *T* = 1, and *Y* (0), without exposure, *T* = 0. The *Individual Treatment Effect* (ITE), *τ*_*i*_, is the difference between these outcomes, evaluating the causal effect of exposure (e.g., OSA) on subject’s outcome (e.g., sleep):

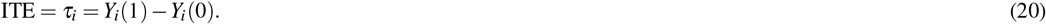

The *Average Treatment Effect* (ATE) is the expected ITE, assessing the effect of *T* across the entire population:

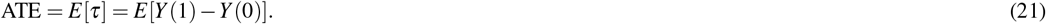

The *Conditional Average Treatment Effect (CATE)* assesses *τ*(*x*), standing for the treatment effect within a specific subgroup of the population characterized by covariates *X*, making it suitable to quantify personalized markers for different conditions:

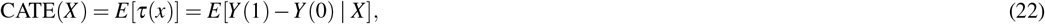

The *fundamental problem of causal inference* is that only one of the two potential outcomes is observed for each individual, according to their treatment/exposure assignment *T*_*i*_:

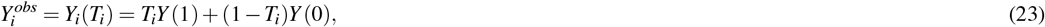

making it impossible to directly calculate all hypothetical estimands (ITE/ATE/CATE) from observed data (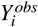, *T*_*i*_, *X*_*i*_).

### Personalized markers using CATE estimates

To estimate (C)ATE from observational data, advanced techniques are required to adjust for confounders and mimic a randomized experiment setting. One method exploits *Propensity Scores* (PS):

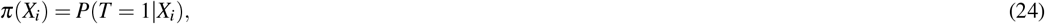

assessing the probability of receiving treatment given the individual’s characteristics *X* . Adjusting for PS removes biases associated with included covariates^36^. In addition, by assuming positivity (i.e., all confounder values can be observed in both treated and controls) and no unobserved confounders, the treatment and potential outcomes become independent conditional on *π*(*X*_*i*_), *T ⊥ Y* (0),*Y* (1) | *π*(*X*), allowing straightforward effect estimation by matching or regressing the outcome on PS^37^.

Another approach, *Inverse Probability Weighting* (IPW), balances the distribution of *X* across treated and controls by creating a pseudo-population where each original subject is re-weighted using weights:

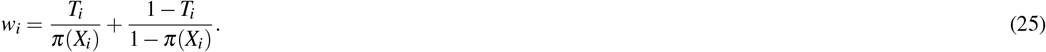

The weights can be, for example, incorporated into flexible, even machine-learning-based, outcome models (e.g., weighted regression) to estimate the treatment effect while mitigating selection bias^38^.

In our study, focusing on quantifying the effects of OSA (*T* = 1) on **P**, we employ IPW within the S-learner framework^40^. The S-learner is a baseline approach of meta-learners, enabling flexible estimation of heterogeneous CATE. The S-learner quantifies the outcome using a single model (hence *S*-Learner), including the treatment indicator *T* as one of its predictors:

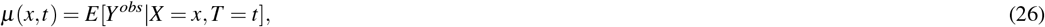

allowing straightforward estimation of CATE from Eq. 22 that is easily extrapolated over the entire range of *X* :

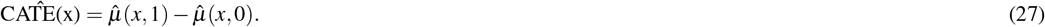

For probabilistic outcomes, the Risk-Ratio CATE (RR-CATE) is preferred as it naturally compares the chances of an event:

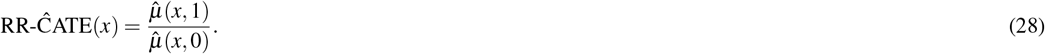

One of the key benefits of S-Learner is its simplicity in extrapolating the (RR-)CATE estimates over and beyond the observed values of *X* . Unlike other meta-learners (e.g., T- or X-learner^40^) that fit separate response functions for exposed (*T* = 1) and control (*T* = 0) subjects, the S-learner estimates a single model and thus requires less data, while assuming that the effects of the other (non-treatment) variables are shared within groups.

### Practical considerations

Care must be taken in interpreting causal effects due to assumptions underlying PS (and so IPW), such as no unobserved confounders and positivity. These assumptions are challenging to validate rigorously. In summary, addressing confounding is better than ignoring it, but interpretations should consider the assumptions made.

### Study use case: effects of OSA on sleep-stage transitions matrix P and derived markers

The practical part of our study links the proposed sleep fingerprint **P** (c.f. Eq. 1) and derived markers (c.f., Eq. 2-13 and Eq. 14) to a causal framework for their efficient quantification and estimation of disorder effect. We demonstrate our approach on OSA, the most prevalent sleep disorder and a significant risk factor, and exploit study dataset from BSDB.

To model PS from Eq. 24, we applied the logistic regression including confounders the most frequently occurring in the literature: age and gender. Both factors are also known to impact the risk of OSA and at the same time, their value range is not constrained between OSA and healthy subjects, thus meeting the positivity assumption. The PS model included separate predictors of the scaled age above 50 years in decades (*X*_(Age−50)*/*10_), gender indicator (𝕀_male_), and their interaction:

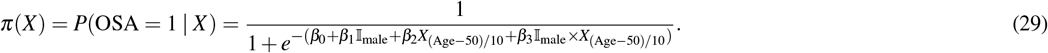

The IPW weights based on Eq. 25 were used to balance the data concerning the main confounders shared.

To estimate the effects, i.e., (RR)-CATE from Eq. 27-28, of OSA on the compositional outcome of **P**, the Dirichlet regression, as introduced in Eq. 16-17, was exploited to model the response within the S-learner framework from Eq. 26. Each of the 25 possible transition proportions captured in **P** and indexed as (*i, j*) *∀i, j* ∈ { W, N1, N2, N3, REM }, was modelled using the predictor specific for the corresponding dimension characterized by *α*_(*i*,*j*)_:

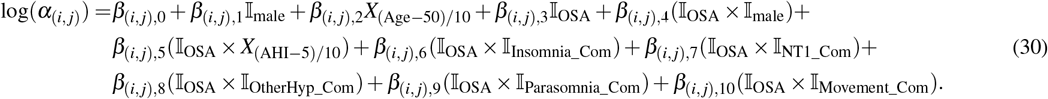

This log-transformed *α*_(*i*,*j*)_ was regressed on several covariates and interaction terms with a primary goal to separate and quantify the effect of OSA, present as an indicator variable I_OSA_. Although this S-learner model was estimated on IPW-balanced data (c.f., Eq. 29), the inclusion of age and gender was justified by the necessary adjustment due to their known influence on sleep manifestation. Next, the interaction of OSA with gender was also included, to investigate potential gender-specific phenotypes. In addition, several variables that violating the positivity assumption were included, as they could not be utilized within the PS model due to their disjoint distributions among healthy and OSA subjects. This included the interaction terms of OSA with scaled Apnea Hypopnea Index (AHI), *X*_(AHI −5)/10_, denoting number of AHI greater than 5 in tens, capturing the apnea severity as the number of breath-arrests per hour. Uniquely, our model adjusts for a comprehensive range of comorbidities present as indicator variables: insomnia (𝕀_Insomnia_Com_), Narcolepsy Type 1 (NT1, 𝕀_NT1_Com_), other hypersomnolence except NT1 (𝕀_OtherHyp_Com_), parasomnias (𝕀_Parasomnia_Com_), and movement-related sleep-disorders (𝕀_Movement_Com_). The distribution of AHI and all the comorbidities is completely disjoint, as healthy subjects do not suffer from any disorder/comorbidity and AHI values in OSA subjects are always greater than 5.

To assess uncertainty and calculate confidence intervals (CI) in all strands of our investigations, including the PS model, IPW-balanced S-learner with Dirichlet regression, and subsequent quantification of **P**-derived markers using (RR)-CATE, we implemented a non-parametric bootstrap procedure with 200 repetitions, inspired by^58^.

## Acknowledgements

The secondary usage of Berner Sleep Data Base (BSDB) from Inselspital, University Hospital Bern, was approved by the local ethics committee (KEK-Nr. 2022-00415), ensuring compliance with the Human Research Act (HRA) and Ordinance on Human Research with the Exception of Clinical trials (HRO), and analyzed in the framework of the E12034 - SPAS (Sleep Physician Assistant System) Eurostar-Horizon 2020 program. The BSDB dataset access may be granted upon individual request, after data transfer agreements were put in place.

## Author contributions

M.B. conceptualized the study, developed the methodology, performed the analysis, drafted the manuscript, and incorporated feedback from all co-authors. A.K. provided detailed feedback on related work, and clinical interpretation of the results, and contributed to the discussion section. J.v.d.M. assisted with data curation and provided detailed feedback on the introduction and discussion sections. L.F., M.S., C.B., A.T., and F.F. read the manuscript and provided their feedback. All co-authors approved the final manuscript and agreed to be listed as co-authors.

## Data availability

The datasets analyzed in this study are subject to different accessibility conditions:

- **Bern Sleep Data Base (BSDB):** Due to patient confidentiality and ethical restrictions, the BSDB dataset is not publicly available. However, de-identified data may be obtained from the corresponding author upon reasonable request, subject to data sharing agreement and approval by the relevant ethics committees.
- **Sleep Heart Health Study (SHHS)**: This dataset is publicly accessible through the National Sleep Research Resource (NSRR) at https://sleepdata.org/datasets/shhs. Researchers can request access by creating an NSRR account and agreeing to the data use terms and conditions.

## Code availability

The underlying code supporting the findings of this study, including the supplementary app for exploring the results and proposed markers, is publicly available at https://github.com/MiBec/Sleep_Dynamics_OSA.

