## Supplementary Materials for "Novel Digital Markers of Sleep Dynamics: A Causal Inference Approach Revealing Age and Gender Phenotypes in Obstructive Sleep Apnea"

January 15, 2025

### Supplementary materials

#### Outcome model of Dirichlet regression

| $\alpha$ | Intercept | $\mathbb{I}_{\text{male}}$ | $X_{(\text{Age}>50)/10}$ | $\mathbb{I}_{\text{OSA}}$ | $\mathbb{I}_{\text{OSA}} \times \mathbb{I}_{\text{male}}$ | $\mathbb{I}_{\text{OSA}} \times X_{(\text{AHI}>5)/10}$ | $\mathbb{I}_{\text{OSA}} \times \mathbb{I}_{\text{Insomnia\_Com}}$ | $\mathbb{I}_{\text{OSA}} \times \mathbb{I}_{\text{NT1\_Com}}$ | $\mathbb{I}_{\text{OSA}} \times \mathbb{I}_{\text{OtherHyp\_Com}}$ | $\mathbb{I}_{\text{OSA}} \times \mathbb{I}_{\text{Parasomnia\_Com}}$ | $\mathbb{I}_{\text{OSA}} \times \mathbb{I}_{\text{Movement\_Com}}$ |
|---|---|---|---|---|---|---|---|---|---|---|---|
| W → W | **1.11*** (0.25, 1.94) | -0.18 (-0.64, 0.09) | 0.00 (-0.10, 0.10) | **-0.59*** (-1.72, -0.19) | **0.50*** (0.12, 1.62) | 0.12 (-1.42, 0.51) | -0.32 (-0.61, 0.36) | 0.01 (-0.29, 0.22) | 0.10 (-0.23, 1.68) | **-0.06*** (-0.11, -0.04) | **0.19*** (0.05, 0.44) |
| W → N1 | 0.65 (-0.33, 2.00) | 0.17 (-0.59, 0.48) | **0.26*** (0.04, 0.65) | 0.07 (-0.80, 0.48) | **0.17*** (0.05, 1.07) | 0.01 (-0.04, 0.06) | **0.34*** (0.09, 0.65) | **-1.34*** (-1.47, -1.12) | **-0.96*** (-1.86, -0.57) | -0.13 (-0.31, 0.02) | **-0.14*** (-0.44, -0.01) |
| W → N2 | **0.74*** (0.28, 2.03) | -0.02 (-0.04, 0.01) | **0.26*** (0.14, 0.48) | **1.18*** (0.80, 1.60) | 0.01 (-0.73, 0.31) | -0.19 (-0.58, 0.06) | **-0.59*** (-1.23, -0.33) | -0.06 (-0.27, 0.59) | 0.17 (-2.32, 0.92) | 0.23 (-0.04, 0.81) | 0.03 (-0.20, 0.23) |
| W → N3 | -0.44 (-2.04, 0.32) | 0.01 (-0.13, 0.18) | **-0.16*** (-0.61, -0.01) | -0.02 (-0.37, 1.51) | -0.40 (-1.64, 0.02) | 0.04 (-0.28, 0.64) | -0.22 (-0.62, 0.09) | -0.00 (-0.05, 0.19) | **-0.23*** (-0.33, -0.12) | **0.33*** (0.17, 0.66) | **-1.59*** (-1.65, -1.50) |
| W → R | -0.49 (-2.21, 0.52) | 0.02 (-0.18, 0.33) | -0.07 (-0.29, 0.08) | **0.14*** (0.03, 0.85) | **-0.04*** (-0.09, -0.00) | **0.48*** (0.22, 0.84) | -0.31 (-0.65, 0.05) | -0.12 (-0.37, 0.05) | **-0.67*** (-1.15, -0.07) | **-0.41*** (-1.18, -0.17) | -0.01 (-0.09, 0.40) |
| N1 → W | -0.06 (-0.11, 0.02) | **0.21*** (0.07, 0.34) | -0.18 (-0.57, 0.24) | **-0.62*** (-1.17, -0.32) | **-0.35*** (-0.59, -0.13) | **-0.71*** (-1.64, -0.34) | 0.03 (-0.29, 1.21) | 0.09 (-0.54, 0.30) | -0.27 (-0.89, 1.21) | -0.17 (-0.53, 0.06) | 0.03 (-0.00, 0.23) |
| N1 → N1 | 0.31 (-0.28, 0.71) | **-0.16*** (-0.41, -0.01) | 0.40 (-0.01, 1.66) | 0.15 (-1.34, 0.54) | **0.48*** (0.18, 1.07) | 0.05 (-0.55, 0.45) | 0.03 (-0.03, 0.47) | **-0.03*** (-0.05, -0.01) | **1.03*** (0.49, 1.58) | **-0.82*** (-1.52, -0.30) | **-0.14*** (-0.26, -0.09) |
| N1 → N2 | **0.82*** (0.39, 1.65) | -0.04 (-0.23, 0.15) | **0.26*** (0.14, 1.07) | -0.02 (-0.07, 0.03) | **0.28*** (0.06, 0.78) | **3.48*** (2.98, 3.98) | **-0.43*** (-0.86, -0.06) | -0.08 (-0.19, 0.04) | **-0.83*** (-2.01, -0.21) | 0.36 (-0.19, 2.02) | 0.01 (-0.46, 0.10) |
| N1 → N3 | **0.51*** (0.13, 1.11) | **-1.52*** (-1.61, -1.39) | -0.25 (-1.03, 0.10) | -0.13 (-0.45, 0.12) | **-0.50*** (-1.44, -0.23) | -0.12 (-0.52, 1.39) | 0.09 (-1.13, 0.44) | -0.09 (-0.24, 0.18) | -0.20 (-0.87, 0.37) | 0.12 (-0.01, 1.19) | **-0.01*** (-0.03, -0.01) |
| N1 → R | **-0.87*** (-1.91, -0.26) | -0.06 (-0.18, 0.43) | -0.25 (-1.48, 0.18) | 0.02 (-0.29, 0.68) | **-0.33*** (-0.82, -0.05) | 0.15 (-0.04, 1.10) | **-0.07*** (-0.11, -0.04) | **0.13*** (0.03, 0.27) | **-1.47*** (-1.60, -1.34) | -0.02 (-0.87, 0.30) | -0.03 (-0.10, 0.02) |
| N2 → W | -0.07 (-0.72, 0.47) | 0.02 (-0.01, 0.23) | 0.01 (-0.03, 0.04) | **0.52*** (0.26, 0.83) | -0.06 (-0.49, 0.44) | **-0.76*** (-1.61, -0.47) | -0.11 (-0.33, 0.06) | -0.03 (-0.25, 0.12) | -0.12 (-0.23, 0.34) | -0.25 (-1.93, 0.25) | 0.04 (-0.02, 0.22) |
| N2 → N1 | **0.59*** (0.21, 1.00) | **-0.20*** (-0.38, -0.11) | 0.09 (-0.12, 0.33) | **-0.76*** (-1.60, -0.36) | -0.03 (-0.45, 1.35) | 0.19 (-1.56, 0.64) | -0.23 (-0.51, 0.28) | -0.02 (-0.19, 0.13) | **0.04*** (0.01, 0.24) | -0.04 (-0.10, 0.01) | **0.10*** (0.05, 0.22) |
| N2 → N2 | 0.18 (-0.18, 1.46) | 0.07 (-0.46, 0.18) | **0.49*** (0.22, 1.03) | -0.04 (-0.65, 0.32) | 0.06 (-0.03, 0.42) | **-0.17*** (-0.24, -0.09) | **0.34*** (0.18, 0.62) | **0.47*** (0.18, 0.84) | **-0.21*** (-0.36, -0.10) | **-0.28*** (-0.52, -0.02) | **-0.12*** (-0.33, -0.04) |
| N2 → N3 | **0.20*** (0.10, 0.96) | **-0.01*** (-0.03, -0.01) | **0.31*** (0.10, 0.77) | **-1.54*** (-1.62, -1.42) | **-0.39*** (-0.91, -0.01) | -0.30 (-0.64, 0.02) | **-0.46*** (-0.93, -0.29) | -0.14 (-0.57, 1.46) | 0.13 (-0.37, 0.25) | 0.25 (-0.14, 0.90) | -0.03 (-0.16, 0.03) |
| N2 → R | **-0.54*** (-1.21, -0.18) | -0.03 (-0.11, 0.03) | **-0.51*** (-1.49, -0.25) | -0.05 (-0.13, 0.37) | 0.08 (-1.43, 0.49) | -0.14 (-0.55, 0.63) | **-0.30*** (-0.61, -0.09) | **0.13*** (0.02, 0.77) | **-0.02*** (-0.04, -0.01) | **0.32*** (0.08, 0.95) | **2.57*** (2.29, 2.99) |
| N3 → W | -0.06 (-1.36, 0.32) | 0.06 (-0.01, 0.25) | -0.04 (-0.87, 0.32) | 0.03 (-0.01, 0.23) | **-0.04*** (-0.08, -0.00) | **0.52*** (0.22, 0.96) | **-0.80*** (-1.05, -0.44) | -0.32 (-0.79, 0.02) | -0.04 (-0.11, 0.02) | **-0.47*** (-1.47, -0.23) | -0.14 (-0.48, 1.26) |
| N3 → N1 | -0.02 (-0.06, 0.02) | **0.12*** (0.07, 0.23) | **1.65*** (1.04, 2.06) | **-0.17*** (-0.32, -0.09) | 0.11 (-0.08, 0.29) | **-0.93*** (-1.97, -0.39) | -0.15 (-0.55, 0.88) | 0.05 (-1.78, 0.50) | 0.03 (-0.04, 0.20) | -0.24 (-0.68, 0.10) | 0.01 (-0.12, 0.57) |
| N3 → N2 | 0.08 (-0.16, 0.31) | **-0.13*** (-0.33, -0.07) | 0.13 (-0.35, 1.50) | 0.06 (-0.43, 0.15) | 0.06 (-0.25, 0.60) | -0.30 (-1.07, 0.17) | 0.07 (-0.05, 0.77) | **-0.13*** (-0.19, -0.08) | **0.11*** (0.04, 0.23) | **-0.98*** (-1.24, -0.68) | **-1.19*** (-1.97, -0.76) |
| N3 → N3 | **0.36*** (0.10, 0.93) | -0.01 (-0.15, 0.09) | **0.33*** (0.13, 1.40) | **-0.01*** (-0.03, -0.00) | **0.36*** (0.14, 0.62) | **0.65*** (0.36, 1.00) | **-0.42*** (-0.89, -0.18) | -0.27 (-0.56, 0.01) | **-0.12*** (-0.34, -0.06) | 0.02 (-0.31, 1.08) | 0.28 (-1.23, 0.74) |
| N3 → R | **0.46*** (0.26, 0.85) | **-1.42*** (-1.58, -1.21) | **-0.90*** (-2.21, -0.52) | -0.04 (-0.10, 0.03) | **-0.39*** (-0.82, -0.17) | -0.16 (-0.55, 1.47) | 0.03 (-1.16, 0.45) | -0.45 (-0.74, 0.28) | -0.02 (-0.16, 0.07) | -0.05 (-0.11, 0.18) | **-0.15*** (-0.22, -0.08) |
| R → W | **-0.55*** (-1.46, -0.30) | 0.05 (-0.16, 0.91) | 0.19 (-1.21, 0.69) | 0.02 (-0.06, 0.18) | -0.12 (-0.47, 0.11) | **0.12*** (0.01, 0.78) | **-0.06*** (-0.10, -0.04) | **0.31*** (0.06, 0.62) | **-0.82*** (-1.10, -0.51) | **-0.31*** (-0.68, -0.04) | **-0.42*** (-0.92, -0.01) |
| R → N1 | -0.05 (-0.67, 0.28) | 0.03 (-0.04, 0.33) | 0.03 (-0.02, 0.09) | **0.13*** (0.07, 0.24) | **0.68*** (0.29, 1.07) | **-0.35*** (-0.85, -0.03) | -0.09 (-0.28, 0.10) | **-0.60*** (-1.19, -0.32) | 0.22 (-0.18, 1.54) | -0.01 (-1.06, 0.35) | 0.08 (-0.62, 1.10) |
| R → N2 | **-1.08*** (-1.32, -0.71) | **-0.19*** (-0.47, -0.05) | -0.12 (-0.46, 0.19) | **-0.10*** (-0.32, -0.02) | 0.06 (-0.35, 1.66) | 0.04 (-1.72, 0.47) | 0.08 (-0.17, 0.49) | -0.24 (-0.67, 0.09) | **0.11*** (0.01, 0.73) | **-0.04*** (-0.07, -0.02) | **0.76*** (0.29, 1.16) |
| R → N3 | -0.21 (-0.52, 0.51) | -0.04 (-0.89, 0.19) | **0.74*** (0.41, 1.41) | 0.01 (-0.12, 0.12) | **0.14*** (0.01, 0.85) | **-0.13*** (-0.19, -0.09) | **0.31*** (0.14, 0.68) | **2.76*** (1.65, 3.17) | -0.26 (-0.72, 0.03) | -0.08 (-0.24, 0.04) | **-0.68*** (-1.48, -0.13) |
| R → R | -0.04 (-0.11, 0.16) | 0.00 (-0.02, 0.02) | **0.50*** (0.13, 0.99) | **-0.91*** (-1.29, -0.58) | **-0.50*** (-1.05, -0.16) | -0.26 (-0.55, 0.01) | **-0.25*** (-1.11, -0.05) | -0.46 (-1.10, 1.20) | -0.12 (-1.42, 0.25) | -0.08 (-0.25, 0.26) | -0.19 (-1.05, 0.31) |

Table 1: Summary of estimated coefficients together with bootstrapped 95% confidence interval for the Dirichlet regression outcome model from Eq. 30. Significant estimates are highlighted in bold *. The rows correspond to individual dimensions, specific to each of the 25 possible sleep-stage transitions.

#### Comparison based on matrices of transition proportions P

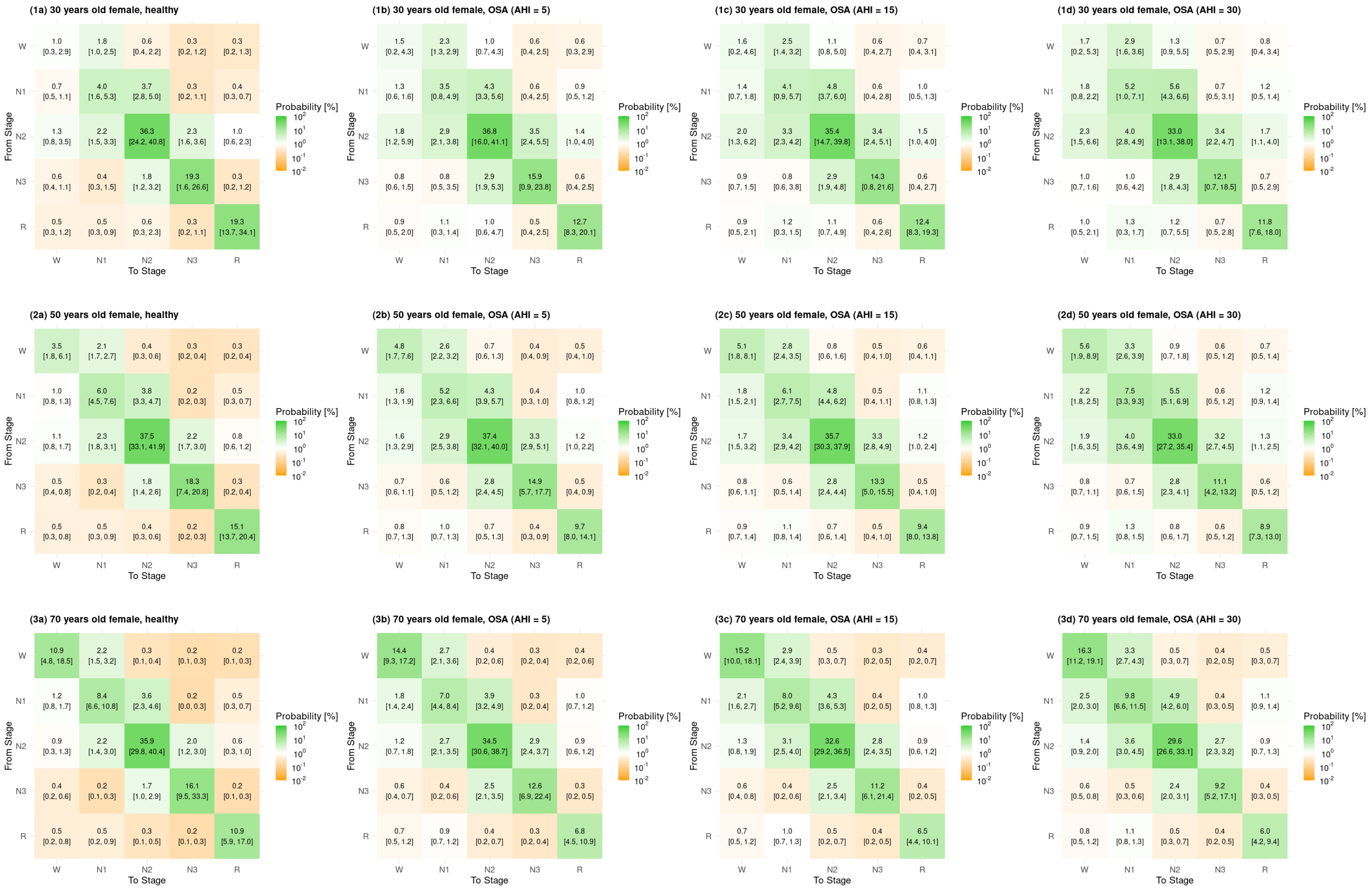


Figure 1: Expected matrices of transition proportions $\mathbf{P}$ for healthy females and females with OSA (AHI = 5, 15, 30) at different ages (30, 50, 70 years). Estimates are supplemented with 95% bootstrapped confidence intervals.

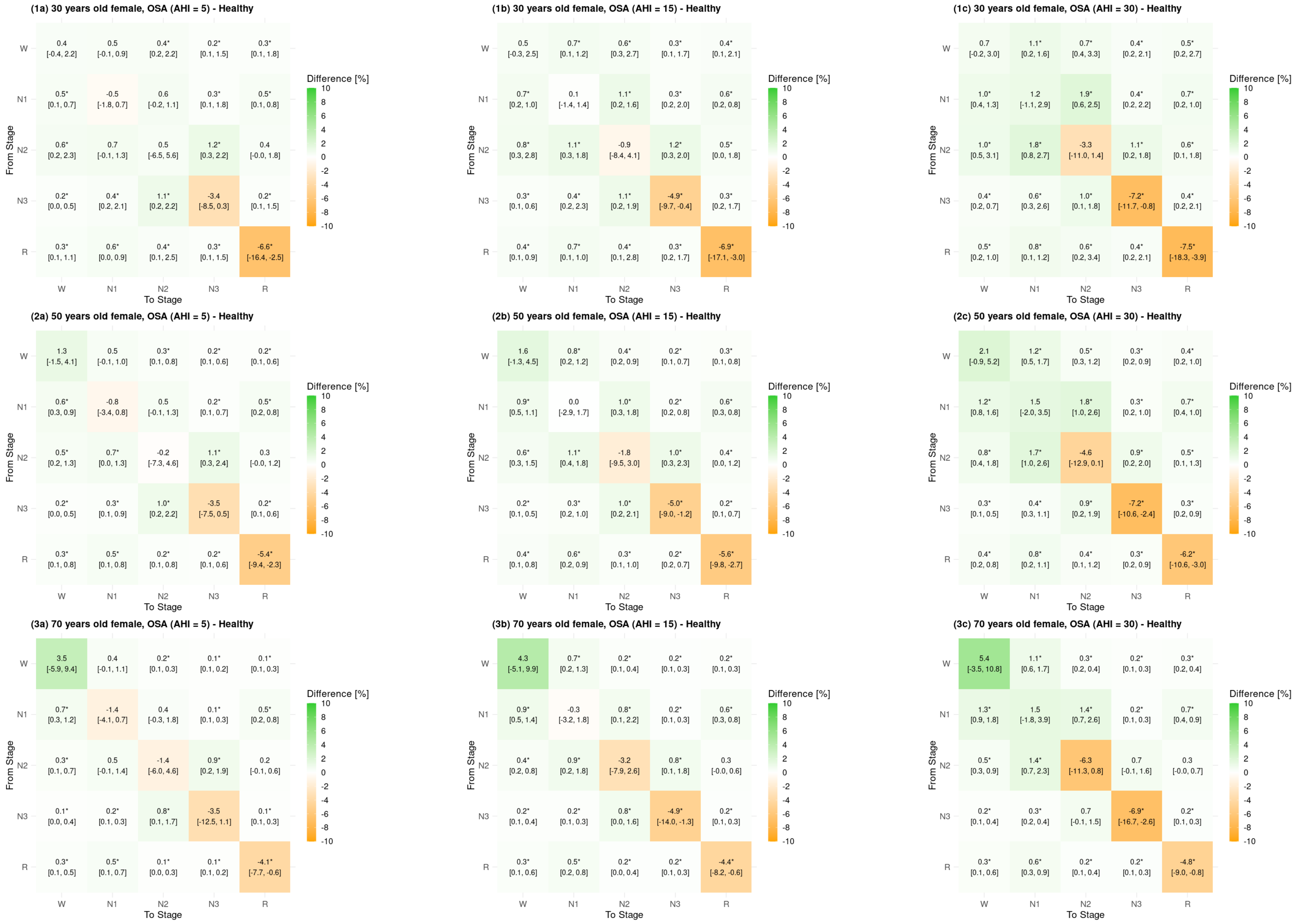


Figure 2: Difference (CATE) of matrices of transition proportions **P** for healthy females versus females with OSA (AHI = 5, 15, 30) at different ages (30, 50, 70 years). Estimates are supplemented with 95% bootstrapped confidence intervals.

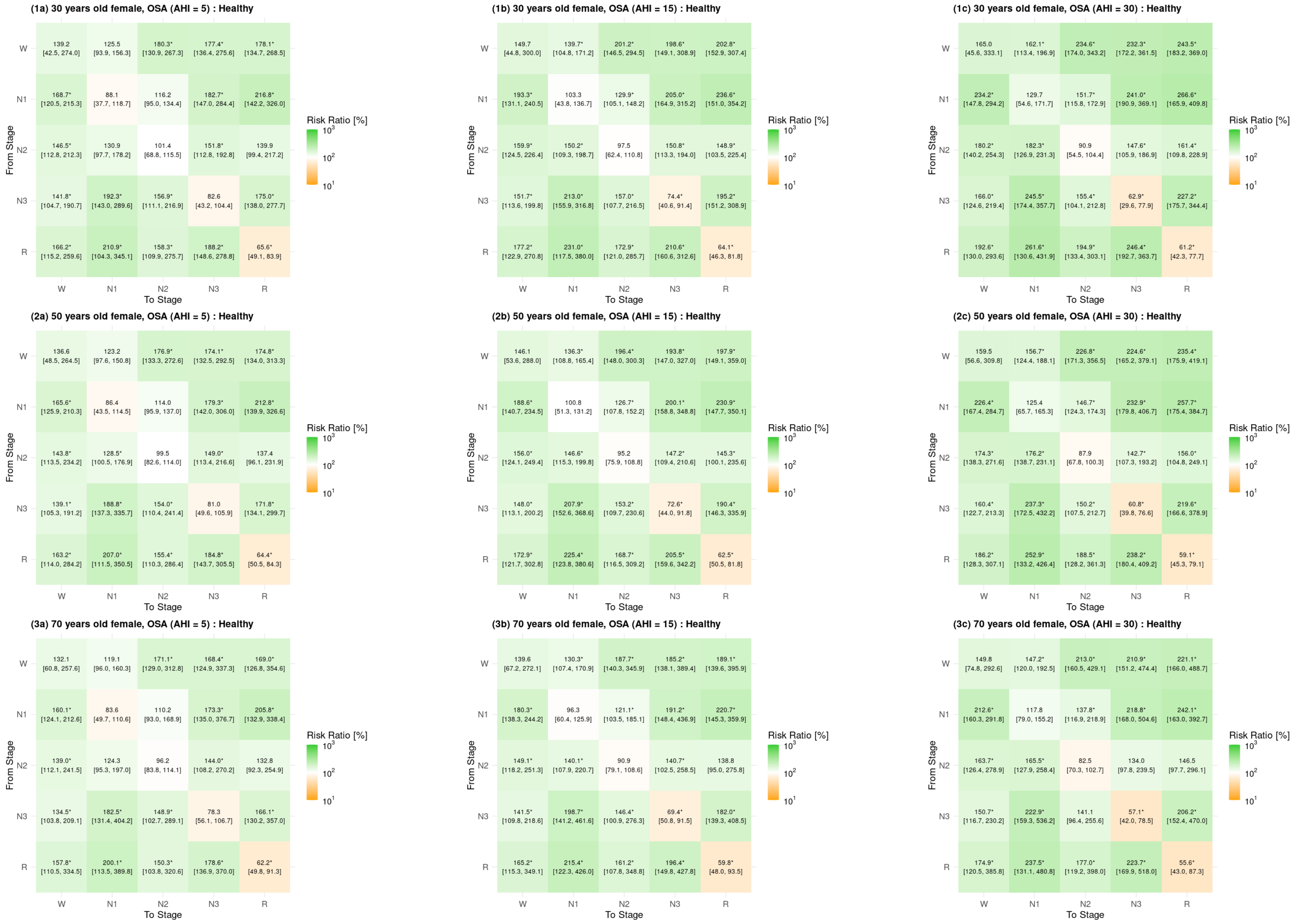


Figure 3: Risk ratio (RR-CATE) of matrices of transition proportions $\mathbf{P}$ for healthy females versus females with OSA (AHI = 5, 15, 30) at different ages (30, 50, 70 years). Estimates are supplemented with 95% bootstrapped confidence intervals.

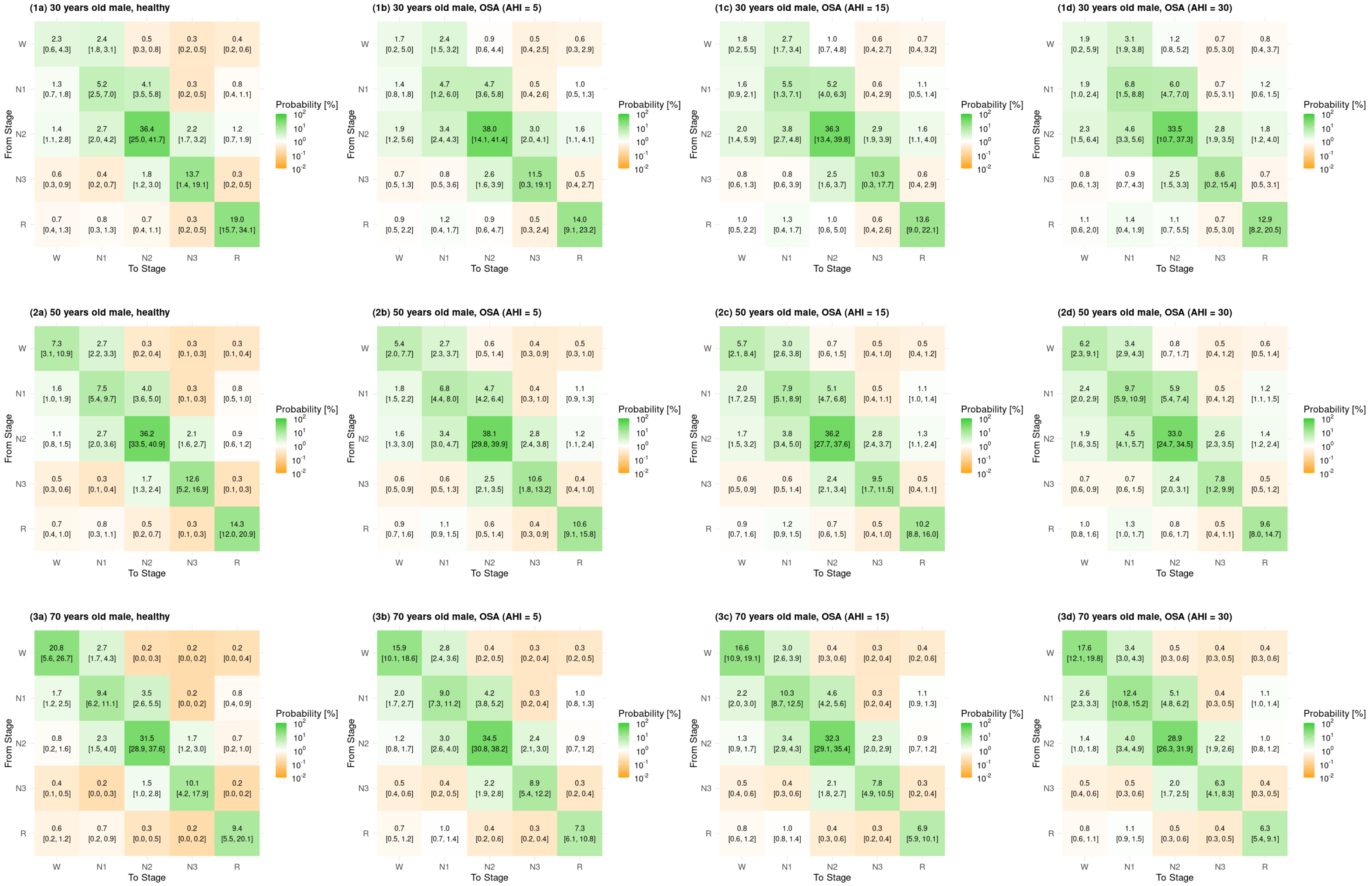


Figure 4: Expected matrices of transition proportions $\mathbf{P}$ for healthy males and males with OSA (AHI = 5, 15, 30) at different ages (30, 50, 70 years). Estimates are supplemented with 95% bootstrapped confidence intervals.

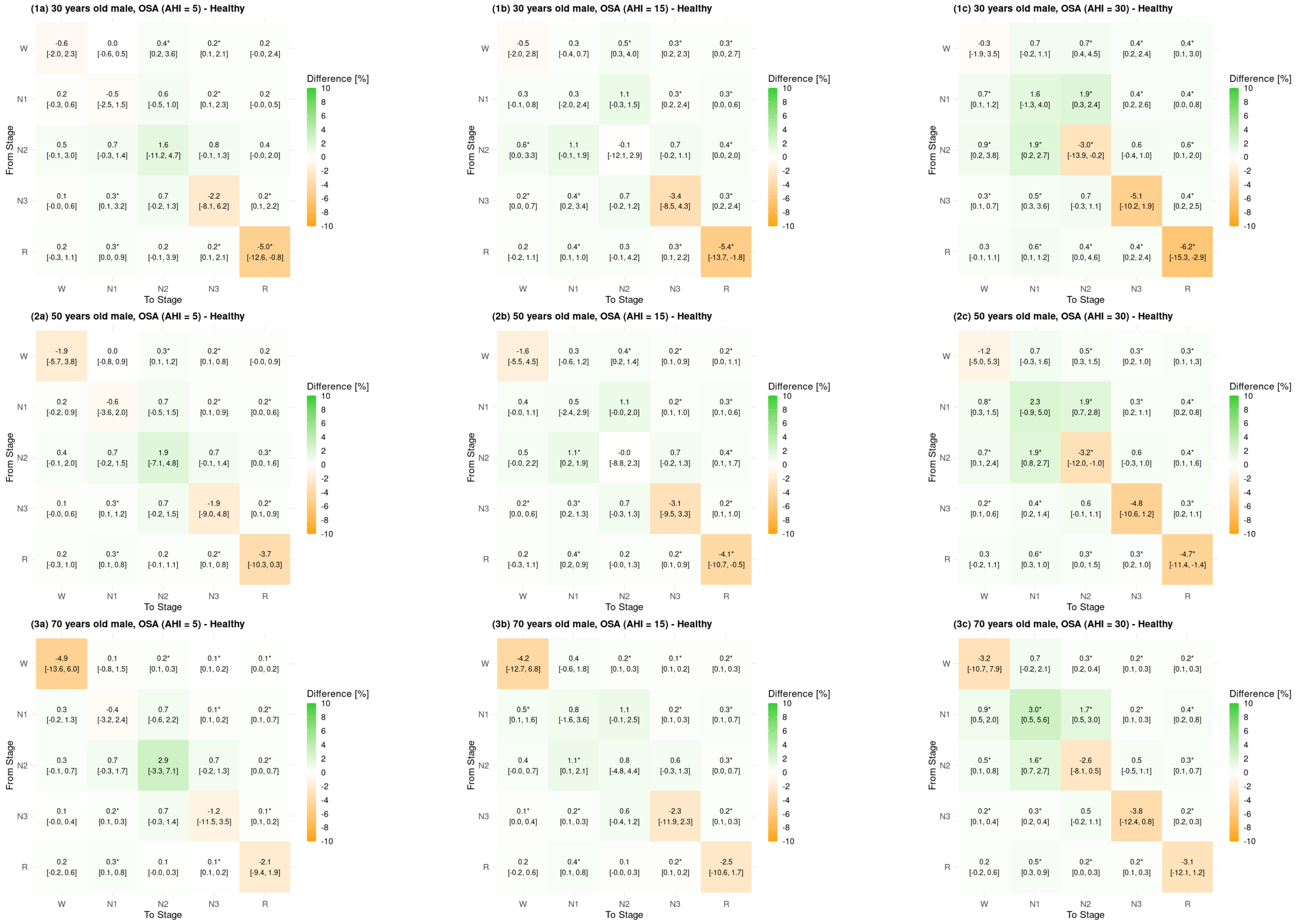


Figure 5: Difference (CATE) of matrices of transition proportions $\mathbf{P}$ for healthy males versus males with OSA (AHI = 5, 15, 30) at different ages (30, 50, 70 years). Estimates are supplemented with 95% bootstrapped confidence intervals.

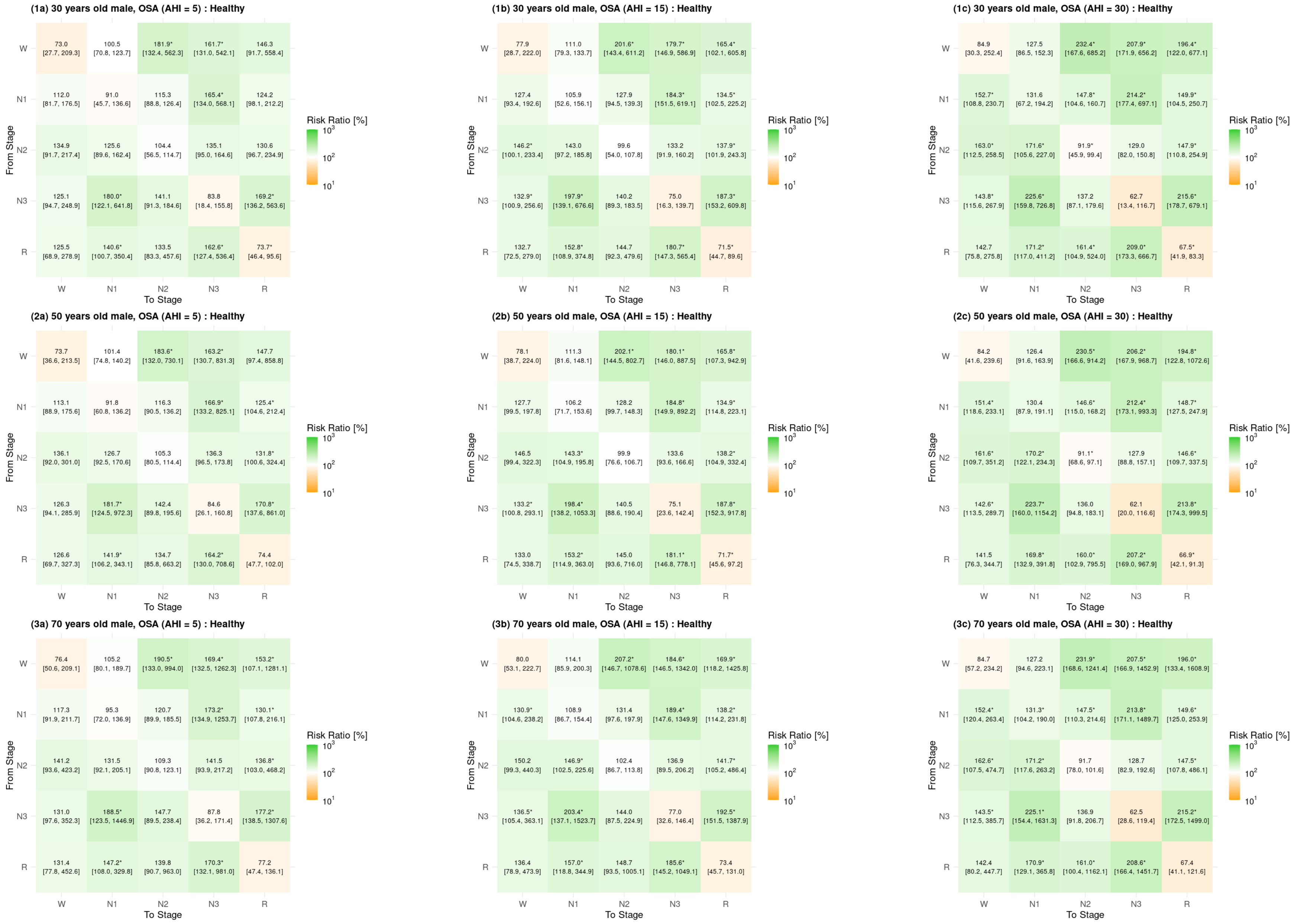


Figure 6: Risk ratio (RR-CATE) of matrices of transition proportions $\mathbf{P}$ for healthy males versus males with OSA (AHI = 5, 15, 30) at different ages (30, 50, 70 years). Estimates are supplemented with 95% bootstrapped confidence intervals.

#### Effect tables

| Quantity | Estimate | Healthy | O1: OSA (AHI = 5) | O2: OSA (AHI = 15) | O3: OSA (AHI = 30) |
|---|---|---|---|---|---|
| $P$(W) | % | 4.17 (2.85, 7.15) | 6.28 (4.45, 11.24) | 6.85 (4.75, 11.67) | 7.74 (5.34, 12.55) |
| | CATE | | **2.1*** (0.87, 4.29) | **2.68*** (1.42, 4.94) | **3.57*** (2.11, 5.91) |
| | RR-CATE | | **150.47*** (118.71, 199.74) | **164.35*** (129.93, 212.12) | **185.63*** (143.44, 236.11) |
| $P$(N1) | % | 8.93 (7.14, 11.04) | 10.52 (8.87, 12.22) | 12 (10.17, 13.57) | 14.45 (11.34, 16.46) |
| | CATE | | 1.59 (-0.52, 3.74) | **3.07*** (0.78, 5.23) | **5.53*** (2.8, 7.61) |
| | RR-CATE | | 117.87 (94.82, 146.85) | **134.41*** (107.51, 164.96) | **161.94*** (125.03, 197.44) |
| $P$(N2) | % | 43.03 (35.97, 46.7) | 45.99 (35.57, 49.56) | 45.29 (34.95, 48.5) | 43.98 (34.44, 47.83) |
| | CATE | | 2.95 (-2.47, 7.24) | 2.26 (-3.24, 6.9) | 0.94 (-4.71, 5.28) |
| | RR-CATE | | 106.86 (94.35, 118.57) | 105.25 (92.97, 117.03) | 102.2 (89.66, 113.11) |
| $P$(N3) | % | 22.45 (7.76, 29.61) | 21.02 (11.32, 28.36) | 19.62 (11.16, 26.19) | 17.65 (11.24, 23.86) |
| | CATE | | -1.43 (-6.72, 5.62) | -2.83 (-7.76, 5.96) | -4.8 (-9.49, 6.39) |
| | RR-CATE | | 93.63 (75.77, 164.23) | 87.4 (71.79, 165.62) | 78.61 (63.55, 171.61) |
| $P$(REM) | % | 21.42 (15.78, 39.37) | 16.2 (11.82, 28.76) | 16.24 (11.7, 28.3) | 16.18 (11.55, 27.31) |
| | CATE | | **-5.22*** (-12.13, -1.35) | **-5.18*** (-12.89, -1.7) | **-5.24*** (-13.99, -1.88) |
| | RR-CATE | | **75.62*** (63.08, 93.18) | **75.8*** (63.54, 91.42) | **75.53*** (62.73, 90.04) |
| $P$((N1,N2,N3,REM) $\to$ W) | % | 3.12 (2.2, 6.87) | 4.82 (3.24, 10.16) | 5.29 (3.6, 10.48) | 6.02 (4.04, 11.3) |
| | CATE | | **1.69*** (0.77, 3.64) | **2.16*** (1.12, 3.83) | **2.89*** (1.6, 4.53) |
| | RR-CATE | | **154.24*** (126.08, 195.75) | **169.25*** (135.59, 209.54) | **192.55*** (151.32, 230.47) |
| $P$((N1,N2) $\to$ W) | % | 2.01 (1.35, 4.25) | 3.11 (2.1, 7.13) | 3.46 (2.36, 7.37) | 4.02 (2.72, 7.83) |
| | CATE | | **1.1*** (0.46, 2.38) | **1.45*** (0.74, 2.88) | **2.01*** (1.18, 3.49) |
| | RR-CATE | | **154.75*** (121.62, 200.22) | **172.33*** (137.19, 215.64) | **200.35*** (154.77, 253.65) |
| $P$(N3 $\to$ W) | % | 0.59 (0.45, 1.09) | 0.84 (0.61, 1.46) | 0.89 (0.66, 1.49) | 0.98 (0.72, 1.56) |
| | CATE | | **0.25*** (0.03, 0.53) | **0.3*** (0.1, 0.58) | **0.39*** (0.17, 0.69) |
| | RR-CATE | | **141.77*** (104.75, 190.72) | **151.7*** (113.57, 199.84) | **165.96*** (124.65, 219.4) |
| $P$(REM $\to$ W) | % | 0.53 (0.25, 1.21) | 0.88 (0.46, 2.05) | 0.93 (0.5, 2.09) | 1.02 (0.51, 2.12) |
| | CATE | | **0.35*** (0.08, 1.1) | **0.41*** (0.12, 0.93) | **0.49*** (0.16, 0.96) |
| | RR-CATE | | **166.23*** (115.25, 259.61) | **177.15*** (122.92, 270.8) | **192.6*** (130.02, 293.61) |
| $P$(NREM $\rightleftarrows$ REM) | % | 3.19 (2.19, 7.53) | 5.53 (4.07, 15.36) | 6.02 (4.39, 16.38) | 6.78 (4.98, 17.83) |
| | CATE | | **2.33*** (1.19, 7.93) | **2.83*** (1.56, 8.93) | **3.59*** (2.06, 10.38) |
| | RR-CATE | | **173.18*** (135.99, 244.33) | **188.79*** (145.45, 260.22) | **212.48*** (159.99, 292.82) |
| $P$(N1 $\rightleftarrows$ N2) | % | 5.89 (4.25, 8.34) | 7.17 (5.41, 9.46) | 8.1 (6.07, 9.92) | 9.61 (7.14, 11.37) |
| | CATE | | 1.28 (-0.2, 2.42) | **2.21*** (0.59, 3.37) | **3.72*** (1.57, 5.04) |
| | RR-CATE | | 121.73 (97.19, 150.33) | **137.52*** (108.78, 167.48) | **163.24*** (125.05, 193.52) |
| $P$(Sleep compactness) | % | 92.82 (85.85, 95.05) | 89.31 (76.67, 92.32) | 88.21 (75.49, 91.52) | 86.49 (73.31, 90.45) |
| | CATE | | **-3.52*** (-8.83, -1.43) | **-4.62*** (-10.46, -2.37) | **-6.34*** (-12.18, -4.02) |
| | RR-CATE | | **96.21*** (89.76, 98.45) | **95.02*** (87.88, 97.4) | **93.17*** (85.11, 95.75) |
| $P$(Sleep fragmentation) | % | 6.13 (4.26, 13.87) | 9.24 (6.28, 23.18) | 10.23 (7.14, 24.35) | 11.79 (8.03, 26.05) |
| | CATE | | **3.11*** (1.43, 9.04) | **4.1*** (2.22, 10.58) | **5.66*** (3.38, 11.96) |
| | RR-CATE | | **150.7*** (124.39, 190.2) | **166.87*** (139.17, 204.44) | **192.33*** (160.07, 236.24) |
| $P$(Sleep-stage compactness) | % | 78.91 (61.89, 82.74) | 68.93 (36.64, 74.62) | 66.28 (34.47, 72.21) | 62.15 (30.04, 68.79) |
| | CATE | | **-9.98*** (-25.15, -6.58) | **-12.64*** (-28.1, -9.02) | **-16.76*** (-32.37, -12.66) |
| | RR-CATE | | **87.35*** (58.8, 91.84) | **83.98*** (55.19, 88.7) | **78.76*** (49.17, 84.41) |
| $P$(Sleep-stage fragmentation) | % | 13.91 (10.74, 23.49) | 20.37 (16, 40.05) | 21.93 (17.32, 40.28) | 24.33 (18.83, 42.09) |
| | CATE | | **6.46*** (3.09, 16.83) | **8.02*** (4.56, 17.45) | **10.42*** (6.5, 18.44) |
| | RR-CATE | | **146.46*** (124.33, 187.22) | **157.66*** (134, 198.38) | **174.94*** (148.28, 216.31) |
| $P$(W $\to$ W) | % | 1.05 (0.33, 2.92) | 1.46 (0.18, 4.27) | 1.57 (0.19, 4.64) | 1.73 (0.17, 5.32) |
| | CATE | | 0.41 (-0.35, 2.23) | 0.52 (-0.28, 2.52) | 0.68 (-0.21, 2.97) |
| | RR-CATE | | 139.22 (42.54, 274.01) | 149.7 (44.75, 299.99) | 164.95 (45.63, 333.06) |
| $P$(N1 $\to$ N1) | % | 4.01 (1.59, 5.26) | 3.53 (0.77, 4.91) | 4.14 (0.86, 5.73) | 5.2 (1.03, 7.1) |
| | CATE | | -0.48 (-1.85, 0.73) | 0.13 (-1.44, 1.44) | 1.19 (-1.11, 2.86) |
| | RR-CATE | | 88.06 (37.68, 118.66) | 103.3 (43.8, 136.66) | 129.7 (54.59, 171.73) |
| $P$(N2 $\to$ N2) | % | 36.34 (24.17, 40.75) | 36.85 (16.05, 41.1) | 35.44 (14.74, 39.78) | 33.03 (13.14, 38) |
| | CATE | | 0.5 (-6.46, 5.55) | -0.9 (-8.42, 4.06) | -3.31 (-10.99, 1.39) |
| | RR-CATE | | 101.39 (68.82, 115.48) | 97.51 (62.45, 110.85) | 90.89 (54.45, 104.41) |
| $P$(N3 $\to$ N3) | % | 19.26 (1.59, 26.61) | 15.9 (0.87, 23.85) | 14.33 (0.78, 21.61) | 12.11 (0.68, 18.55) |
| | CATE | | -3.36 (-8.53, 0.35) | **-4.93*** (-9.66, -0.44) | **-7.15*** (-11.67, -0.76) |
| | RR-CATE | | 82.55 (43.2, 104.4) | **74.38*** (40.56, 91.4) | **62.87*** (29.6, 77.92) |
| $P$(REM $\to$ REM) | % | 19.3 (13.72, 34.14) | 12.66 (8.28, 20.06) | 12.37 (8.32, 19.29) | 11.81 (7.62, 18.04) |
| | CATE | | **-6.65*** (-16.35, -2.47) | **-6.93*** (-17.11, -2.97) | **-7.49*** (-18.28, -3.86) |
| | RR-CATE | | **65.57*** (49.07, 83.92) | **64.08*** (46.32, 81.8) | **61.18*** (42.31, 77.66) |
| $P$(W-fragmentation) | % | 3.01 (2.09, 7.01) | 4.42 (3.06, 12.3) | 4.94 (3.51, 13.03) | 5.77 (4, 14.79) |
| | CATE | | **1.41*** (0.65, 5.38) | **1.94*** (1.05, 6.47) | **2.77*** (1.74, 7.97) |
| | RR-CATE | | **147.02*** (125.01, 186.42) | **164.39*** (139.85, 201.78) | **192.1*** (163.44, 238.83) |
| $P$(N1-fragmentation) | % | 5.16 (3.94, 7.31) | 7.02 (5.7, 9.56) | 7.86 (6.47, 10.26) | 9.21 (7.52, 11.44) |
| | CATE | | **1.86*** (0.96, 2.8) | **2.71*** (1.65, 3.78) | **4.05*** (2.86, 5.17) |
| | RR-CATE | | **136.16*** (115.95, 162.61) | **152.47*** (130.35, 178.28) | **178.61*** (143.53, 209.53) |
| $P$(N2-fragmentation) | % | 6.78 (5.13, 12.06) | 9.65 (7.38, 18.19) | 10.31 (7.8, 18.48) | 11.33 (8.47, 18.81) |
| | CATE | | **2.86*** (1.26, 5.9) | **3.53*** (1.81, 5.97) | **4.55*** (2.69, 6.7) |
| | RR-CATE | | **142.19*** (118.74, 170.61) | **151.99*** (127.54, 182.91) | **167.08*** (139.25, 199.61) |
| $P$(N3-fragmentation) | % | 3.15 (2.3, 6.58) | 5.05 (3.71, 12.17) | 5.26 (3.92, 12.5) | 5.54 (4.11, 12.97) |
| | CATE | | **1.9*** (0.77, 6.35) | **2.11*** (1.02, 6.56) | **2.39*** (1.18, 6.62) |
| | RR-CATE | | **160.31*** (123.48, 224.18) | **166.87*** (132.04, 230.58) | **175.86*** (138.11, 238.99) |
| $P$(REM-fragmentation) | % | 1.94 (1.21, 4.89) | 3.47 (2.45, 10.09) | 3.79 (2.7, 10.78) | 4.26 (3.06, 11.58) |
| | CATE | | **1.53*** (0.75, 5.22) | **1.84*** (0.97, 5.95) | **2.32*** (1.29, 7) |
| | RR-CATE | | **178.76*** (134.92, 258.28) | **194.95*** (147.82, 280.14) | **219.51*** (165.81, 312.96) |

Table 2: Summary of expected probabilities (%) and estimated effects of OSA (CATE, RR-CATE) for 30-year-old females.

| Quantity | Estimate | Healthy | O1: OSA (AHI = 5) | O2: OSA (AHI = 15) | O3: OSA (AHI = 30) |
|---|---|---|---|---|---|
| $P$(W) | % | 6.59 (4.83, 9.19) | 9.51 (7.29, 12.83) | 10.3 (8.11, 13.63) | 11.46 (9.19, 14.72) |
| | CATE | | **2.92*** (0.29, 6.05) | **3.7*** (1.11, 6.77) | **4.87*** (2.14, 7.86) |
| | RR-CATE | | **144.3*** (104.48, 208.14) | **156.17*** (114.78, 221.8) | **173.85*** (126.1, 244.19) |
| $P$(N1) | % | 11.2 (9.66, 13.59) | 12.33 (10.63, 13.97) | 14.04 (12.07, 15.91) | 16.85 (14.28, 18.41) |
| | CATE | | 1.13 (-1.63, 3.67) | 2.83 (-0.11, 5.36) | **5.65*** (2.13, 7.98) |
| | RR-CATE | | 110.1 (86.24, 135.03) | 125.29 (99.12, 151.07) | **150.4*** (117.31, 180.77) |
| $P$(N2) | % | 43.99 (40.42, 48.3) | 45.87 (43.03, 49) | 44.83 (42.38, 47.65) | 43 (40.43, 45.75) |
| | CATE | | 1.88 (-3.03, 6.14) | 0.85 (-4.33, 5.13) | -0.99 (-6.38, 3.08) |
| | RR-CATE | | 104.28 (93.63, 115.16) | 101.93 (91, 112.67) | 97.75 (86.82, 107.5) |
| $P$(N3) | % | 21.29 (11.26, 24.02) | 19.49 (12.84, 22.54) | 18.05 (12.38, 20.5) | 16.02 (11.57, 18.25) |
| | CATE | | -1.81 (-5.97, 3.33) | -3.25 (-7.51, 2.15) | -5.28 (-9.29, 1.1) |
| | RR-CATE | | 91.5 (72.53, 128.84) | 84.75 (68.01, 123.55) | 75.22 (60.11, 112.02) |
| $P$(REM) | % | 16.92 (15.51, 22.66) | 12.8 (11.05, 18.45) | 12.79 (11.08, 18.39) | 12.67 (11.04, 18.38) |
| | CATE | | **-4.13*** (-7.41, -1.08) | **-4.14*** (-7.11, -1.2) | **-4.25*** (-7.32, -1.23) |
| | RR-CATE | | **75.61*** (63.48, 93.29) | **75.56*** (64.48, 92.93) | **74.9*** (63.2, 92.36) |
| $P$((N1,N2,N3,REM) $\to$ W) | % | 3.08 (2.66, 4.38) | 4.71 (4.06, 7.35) | 5.16 (4.48, 7.77) | 5.86 (5.02, 8.6) |
| | CATE | | **1.63*** (0.91, 3.19) | **2.08*** (1.34, 3.81) | **2.78*** (1.94, 4.76) |
| | RR-CATE | | **153.03*** (129.36, 197.2) | **167.68*** (141.27, 208.08) | **190.25*** (158.06, 239.26) |
| $P$((N1,N2) $\to$ W) | % | 2.05 (1.65, 2.95) | 3.16 (2.72, 4.69) | 3.52 (3.05, 5.1) | 4.08 (3.47, 5.9) |
| | CATE | | **1.11*** (0.59, 1.99) | **1.46*** (0.95, 2.46) | **2.03*** (1.44, 3.24) |
| | RR-CATE | | **154.05*** (125.49, 200.21) | **171.38*** (141.51, 224.97) | **198.82*** (162.16, 251.85) |
| $P$(N3 $\to$ W) | % | 0.52 (0.42, 0.77) | 0.72 (0.57, 1.09) | 0.77 (0.61, 1.12) | 0.83 (0.67, 1.15) |
| | CATE | | **0.2*** (0.03, 0.46) | **0.25*** (0.07, 0.49) | **0.31*** (0.14, 0.54) |
| | RR-CATE | | **139.15*** (105.33, 191.21) | **148.04*** (113.09, 200.2) | **160.43*** (122.7, 213.34) |
| $P$(REM $\to$ W) | % | 0.51 (0.33, 0.79) | 0.83 (0.66, 1.34) | 0.88 (0.7, 1.4) | 0.94 (0.74, 1.52) |
| | CATE | | **0.32*** (0.09, 0.78) | **0.37*** (0.13, 0.77) | **0.44*** (0.17, 0.84) |
| | RR-CATE | | **163.17*** (114.04, 284.16) | **172.87*** (121.72, 302.79) | **186.19*** (128.28, 307.05) |
| $P$(NREM $\rightleftarrows$ REM) | % | 2.75 (2.07, 3.73) | 4.77 (3.96, 7.34) | 5.17 (4.34, 8) | 5.76 (4.85, 8.8) |
| | CATE | | **2.01*** (1.08, 4.42) | **2.41*** (1.34, 5.09) | **3.01*** (1.78, 6.07) |
| | RR-CATE | | **173.2*** (133.27, 253.24) | **187.76*** (140.87, 274.42) | **209.38*** (153.92, 308.82) |
| $P$(N1 $\rightleftarrows$ N2) | % | 6.06 (5.12, 7.63) | 7.24 (6.43, 9.46) | 8.13 (7.32, 10.42) | 9.56 (8.73, 11.75) |
| | CATE | | 1.18 (-0.08, 2.65) | **2.07*** (0.77, 3.61) | **3.5*** (2.11, 5.21) |
| | RR-CATE | | 119.5 (98.73, 150.25) | **134.22*** (111.38, 169.78) | **157.84*** (130.84, 192.88) |
| $P$(Sleep compactness) | % | 90.39 (87.53, 92.32) | 86.29 (81.86, 88.51) | 85.04 (80.68, 87.24) | 83.15 (78.95, 85.5) |
| | CATE | | **-4.1*** (-8.04, -1.22) | **-5.35*** (-8.92, -2.57) | **-7.24*** (-11.29, -4.49) |
| | RR-CATE | | **95.46*** (91.07, 98.63) | **94.08*** (89.48, 97.11) | **91.99*** (87.49, 94.94) |
| $P$(Sleep fragmentation) | % | 6.09 (5.31, 8.39) | 8.91 (7.69, 14.29) | 9.82 (8.49, 15.36) | 11.25 (9.61, 16.91) |
| | CATE | | **2.82*** (1.55, 6.03) | **3.73*** (2.34, 7.29) | **5.16*** (3.61, 9.23) |
| | RR-CATE | | **146.23*** (124.43, 185.01) | **161.26*** (136.57, 198.55) | **184.62*** (155.03, 230.8) |
| $P$(Sleep-stage compactness) | % | 76.97 (71.41, 78.93) | 67.13 (53.56, 70.5) | 64.53 (51.15, 67.77) | 60.6 (47.5, 63.3) |
| | CATE | | **-9.85*** (-19.52, -7.1) | **-12.44*** (-21.56, -9.78) | **-16.38*** (-24.97, -13.75) |
| | RR-CATE | | **87.21*** (73.02, 90.71) | **83.84*** (70.12, 87.17) | **78.73*** (65.58, 82.08) |
| $P$(Sleep-stage fragmentation) | % | 13.42 (11.04, 17.5) | 19.16 (17.2, 29.1) | 20.51 (18.28, 29.68) | 22.55 (20.13, 31.66) |
| | CATE | | **5.74*** (2.85, 14.85) | **7.09*** (3.97, 15.67) | **9.13*** (5.63, 17.11) |
| | RR-CATE | | **142.78*** (120.55, 198.49) | **152.82*** (127.93, 205.73) | **168.03*** (139.01, 219.11) |
| $P$(W $\to$ W) | % | 3.51 (1.82, 6.07) | 4.8 (1.68, 7.56) | 5.13 (1.75, 8.07) | 5.6 (1.92, 8.91) |
| | CATE | | 1.29 (-1.47, 4.12) | 1.62 (-1.28, 4.51) | 2.09 (-0.93, 5.24) |
| | RR-CATE | | 136.65 (48.5, 264.53) | 146.08 (53.57, 288.02) | 159.46 (56.6, 309.79) |
| $P$(N1 $\to$ N1) | % | 6.02 (4.46, 7.56) | 5.2 (2.34, 6.61) | 6.07 (2.68, 7.5) | 7.55 (3.27, 9.26) |
| | CATE | | -0.82 (-3.36, 0.8) | 0.05 (-2.87, 1.72) | 1.53 (-1.97, 3.47) |
| | RR-CATE | | 86.44 (43.49, 114.53) | 100.81 (51.32, 131.2) | 125.39 (65.66, 165.26) |
| $P$(N2 $\to$ N2) | % | 37.55 (33.15, 41.94) | 37.37 (32.08, 39.99) | 35.73 (30.25, 37.95) | 32.99 (27.23, 35.42) |
| | CATE | | -0.18 (-7.32, 4.63) | -1.82 (-9.5, 3.01) | -4.55 (-12.92, 0.09) |
| | RR-CATE | | 99.52 (82.57, 113.95) | 95.16 (75.88, 108.8) | 87.87 (67.8, 100.27) |
| $P$(N3 $\to$ N3) | % | 18.34 (7.43, 20.79) | 14.86 (5.7, 17.68) | 13.31 (4.96, 15.55) | 11.15 (4.19, 13.2) |
| | CATE | | -3.48 (-7.51, 0.55) | **-5.03*** (-9.05, -1.16) | **-7.19*** (-10.64, -2.35) |
| | RR-CATE | | 81.03 (49.63, 105.85) | **72.59*** (44.01, 91.8) | **60.77*** (39.8, 76.63) |
| $P$(REM $\to$ REM) | % | 15.07 (13.69, 20.4) | 9.7 (8.03, 14.05) | 9.42 (7.99, 13.8) | 8.91 (7.28, 13.01) |
| | CATE | | **-5.37*** (-9.41, -2.3) | **-5.64*** (-9.83, -2.68) | **-6.15*** (-10.56, -3.05) |
| | RR-CATE | | **64.36*** (50.52, 84.26) | **62.54*** (50.52, 81.8) | **59.15*** (45.33, 79.15) |
| $P$(W-fragmentation) | % | 3.01 (2.62, 4) | 4.2 (3.55, 6.48) | 4.66 (4.02, 7.11) | 5.39 (4.59, 8.52) |
| | CATE | | **1.18*** (0.6, 2.7) | **1.65*** (0.99, 3.31) | **2.38*** (1.65, 4.44) |
| | RR-CATE | | **139.27*** (119.02, 173.29) | **154.69*** (133.34, 190.31) | **178.86*** (152.82, 225.96) |
| $P$(N1-fragmentation) | % | 5.45 (4.72, 6.8) | 7.34 (6.58, 9.43) | 8.18 (7.48, 10.29) | 9.5 (8.7, 11.5) |
| | CATE | | **1.89*** (0.88, 3.13) | **2.72*** (1.8, 3.98) | **4.05*** (3, 5.45) |
| | RR-CATE | | **134.59*** (115.31, 159.43) | **149.96*** (131.13, 174.85) | **174.24*** (149.31, 198.76) |
| $P$(N2-fragmentation) | % | 6.44 (5.32, 8.59) | 8.97 (8, 14.33) | 9.54 (8.51, 14.65) | 10.41 (9.35, 15.37) |
| | CATE | | **2.53*** (1.1, 6.61) | **3.1*** (1.66, 7.07) | **3.97*** (2.54, 7.94) |
| | RR-CATE | | **139.3*** (116.03, 183.45) | **148.19*** (121.75, 191.06) | **161.68*** (135.07, 202.4) |
| $P$(N3-fragmentation) | % | 2.93 (2.23, 4.29) | 4.59 (3.88, 7.89) | 4.73 (3.99, 8.03) | 4.9 (4.15, 8.33) |
| | CATE | | **1.66*** (0.59, 4.89) | **1.8*** (0.79, 4.81) | **1.97*** (0.93, 4.67) |
| | RR-CATE | | **156.56*** (117.24, 230.22) | **161.32*** (122.44, 232.53) | **167.29*** (128.36, 238.25) |
| $P$(REM-fragmentation) | % | 1.68 (1.19, 2.37) | 2.98 (2.46, 4.7) | 3.23 (2.72, 5.02) | 3.6 (3, 5.37) |
| | CATE | | **1.3*** (0.64, 3.01) | **1.55*** (0.84, 3.28) | **1.92*** (1.14, 3.67) |
| | RR-CATE | | **177.4*** (131.47, 280.3) | **192.2*** (144.46, 301.09) | **214.16*** (157.16, 324.83) |

Table 3: Summary of expected probabilities (%) and estimated effects of OSA (CATE, RR-CATE) for 50-year-old females.

| Quantity | Estimate | Healthy | O1: OSA (AHI = 5) | O2: OSA (AHI = 15) | O3: OSA (AHI = 30) |
|---|---|---|---|---|---|
| $P$(W) | % | 13.8 (7.73, 20.64) | 18.74 (13.44, 22.39) | 19.94 (14.68, 23.54) | 21.63 (16.57, 24.99) |
| | CATE | | 4.94 (-4.07, 11.5) | 6.14 (-2.46, 12.33) | 7.83 (-0.68, 13.55) |
| | RR-CATE | | 135.8 (79.4, 225.94) | 144.51 (87.08, 242.92) | 156.75 (96.45, 267.43) |
| $P$(N1) | % | 13.45 (10.83, 16.5) | 13.68 (10.24, 16.48) | 15.45 (11.91, 18.02) | 18.33 (14.32, 20.73) |
| | CATE | | 0.23 (-3.09, 3.34) | 2 (-1.78, 5.02) | **4.87*** (1.03, 8.02) |
| | RR-CATE | | 101.7 (76.75, 127.51) | 114.84 (89.14, 140.83) | **136.23*** (105.89, 166.41) |
| $P$(N2) | % | 41.67 (34.75, 46.54) | 41.83 (37.94, 46.19) | 40.36 (36.81, 44.91) | 37.96 (34.94, 42.21) |
| | CATE | | 0.16 (-4.67, 7.61) | -1.3 (-6.17, 5.71) | -3.71 (-8.8, 4.44) |
| | RR-CATE | | 100.39 (89.51, 121.1) | 96.87 (86.38, 116.94) | 91.1 (80.64, 111.68) |
| $P$(N3) | % | 18.69 (11.54, 34.87) | 16.47 (11.19, 25.59) | 15.06 (10.31, 24.49) | 13.09 (9.34, 20.67) |
| | CATE | | -2.21 (-11.12, 2.79) | -3.63 (-12.84, 0.61) | **-5.6*** (-15.5, -0.66) |
| | RR-CATE | | 88.16 (64.41, 115.1) | 80.57 (61.66, 102.97) | **70.04*** (52.99, 93.94) |
| $P$(REM) | % | 12.39 (6.86, 18.64) | 9.27 (6.49, 13.55) | 9.19 (6.58, 13.02) | 8.99 (6.53, 12.45) |
| | CATE | | -3.12 (-6.41, 0.65) | -3.21 (-6.65, 0.8) | -3.4 (-7.42, 0.65) |
| | RR-CATE | | 74.81 (61.93, 108.95) | 74.13 (61.67, 110.74) | 72.57 (58.98, 107.88) |
| $P$((N1,N2,N3,REM) $\rightarrow$ W) | % | 2.89 (1.76, 4.04) | 4.33 (3.19, 6.02) | 4.72 (3.59, 6.39) | 5.3 (4.15, 6.77) |
| | CATE | | **1.44*** (0.81, 2.59) | **1.82*** (1.22, 2.96) | **2.4*** (1.81, 3.52) |
| | RR-CATE | | **149.69*** (125.93, 205.77) | **162.94*** (136.15, 227.13) | **183.04*** (151.34, 257.14) |
| $P$((N1,N2) $\rightarrow$ W) | % | 2.02 (1.25, 2.84) | 3.05 (2.24, 4.08) | 3.37 (2.56, 4.45) | 3.87 (3.07, 4.82) |
| | CATE | | **1.03*** (0.54, 1.84) | **1.35*** (0.92, 2.15) | **1.85*** (1.4, 2.64) |
| | RR-CATE | | **151.07*** (122.94, 203.07) | **166.94*** (137.15, 228.97) | **191.67*** (155.42, 261.8) |
| $P$(N3 $\rightarrow$ W) | % | 0.42 (0.25, 0.55) | 0.57 (0.41, 0.74) | 0.6 (0.45, 0.78) | 0.64 (0.49, 0.81) |
| | CATE | | **0.15*** (0.02, 0.35) | **0.18*** (0.05, 0.37) | **0.22*** (0.09, 0.41) |
| | RR-CATE | | **134.54*** (103.84, 209.11) | **141.49*** (109.8, 218.61) | **150.68*** (116.73, 230.17) |
| $P$(REM $\rightarrow$ W) | % | 0.45 (0.19, 0.75) | 0.71 (0.47, 1.19) | 0.75 (0.49, 1.17) | 0.79 (0.54, 1.18) |
| | CATE | | **0.26*** (0.06, 0.53) | **0.29*** (0.09, 0.56) | **0.34*** (0.12, 0.6) |
| | RR-CATE | | **157.76*** (110.52, 334.5) | **165.22*** (115.34, 349.12) | **174.87*** (120.53, 385.77) |
| $P$(NREM $\rightleftarrows$ REM) | % | 2.24 (1.27, 3.46) | 3.82 (2.8, 5.02) | 4.09 (3.05, 5.21) | 4.49 (3.3, 5.38) |
| | CATE | | **1.58*** (0.85, 2.52) | **1.86*** (1.02, 2.76) | **2.25*** (1.34, 3.09) |
| | RR-CATE | | **170.81*** (129.14, 284.15) | **183.08*** (135.16, 303.51) | **200.68*** (143.42, 330.32) |
| $P$(N1 $\rightleftarrows$ N2) | % | 5.76 (3.82, 7.61) | 6.66 (5.39, 8.42) | 7.39 (6.04, 9.22) | 8.54 (7.14, 10.54) |
| | CATE | | 0.9 (-0.41, 2.98) | **1.63*** (0.37, 3.7) | **2.78*** (1.51, 4.79) |
| | RR-CATE | | 115.55 (94.17, 178) | **128.3*** (105.39, 196.79) | **148.27*** (122.41, 225.89) |
| $P$(Sleep compactness) | % | 83.3 (76.47, 89.69) | 77.47 (72.87, 83.36) | 75.9 (71.72, 81.81) | 73.65 (70.14, 79.12) |
| | CATE | | -5.83 (-12.03, 3.03) | -7.4 (-13.69, 1.33) | **-9.65*** (-15.89, -0.76) |
| | RR-CATE | | 93 (86.19, 103.88) | 91.12 (84.42, 101.74) | **88.42*** (81.88, 99.03) |
| $P$(Sleep fragmentation) | % | 5.79 (3.58, 7.86) | 8.12 (5.96, 11.21) | 8.87 (6.72, 11.91) | 10.01 (7.78, 12.72) |
| | CATE | | **2.32*** (1.22, 4.35) | **3.08*** (2.01, 4.97) | **4.22*** (3.16, 6.22) |
| | RR-CATE | | **140.11*** (118.84, 184.86) | **153.08*** (129.66, 204.25) | **172.79*** (144.49, 237.49) |
| $P$(Sleep-stage compactness) | % | 71.21 (66.28, 79.03) | 60.88 (54.79, 68.14) | 58.35 (53.03, 65.4) | 54.67 (50.01, 62.23) |
| | CATE | | **-10.33*** (-15.96, -3.3) | **-12.87*** (-18.33, -6.4) | **-16.54*** (-21.92, -9.7) |
| | RR-CATE | | **85.5*** (78.04, 94.81) | **81.93*** (75.07, 90.62) | **76.77*** (70.92, 86.32) |
| $P$(Sleep-stage fragmentation) | % | 12.09 (7.7, 16.15) | 16.59 (13.43, 21.14) | 17.56 (14.4, 21.82) | 18.98 (15.98, 22.68) |
| | CATE | | **4.5*** (1.81, 9.37) | **5.47*** (2.72, 10.01) | **6.89*** (3.7, 11.3) |
| | RR-CATE | | **137.23*** (112.42, 230.9) | **145.24*** (117.68, 234.03) | **157.03*** (126, 255.23) |
| $P$(W $\rightarrow$ W) | % | 10.91 (4.79, 18.51) | 14.41 (9.29, 17.2) | 15.23 (10.04, 18.13) | 16.33 (11.23, 19.13) |
| | CATE | | 3.5 (-5.93, 9.37) | 4.32 (-5.09, 9.86) | 5.43 (-3.51, 10.84) |
| | RR-CATE | | 132.12 (60.79, 257.6) | 139.62 (67.22, 272.05) | 149.77 (74.8, 292.61) |
| $P$(N1 $\rightarrow$ N1) | % | 8.35 (6.57, 10.76) | 6.98 (4.45, 8.43) | 8.05 (5.22, 9.61) | 9.83 (6.55, 11.54) |
| | CATE | | -1.37 (-4.15, 0.72) | -0.31 (-3.22, 1.84) | 1.48 (-1.84, 3.87) |
| | RR-CATE | | 83.57 (49.73, 110.6) | 96.35 (60.43, 125.95) | 117.76 (79.01, 155.24) |
| $P$(N2 $\rightarrow$ N2) | % | 35.85 (29.77, 40.35) | 34.5 (30.6, 38.71) | 32.61 (29.19, 36.5) | 29.59 (26.58, 33.06) |
| | CATE | | -1.36 (-6.04, 4.57) | -3.25 (-7.91, 2.61) | -6.26 (-11.26, 0.82) |
| | RR-CATE | | 96.22 (83.82, 114.11) | 90.95 (79.1, 108.59) | 82.53 (70.26, 102.74) |
| $P$(N3 $\rightarrow$ N3) | % | 16.14 (9.49, 33.3) | 12.64 (6.87, 22.38) | 11.2 (6.08, 21.43) | 9.21 (5.23, 17.07) |
| | CATE | | -3.5 (-12.45, 1.11) | **-4.94*** (-13.98, -1.32) | **-6.93*** (-16.72, -2.62) |
| | RR-CATE | | 78.34 (56.09, 106.72) | **69.37*** (50.76, 91.53) | **57.08*** (42.04, 78.53) |
| $P$(REM $\rightarrow$ REM) | % | 10.87 (5.9, 17.05) | 6.76 (4.53, 10.91) | 6.49 (4.37, 10.14) | 6.04 (4.24, 9.38) |
| | CATE | | **-4.1*** (-7.73, -0.55) | **-4.37*** (-8.19, -0.58) | **-4.83*** (-8.96, -0.85) |
| | RR-CATE | | **62.23*** (49.83, 91.26) | **59.77*** (48.03, 93.54) | **55.55*** (42.97, 87.33) |
| $P$(W-fragmentation) | % | 2.9 (1.82, 3.94) | 3.79 (2.77, 5.17) | 4.15 (3.13, 5.4) | 4.71 (3.68, 5.94) |
| | CATE | | **0.89*** (0.36, 1.78) | **1.25*** (0.75, 2.08) | **1.81*** (1.34, 2.74) |
| | RR-CATE | | **130.55*** (110.34, 171.89) | **143.23*** (122.03, 184.49) | **162.55*** (138.65, 218.91) |
| $P$(N1-fragmentation) | % | 5.39 (3.84, 6.86) | 7.08 (5.69, 8.61) | 7.8 (6.51, 9.36) | 8.92 (7.62, 10.35) |
| | CATE | | **1.69*** (0.72, 3.21) | **2.41*** (1.45, 3.95) | **3.53*** (2.56, 5.23) |
| | RR-CATE | | **131.36*** (111.36, 185.69) | **144.83*** (122.94, 205.3) | **165.62*** (139.92, 231.63) |
| $P$(N2-fragmentation) | % | 5.68 (3.42, 7.75) | 7.64 (6.11, 10) | 8.04 (6.58, 10.24) | 8.64 (7.19, 10.72) |
| | CATE | | **1.96*** (0.71, 4.31) | **2.36*** (1.12, 4.55) | **2.96*** (1.69, 5.05) |
| | RR-CATE | | **134.4*** (111.99, 229.52) | **141.51*** (114.46, 238.97) | **152.04*** (124.4, 253) |
| $P$(N3-fragmentation) | % | 2.54 (1.52, 3.79) | 3.83 (3.04, 5.18) | 3.88 (3.08, 5.09) | 3.93 (3.13, 4.93) |
| | CATE | | **1.29*** (0.3, 2.71) | **1.34*** (0.37, 2.65) | **1.39*** (0.45, 2.55) |
| | RR-CATE | | **150.72*** (109.06, 278.93) | **152.91*** (110.17, 275.26) | **154.84*** (112.07, 268.2) |
| $P$(REM-fragmentation) | % | 1.37 (0.74, 2.21) | 2.38 (1.73, 3.27) | 2.55 (1.9, 3.38) | 2.79 (2.02, 3.53) |
| | CATE | | **1.01*** (0.42, 1.62) | **1.17*** (0.55, 1.78) | **1.41*** (0.74, 2.01) |
| | RR-CATE | | **173.23*** (123.6, 301.1) | **185.4*** (131.69, 314.71) | **202.79*** (142.65, 332.9) |

Table 4: Summary of expected probabilities (%) and estimated effects of OSA (CATE, RR-CATE) for 70-year-old females.

| Quantity | Estimate | Healthy | O1: OSA (AHI = 5) | O2: OSA (AHI = 15) | O3: OSA (AHI = 30) |
|---|---|---|---|---|---|
| $P$(W) | % | 6.24 (4.41, 9.27) | 6.6 (4.59, 11.36) | 7.16 (4.95, 11.89) | 8.01 (5.61, 12.25) |
| | CATE | | 0.35 (-1.68, 4.55) | 0.92 (-1.05, 5.07) | 1.76 (-0.36, 6.27) |
| | RR-CATE | | 105.69 (76.98, 186.83) | 114.68 (85.13, 201.2) | 128.26 (94.34, 224.75) |
| $P$(N1) | % | 11.51 (8.83, 14) | 12.42 (10.39, 14.18) | 14.09 (11.59, 15.7) | 16.85 (13.59, 18.63) |
| | CATE | | 0.9 (-0.97, 3.82) | **2.57*** (0.66, 5.26) | **5.33*** (2.66, 8.02) |
| | RR-CATE | | 107.85 (91.92, 140.3) | **122.36*** (105.28, 159.89) | **146.31*** (120.47, 187.6) |
| $P$(N2) | % | 43.53 (35.5, 48.16) | 47.15 (31.96, 49.36) | 46.11 (31.57, 48.49) | 44.3 (31.25, 47.09) |
| | CATE | | 3.62 (-4.01, 6.34) | 2.57 (-4.99, 5.7) | 0.77 (-7.05, 3.88) |
| | RR-CATE | | 108.3 (90.39, 116.23) | 105.91 (87.91, 113.33) | 101.76 (84.46, 109.71) |
| $P$(N3) | % | 16.93 (5.67, 21.61) | 16.1 (9.13, 23.13) | 15.03 (8.94, 21.79) | 13.53 (8.7, 19.75) |
| | CATE | | -0.83 (-5.03, 7.79) | -1.91 (-6.21, 6.34) | -3.41 (-7.25, 5.82) |
| | RR-CATE | | 95.1 (72.79, 173.01) | 88.74 (68.08, 173.22) | 79.88 (62.77, 178.25) |
| $P$(REM) | % | 21.77 (17.6, 38.26) | 17.73 (12.45, 32.81) | 17.62 (12.51, 32.06) | 17.32 (12.07, 31.15) |
| | CATE | | -4.04 (-10.82, 1) | -4.16 (-10.92, 0.54) | **-4.46*** (-11.44, -0.25) |
| | RR-CATE | | 81.43 (57.64, 105.31) | 80.91 (57.82, 102.88) | **79.54*** (54.34, 98.67) |
| $P$((N1,N2,N3,REM) $\to$ W) | % | 3.97 (2.74, 5.53) | 4.94 (3.27, 9.76) | 5.39 (3.57, 10.25) | 6.08 (4.03, 10.95) |
| | CATE | | 0.97 (-0.16, 4.48) | **1.42*** (0.2, 5) | **2.11*** (0.66, 5.33) |
| | RR-CATE | | 124.42 (95.66, 197.26) | **135.75*** (105.9, 209.74) | **153.11*** (119.35, 219.6) |
| $P$((N1,N2) $\to$ W) | % | 2.65 (2.01, 4) | 3.28 (2.17, 7.33) | 3.63 (2.39, 7.66) | 4.18 (2.78, 7.96) |
| | CATE | | 0.64 (-0.14, 3.1) | **0.99*** (0.11, 3.55) | **1.54*** (0.47, 3.94) |
| | RR-CATE | | 123.99 (94.43, 189.08) | **137.23*** (105.04, 200) | **158.07*** (120.57, 219.84) |
| $P$(N3 $\to$ W) | % | 0.57 (0.3, 0.88) | 0.72 (0.52, 1.25) | 0.76 (0.56, 1.26) | 0.83 (0.59, 1.31) |
| | CATE | | 0.14 (-0.04, 0.62) | **0.19*** (0.01, 0.67) | **0.25*** (0.1, 0.7) |
| | RR-CATE | | 125.09 (94.73, 248.93) | **132.88*** (100.93, 256.6) | **143.77*** (115.55, 267.93) |
| $P$(REM $\to$ W) | % | 0.75 (0.36, 1.32) | 0.94 (0.49, 2.19) | 0.99 (0.53, 2.21) | 1.07 (0.55, 2.04) |
| | CATE | | 0.19 (-0.26, 1.06) | 0.24 (-0.21, 1.06) | 0.32 (-0.14, 1.06) |
| | RR-CATE | | 125.45 (68.89, 278.93) | 132.72 (72.47, 279.01) | 142.71 (75.83, 275.8) |
| $P$(NREM $\rightleftarrows$ REM) | % | 4.19 (2.29, 6) | 5.75 (4.26, 14.95) | 6.22 (4.57, 15.85) | 6.91 (5.18, 17.1) |
| | CATE | | **1.56*** (0.63, 10.4) | **2.03*** (1.08, 11.05) | **2.73*** (1.64, 11.75) |
| | RR-CATE | | **137.26*** (115.38, 329.3) | **148.46*** (126.77, 337.19) | **165.11*** (138.22, 348.28) |
| $P$(N1 $\rightleftarrows$ N2) | % | 6.74 (5.51, 9.8) | 8.04 (6.05, 10.06) | 9.02 (6.83, 10.91) | 10.6 (8.1, 12.59) |
| | CATE | | 1.3 (-0.93, 2.28) | 2.28 (-0.41, 3.28) | **3.86*** (0.27, 4.94) |
| | RR-CATE | | 119.35 (88.57, 136.32) | 133.89 (95.73, 153.19) | **157.27*** (102.64, 182.3) |
| $P$(Sleep compactness) | % | 90.09 (86.76, 93.12) | 88.91 (77.31, 92.11) | 87.86 (75.78, 91.49) | 86.23 (73.58, 90.35) |
| | CATE | | -1.18 (-12.26, 1.46) | -2.23 (-13.14, 0.35) | **-3.85*** (-14.94, -0.97) |
| | RR-CATE | | 98.69 (86.34, 101.66) | 97.52 (85.03, 100.39) | **95.72*** (83.55, 98.9) |
| $P$(Sleep fragmentation) | % | 7.64 (5.14, 10.41) | 9.43 (6.36, 22.33) | 10.37 (7, 23.46) | 11.83 (7.99, 25.47) |
| | CATE | | 1.79 (-0.15, 12.7) | **2.73*** (0.69, 14.03) | **4.2*** (1.85, 15.68) |
| | RR-CATE | | 123.47 (97.84, 231.34) | **135.8*** (109.56, 244.75) | **154.96*** (126.99, 267.12) |
| $P$(Sleep-stage compactness) | % | 74.39 (65.36, 80.53) | 68.28 (39.47, 74.61) | 65.7 (37.23, 71.94) | 61.74 (33.12, 68.45) |
| | CATE | | **-6.11*** (-28.22, -1.72) | **-8.7*** (-31.06, -4.67) | **-12.66*** (-34.15, -8.66) |
| | RR-CATE | | **91.78*** (57.61, 97.68) | **88.31*** (53.79, 93.69) | **82.99*** (49.69, 88.56) |
| $P$(Sleep-stage fragmentation) | % | 15.7 (12.01, 22.29) | 20.63 (16.3, 37.53) | 22.16 (17.53, 38.56) | 24.5 (19.28, 40.46) |
| | CATE | | **4.93*** (2.07, 15.87) | **6.46*** (3.51, 17.25) | **8.8*** (5.62, 18.57) |
| | RR-CATE | | **131.44*** (112.95, 178.98) | **141.18*** (122.93, 180.9) | **156.07*** (137.57, 196.56) |
| $P$(W $\to$ W) | % | 2.27 (0.56, 4.3) | 1.66 (0.2, 5.04) | 1.77 (0.21, 5.48) | 1.93 (0.21, 5.95) |
| | CATE | | -0.61 (-2.04, 2.34) | -0.5 (-1.96, 2.83) | -0.34 (-1.86, 3.53) |
| | RR-CATE | | 72.97 (27.74, 209.34) | 77.9 (28.66, 222.01) | 84.89 (30.26, 252.4) |
| $P$(N1 $\to$ N1) | % | 5.17 (2.5, 7.02) | 4.7 (1.2, 6.01) | 5.47 (1.29, 7.09) | 6.8 (1.45, 8.75) |
| | CATE | | -0.47 (-2.46, 1.46) | 0.31 (-2.03, 2.41) | 1.63 (-1.27, 4.05) |
| | RR-CATE | | 90.98 (45.72, 136.63) | 105.94 (52.65, 156.15) | 131.56 (67.16, 194.25) |
| $P$(N2 $\to$ N2) | % | 36.43 (25.04, 41.75) | 38.02 (14.13, 41.41) | 36.3 (13.4, 39.8) | 33.47 (10.67, 37.31) |
| | CATE | | 1.59 (-11.16, 4.74) | -0.13 (-12.11, 2.87) | **-2.96*** (-13.94, -0.23) |
| | RR-CATE | | 104.37 (56.45, 114.73) | 99.65 (54.01, 107.84) | **91.87*** (45.9, 99.37) |
| $P$(N3 $\to$ N3) | % | 13.74 (1.39, 19.1) | 11.52 (0.29, 19.09) | 10.3 (0.27, 17.7) | 8.61 (0.24, 15.4) |
| | CATE | | -2.23 (-8.07, 6.21) | -3.44 (-8.55, 4.26) | -5.13 (-10.17, 1.88) |
| | RR-CATE | | 83.81 (18.41, 155.79) | 74.96 (16.25, 139.74) | 62.66 (13.37, 116.7) |
| $P$(REM $\to$ REM) | % | 19.05 (15.74, 34.09) | 14.04 (9.12, 23.19) | 13.62 (8.99, 22.08) | 12.86 (8.21, 20.54) |
| | CATE | | **-5.01*** (-12.62, -0.82) | **-5.43*** (-13.65, -1.78) | **-6.19*** (-15.34, -2.91) |
| | RR-CATE | | **73.68*** (46.42, 95.58) | **71.48*** (44.74, 89.62) | **67.5*** (41.94, 83.34) |
| $P$(W-fragmentation) | % | 3.67 (2.44, 4.71) | 4.49 (3.07, 12.63) | 4.98 (3.43, 13.44) | 5.76 (3.98, 14.9) |
| | CATE | | 0.82 (-0.09, 8.2) | **1.32*** (0.43, 9.21) | **2.09*** (1.08, 10.29) |
| | RR-CATE | | 122.43 (97, 282.77) | **135.86*** (111.56, 299.2) | **156.97*** (132.91, 323.74) |
| $P$(N1-fragmentation) | % | 6.46 (5.06, 8.57) | 7.64 (6.16, 10.09) | 8.5 (6.91, 10.92) | 9.85 (8.09, 12.24) |
| | CATE | | **1.18*** (0.26, 2.35) | **2.04*** (1.07, 3.08) | **3.39*** (2.28, 4.4) |
| | RR-CATE | | **118.29*** (104.19, 140.53) | **131.5*** (117.26, 154.57) | **152.39*** (133.8, 174.95) |
| $P$(N2-fragmentation) | % | 7.46 (5.96, 11.83) | 9.76 (7.27, 16.94) | 10.43 (7.77, 17.27) | 11.45 (8.41, 17.67) |
| | CATE | | **2.3*** (0.54, 5.29) | **2.97*** (0.94, 5.77) | **4*** (1.55, 6.24) |
| | RR-CATE | | **130.91*** (107.81, 162.45) | **139.87*** (113.93, 165.49) | **153.61*** (122.92, 181.7) |
| $P$(N3-fragmentation) | % | 3.13 (2.19, 4.81) | 4.58 (3.38, 10.81) | 4.74 (3.51, 10.99) | 4.96 (3.7, 11.26) |
| | CATE | | **1.45*** (0.69, 6.26) | **1.61*** (0.86, 6.41) | **1.82*** (1.11, 6.42) |
| | RR-CATE | | **146.22*** (121.73, 238.07) | **151.37*** (128.16, 236.6) | **158.23*** (136.63, 243.25) |
| $P$(REM-fragmentation) | % | 2.62 (1.31, 3.75) | 3.59 (2.54, 10.36) | 3.88 (2.75, 10.83) | 4.32 (3.06, 11.6) |
| | CATE | | **0.97*** (0.16, 7.21) | **1.26*** (0.48, 7.82) | **1.7*** (0.87, 8.38) |
| | RR-CATE | | **137.07*** (105.93, 322.35) | **148.33*** (117.64, 343.87) | **165.09*** (133.09, 368.89) |

Table 5: Summary of expected probabilities (%) and estimated effects of OSA (CATE, RR-CATE) for 30-year-old males.

| Quantity | Estimate | Healthy | O1: OSA (AHI = 5) | O2: OSA (AHI = 15) | O3: OSA (AHI = 30) |
|---|---|---|---|---|---|
| $P$(W) | % | 11.24 (6.32, 14.97) | 10.23 (8.07, 12.87) | 10.99 (8.65, 13.52) | 12.08 (9.58, 14.94) |
| | CATE | | -1 (-4.77, 5.2) | -0.25 (-4.31, 6.31) | 0.84 (-3.04, 7.9) |
| | RR-CATE | | 91.08 (67.3, 179.25) | 97.77 (70.51, 196.92) | 107.52 (78.54, 216.37) |
| $P$(N1) | % | 13.92 (11.14, 16.57) | 14.65 (12.98, 16.52) | 16.57 (14.99, 18.22) | 19.71 (18.14, 21.23) |
| | CATE | | 0.73 (-1.52, 3.8) | **2.65*** (0.5, 5.49) | **5.79*** (3.49, 8.58) |
| | RR-CATE | | 105.21 (90.75, 134.95) | **119.02*** (103.32, 150.89) | **141.56*** (123.49, 176.82) |
| $P$(N2) | % | 42.78 (40.29, 47.62) | 46.56 (42.59, 48.67) | 45.15 (41.37, 46.85) | 42.79 (38.66, 44.68) |
| | CATE | | 3.79 (-3.26, 6.64) | 2.37 (-4.88, 4.75) | 0.01 (-7.33, 1.96) |
| | RR-CATE | | 108.85 (93.17, 116.39) | 105.55 (89.76, 111.91) | 100.03 (85.14, 104.85) |
| $P$(N3) | % | 15.41 (8.24, 19.16) | 14.72 (7.67, 17.32) | 13.61 (7.51, 15.62) | 12.06 (7.73, 13.75) |
| | CATE | | -0.69 (-6.21, 6.17) | -1.81 (-6.96, 4.71) | -3.36 (-8.05, 2.76) |
| | RR-CATE | | 95.51 (65.63, 161.66) | 88.29 (63, 147.19) | 78.23 (57.57, 126.81) |
| $P$(REM) | % | 16.65 (13.92, 23) | 13.83 (12.06, 21.33) | 13.69 (12.13, 21.48) | 13.37 (11.58, 20.8) |
| | CATE | | -2.82 (-9.2, 2.94) | -2.96 (-9.35, 2.55) | -3.29 (-9.84, 2.05) |
| | RR-CATE | | 83.08 (58.65, 116.08) | 82.2 (58.03, 113.47) | 80.26 (55.27, 112.83) |
| $P$((N1,N2,N3,REM) $\to$ W) | % | 3.89 (2.81, 4.89) | 4.83 (4.08, 7.49) | 5.25 (4.52, 8.01) | 5.9 (5.1, 8.7) |
| | CATE | | 0.93 (-0.18, 4.31) | **1.36*** (0.22, 4.7) | **2.01*** (0.73, 5.36) |
| | RR-CATE | | 123.93 (95.69, 233.47) | **134.9*** (104.54, 246.1) | **151.53*** (116.55, 269.71) |
| $P$((N1,N2) $\to$ W) | % | 2.72 (2.12, 3.28) | 3.34 (2.81, 5.1) | 3.68 (3.17, 5.59) | 4.23 (3.66, 6.27) |
| | CATE | | 0.62 (-0.12, 2.8) | **0.97*** (0.22, 3.12) | **1.51*** (0.68, 3.77) |
| | RR-CATE | | 122.82 (96.02, 214.24) | **135.67*** (107.12, 226.57) | **155.7*** (121.47, 257.59) |
| $P$(N3 $\to$ W) | % | 0.49 (0.27, 0.6) | 0.62 (0.5, 0.91) | 0.65 (0.53, 0.92) | 0.7 (0.59, 0.95) |
| | CATE | | 0.13 (-0.04, 0.56) | 0.16 (0, 0.57) | **0.21*** (0.08, 0.59) |
| | RR-CATE | | 126.26 (94.09, 285.85) | **133.21*** (100.83, 293.09) | **142.55*** (113.48, 289.69) |
| $P$(REM $\to$ W) | % | 0.69 (0.38, 1.04) | 0.87 (0.69, 1.6) | 0.92 (0.74, 1.64) | 0.98 (0.78, 1.57) |
| | CATE | | 0.18 (-0.29, 0.99) | 0.23 (-0.26, 1.05) | 0.29 (-0.25, 1.1) |
| | RR-CATE | | 126.63 (69.68, 327.29) | 133.05 (74.48, 338.65) | 141.5 (76.35, 344.74) |
| $P$(NREM $\rightleftarrows$ REM) | % | 3.57 (2.06, 4.17) | 4.92 (4.17, 7.93) | 5.29 (4.63, 8.25) | 5.82 (5.13, 9.05) |
| | CATE | | **1.36*** (0.59, 5.02) | **1.72*** (1.03, 5.45) | **2.25*** (1.39, 6.12) |
| | RR-CATE | | **138.03*** (115.19, 322.55) | **148.3*** (127.19, 333.03) | **163.19*** (135.24, 347.69) |
| $P$(N1 $\rightleftarrows$ N2) | % | 6.67 (5.58, 8.53) | 8.03 (7.22, 11.05) | 8.95 (8.15, 11.9) | 10.4 (9.44, 13.2) |
| | CATE | | 1.37 (-0.64, 3.14) | **2.28*** (0.21, 4.01) | **3.73*** (1.59, 5.37) |
| | RR-CATE | | 120.48 (92.28, 145.73) | **134.24*** (102.62, 160.17) | **155.96*** (119.33, 184.75) |
| $P$(Sleep compactness) | % | 85.13 (81.26, 90.66) | 85.49 (81.46, 87.39) | 84.31 (80.57, 86.34) | 82.55 (79.16, 84.74) |
| | CATE | | 0.36 (-6.99, 4.58) | -0.83 (-8.26, 3.63) | -2.59 (-10.21, 2.07) |
| | RR-CATE | | 100.42 (92.38, 105.67) | 99.03 (90.93, 104.45) | 96.96 (88.7, 102.55) |
| $P$(Sleep fragmentation) | % | 7.52 (5.6, 9.16) | 9.1 (7.75, 14.61) | 9.96 (8.62, 15.84) | 11.27 (9.77, 17.24) |
| | CATE | | 1.58 (-0.46, 8.5) | **2.44*** (0.41, 9.44) | **3.75*** (1.43, 10.52) |
| | RR-CATE | | 120.95 (94.71, 231.03) | **132.38*** (104.62, 244.07) | **149.83*** (116.84, 268.84) |
| $P$(Sleep-stage compactness) | % | 70.53 (66.74, 76.39) | 66.25 (53.63, 69.03) | 63.77 (51.35, 66.39) | 60.08 (47.63, 62.8) |
| | CATE | | -4.28 (-18.11, 0.4) | **-6.75*** (-19.95, -2.64) | **-10.45*** (-22.81, -6.6) |
| | RR-CATE | | 93.94 (75.3, 100.6) | **90.42*** (71.73, 96.17) | **85.19*** (67.67, 90.29) |
| $P$(Sleep-stage fragmentation) | % | 14.6 (12.21, 16.73) | 19.24 (17.38, 28.47) | 20.53 (18.59, 29.25) | 22.46 (20.37, 30.79) |
| | CATE | | **4.63*** (1.93, 12.96) | **5.93*** (3.34, 14) | **7.86*** (5.18, 15.38) |
| | RR-CATE | | **131.74*** (112.14, 182.36) | **140.58*** (120.97, 188.27) | **153.82*** (132.46, 200.52) |
| $P$(W $\to$ W) | % | 7.34 (3.15, 10.9) | 5.41 (1.99, 7.71) | 5.73 (2.1, 8.4) | 6.18 (2.33, 9.09) |
| | CATE | | -1.93 (-5.73, 3.84) | -1.61 (-5.5, 4.5) | -1.16 (-5.04, 5.34) |
| | RR-CATE | | 73.66 (36.6, 213.54) | 78.09 (38.66, 223.98) | 84.17 (41.58, 239.56) |
| $P$(N1 $\to$ N1) | % | 7.46 (5.36, 9.7) | 6.85 (4.4, 7.99) | 7.92 (5.15, 8.92) | 9.73 (5.87, 10.89) |
| | CATE | | -0.61 (-3.63, 2) | 0.46 (-2.42, 2.93) | 2.27 (-0.9, 4.96) |
| | RR-CATE | | 91.83 (60.77, 136.2) | 106.21 (71.69, 153.63) | 130.45 (87.87, 191.07) |
| $P$(N2 $\to$ N2) | % | 36.19 (33.54, 40.89) | 38.13 (29.83, 39.94) | 36.16 (27.67, 37.6) | 32.97 (24.66, 34.48) |
| | CATE | | 1.94 (-7.08, 4.81) | -0.04 (-8.8, 2.31) | **-3.22*** (-12.03, -1) |
| | RR-CATE | | 105.35 (80.47, 114.4) | 99.9 (76.64, 106.67) | **91.09*** (68.65, 97.12) |
| $P$(N3 $\to$ N3) | % | 12.58 (5.16, 16.85) | 10.64 (1.77, 13.24) | 9.46 (1.68, 11.51) | 7.82 (1.25, 9.85) |
| | CATE | | -1.94 (-8.96, 4.8) | -3.13 (-9.45, 3.27) | -4.77 (-10.57, 1.2) |
| | RR-CATE | | 84.59 (26.11, 160.79) | 75.15 (23.56, 142.39) | 62.13 (20, 116.59) |
| $P$(REM $\to$ REM) | % | 14.29 (12.02, 20.89) | 10.63 (9.07, 15.82) | 10.24 (8.83, 16.01) | 9.57 (7.98, 14.74) |
| | CATE | | -3.66 (-10.28, 0.27) | **-4.05*** (-10.69, -0.47) | **-4.73*** (-11.41, -1.35) |
| | RR-CATE | | 74.37 (47.68, 102.02) | **71.66*** (45.58, 97.18) | **66.93*** (42.1, 91.3) |
| $P$(W-fragmentation) | % | 3.63 (2.76, 4.24) | 4.27 (3.6, 7.24) | 4.71 (4.01, 7.9) | 5.37 (4.65, 8.81) |
| | CATE | | 0.64 (-0.31, 3.97) | **1.08*** (0.12, 4.68) | **1.74*** (0.71, 5.23) |
| | RR-CATE | | 117.75 (91.99, 223.8) | **129.68*** (102.91, 242.99) | **148.01*** (118.69, 265.48) |
| $P$(N1-fragmentation) | % | 6.68 (5.68, 7.58) | 7.92 (7.11, 10.71) | 8.75 (7.98, 11.37) | 10.05 (9.2, 12.87) |
| | CATE | | **1.24*** (0.19, 3.22) | **2.08*** (1.1, 3.96) | **3.37*** (2.28, 5.24) |
| | RR-CATE | | **118.65*** (102.67, 150.56) | **131.12*** (114.5, 163.56) | **150.49*** (132.49, 183.02) |
| $P$(N2-fragmentation) | % | 6.81 (5.74, 8.11) | 8.98 (8.04, 13.65) | 9.54 (8.61, 14.16) | 10.39 (9.41, 14.75) |
| | CATE | | **2.17*** (0.54, 6.23) | **2.74*** (0.96, 6.73) | **3.58*** (1.51, 7.4) |
| | RR-CATE | | **131.94*** (106.68, 180.12) | **140.2*** (112.16, 187.98) | **152.65*** (119.04, 199.3) |
| $P$(N3-fragmentation) | % | 2.8 (2.07, 3.14) | 4.1 (3.53, 6.75) | 4.19 (3.65, 6.75) | 4.31 (3.79, 6.97) |
| | CATE | | **1.3*** (0.66, 4.02) | **1.4*** (0.74, 4.15) | **1.51*** (0.85, 4.05) |
| | RR-CATE | | **146.53*** (122.09, 247.27) | **149.99*** (126, 245.88) | **154*** (129.54, 235.46) |
| $P$(REM-fragmentation) | % | 2.22 (1.12, 2.71) | 3.07 (2.62, 5.16) | 3.29 (2.82, 5.3) | 3.62 (3.13, 5.72) |
| | CATE | | **0.85*** (0.17, 3.46) | **1.07*** (0.4, 3.63) | **1.4*** (0.72, 4.01) |
| | RR-CATE | | **138.16*** (107.36, 329.54) | **148.39*** (116.67, 340.06) | **163.17*** (128.56, 342.75) |

Table 6: Summary of expected probabilities (%) and estimated effects of OSA (CATE, RR-CATE) for 50-year-old males.

| Quantity | Estimate | Healthy | O1: OSA (AHI = 5) | O2: OSA (AHI = 15) | O3: OSA (AHI = 30) |
|---|---|---|---|---|---|
| $P$(W) | % | 24.25 (9.24, 31.04) | 20.28 (14.57, 23.61) | 21.39 (15.68, 24.18) | 22.89 (17.57, 25.22) |
| | CATE | | -3.98 (-12.28, 7.19) | -2.87 (-10.6, 8.48) | -1.37 (-8.93, 10.23) |
| | RR-CATE | | 83.61 (58.88, 179.93) | 88.17 (64.35, 190.48) | 94.36 (70.98, 208.42) |
| $P$(N1) | % | 15.26 (12.59, 17.75) | 16.17 (14.35, 19.68) | 18.14 (16.67, 21.86) | 21.3 (19.9, 25.12) |
| | CATE | | 0.91 (-2.07, 4.87) | 2.88 (0, 7.05) | **6.04*** (3.33, 10.05) |
| | RR-CATE | | 105.97 (88.64, 135.5) | **118.86*** (100.01, 148.98) | **139.58*** (118.92, 173.61) |
| $P$(N2) | % | 36.95 (34.21, 45.05) | 41.6 (38.29, 45.66) | 39.8 (37.04, 43.64) | 36.96 (34.69, 40.61) |
| | CATE | | 4.65 (-2.41, 10.6) | 2.85 (-3.78, 8.58) | 0.01 (-6.45, 4.89) |
| | RR-CATE | | 112.58 (94.63, 130.46) | 107.71 (91.72, 124.14) | 100.03 (85.57, 114.11) |
| $P$(N3) | % | 12.32 (6.17, 19.21) | 12.15 (8.56, 15.05) | 11.07 (7.99, 13.4) | 9.6 (7.31, 11.37) |
| | CATE | | -0.16 (-9.43, 4.58) | -1.24 (-9.87, 3.39) | -2.72 (-10.59, 1.97) |
| | RR-CATE | | 98.69 (49.31, 162.84) | 89.92 (46.76, 147.29) | 77.91 (42.11, 130) |
| $P$(REM) | % | 11.22 (6.3, 21.78) | 9.8 (8.31, 13.78) | 9.61 (8.34, 13.08) | 9.26 (8.03, 12.03) |
| | CATE | | -1.42 (-8.22, 3.48) | -1.61 (-8.95, 3.42) | -1.96 (-10.27, 3.21) |
| | RR-CATE | | 87.32 (58.31, 155.78) | 85.61 (57.35, 154.62) | 82.5 (53.48, 150.53) |
| $P$((N1,N2,N3,REM) $\rightarrow$ W) | % | 3.46 (1.76, 5.31) | 4.39 (3.53, 5.99) | 4.75 (3.96, 6.23) | 5.28 (4.5, 6.55) |
| | CATE | | 0.93 (-0.1, 3.15) | **1.28*** (0.3, 3.38) | **1.81*** (0.92, 3.83) |
| | RR-CATE | | 126.78 (97.65, 274.51) | **137.01*** (107.01, 287.19) | **152.28*** (118.33, 318.1) |
| $P$((N1,N2) $\rightarrow$ W) | % | 2.54 (1.44, 3.7) | 3.18 (2.54, 4.22) | 3.49 (2.92, 4.45) | 3.96 (3.46, 4.87) |
| | CATE | | **0.64*** (0.01, 1.93) | **0.95*** (0.32, 2.27) | **1.42*** (0.8, 2.81) |
| | RR-CATE | | **125.16*** (100.25, 234.19) | **137.22*** (109.17, 259.82) | **155.72*** (122.68, 293.63) |
| $P$(N3 $\rightarrow$ W) | % | 0.36 (0.14, 0.46) | 0.48 (0.37, 0.59) | 0.5 (0.39, 0.6) | 0.52 (0.42, 0.62) |
| | CATE | | 0.11 (-0.01, 0.37) | **0.13*** (0.02, 0.41) | **0.16*** (0.06, 0.42) |
| | RR-CATE | | 130.99 (97.6, 352.35) | **136.52*** (105.43, 363.08) | **143.46*** (112.55, 385.68) |
| $P$(REM $\rightarrow$ W) | % | 0.56 (0.17, 1.17) | 0.74 (0.55, 1.17) | 0.76 (0.58, 1.16) | 0.8 (0.61, 1.14) |
| | CATE | | 0.18 (-0.24, 0.55) | 0.2 (-0.23, 0.57) | 0.24 (-0.23, 0.6) |
| | RR-CATE | | 131.38 (77.78, 452.58) | 136.36 (78.9, 473.86) | 142.4 (80.22, 447.72) |
| $P$(NREM $\rightleftarrows$ REM) | % | 2.72 (1.42, 3.48) | 3.88 (3.03, 4.74) | 4.12 (3.35, 4.9) | 4.45 (3.82, 5.22) |
| | CATE | | **1.16*** (0.55, 2.75) | **1.4*** (0.82, 2.86) | **1.73*** (1.09, 3.12) |
| | RR-CATE | | **142.56*** (117.62, 277.57) | **151.35*** (125.04, 290.05) | **163.59*** (133.12, 320.97) |
| $P$(N1 $\rightleftarrows$ N2) | % | 5.78 (4.38, 9.42) | 7.23 (6.49, 9.17) | 7.96 (7.23, 9.96) | 9.08 (8.4, 11.09) |
| | CATE | | 1.45 (-0.8, 3.93) | 2.17 (-0.02, 4.56) | **3.3*** (1.34, 5.53) |
| | RR-CATE | | 125.01 (91.53, 189.09) | 137.6 (99.81, 201.05) | **156.98*** (113.74, 223.75) |
| $P$(Sleep compactness) | % | 72.49 (64.61, 87.34) | 75.9 (71.91, 81.17) | 74.46 (71.08, 79.54) | 72.46 (69.58, 77.56) |
| | CATE | | 3.41 (-7.93, 12.57) | 1.97 (-9.56, 10.49) | -0.03 (-11.87, 8.2) |
| | RR-CATE | | 104.7 (90.85, 118.97) | 102.72 (88.99, 115.82) | 99.96 (86.31, 112.71) |
| $P$(Sleep fragmentation) | % | 6.72 (3.57, 10.33) | 8.21 (6.6, 10.69) | 8.9 (7.43, 11.21) | 9.92 (8.49, 12.25) |
| | CATE | | 1.49 (-0.42, 5.25) | **2.18*** (0.37, 5.76) | **3.21*** (1.34, 6.81) |
| | RR-CATE | | 122.22 (95.17, 250.28) | **132.44*** (104.15, 261.75) | **147.7*** (113.64, 289.47) |
| $P$(Sleep-stage compactness) | % | 60.46 (53.83, 71.54) | 59.56 (55, 64.01) | 57.23 (53.36, 61.45) | 53.93 (50.72, 58.13) |
| | CATE | | -0.9 (-11.58, 5.88) | -3.23 (-13.72, 2.9) | -6.53 (-16.86, 0.31) |
| | RR-CATE | | 98.51 (83.6, 110.46) | 94.65 (80.89, 105.56) | 89.2 (75.93, 100.6) |
| $P$(Sleep-stage fragmentation) | % | 12.03 (8.14, 17.09) | 16.34 (14.29, 19.79) | 17.24 (15.23, 20.49) | 18.53 (17.02, 21.52) |
| | CATE | | **4.31*** (1.79, 10.1) | **5.2*** (2.77, 10.34) | **6.5*** (3.98, 10.88) |
| | RR-CATE | | **135.82*** (111.39, 224.05) | **143.25*** (116.4, 226.95) | **154.04*** (124.69, 239.48) |
| $P$(W $\rightarrow$ W) | % | 20.79 (5.55, 26.66) | 15.89 (10.1, 18.62) | 16.64 (10.91, 19.14) | 17.61 (12.07, 19.78) |
| | CATE | | -4.9 (-13.62, 5.95) | -4.15 (-12.72, 6.81) | -3.18 (-10.73, 7.89) |
| | RR-CATE | | 76.42 (50.56, 209.13) | 80.03 (53.06, 222.71) | 84.71 (57.23, 234.23) |
| $P$(N1 $\rightarrow$ N1) | % | 9.43 (6.19, 11.12) | 8.99 (7.34, 11.23) | 10.27 (8.68, 12.48) | 12.39 (10.81, 15.18) |
| | CATE | | -0.45 (-3.15, 2.45) | 0.84 (-1.6, 3.6) | **2.95*** (0.48, 5.59) |
| | RR-CATE | | 95.27 (72.01, 136.91) | 108.85 (86.65, 154.44) | **131.28*** (104.19, 189.99) |
| $P$(N2 $\rightarrow$ N2) | % | 31.52 (28.89, 37.56) | 34.45 (30.83, 38.17) | 32.27 (29.1, 35.36) | 28.89 (26.32, 31.94) |
| | CATE | | 2.93 (-3.34, 7.11) | 0.75 (-4.82, 4.42) | -2.63 (-8.14, 0.51) |
| | RR-CATE | | 109.3 (90.85, 123.11) | 102.38 (86.68, 113.78) | 91.67 (78.04, 101.58) |
| $P$(N3 $\rightarrow$ N3) | % | 10.1 (4.16, 17.88) | 8.86 (5.38, 12.2) | 7.78 (4.87, 10.51) | 6.32 (4.1, 8.32) |
| | CATE | | -1.24 (-11.52, 3.46) | -2.32 (-11.89, 2.35) | -3.79 (-12.42, 0.84) |
| | RR-CATE | | 87.76 (36.17, 171.39) | 77.02 (32.56, 146.38) | 62.53 (28.65, 119.37) |
| $P$(REM $\rightarrow$ REM) | % | 9.4 (5.48, 20.11) | 7.26 (6.07, 10.79) | 6.91 (5.9, 10.11) | 6.33 (5.39, 9.11) |
| | CATE | | -2.15 (-9.37, 1.95) | -2.5 (-10.57, 1.67) | -3.07 (-12.08, 1.21) |
| | RR-CATE | | 77.16 (47.39, 136.07) | 73.44 (45.74, 131) | 67.35 (41.12, 121.62) |
| $P$(W-fragmentation) | % | 3.25 (1.81, 5.02) | 3.82 (3.07, 4.97) | 4.15 (3.42, 5.2) | 4.65 (3.99, 5.67) |
| | CATE | | 0.57 (-0.43, 2.19) | 0.9 (-0.08, 2.5) | **1.39*** (0.37, 2.97) |
| | RR-CATE | | 117.37 (90.6, 219.1) | 127.59 (98.35, 238.16) | **142.83*** (107.28, 262.14) |
| $P$(N1-fragmentation) | % | 6.12 (4.42, 8.02) | 7.49 (6.58, 9.06) | 8.19 (7.34, 9.74) | 9.24 (8.57, 10.94) |
| | CATE | | **1.37*** (0.05, 4.43) | **2.07*** (1.01, 5.04) | **3.12*** (2.08, 6) |
| | RR-CATE | | **122.43*** (100.68, 199.22) | **133.8*** (112.74, 214.14) | **151.02*** (126.83, 234.56) |
| $P$(N2-fragmentation) | % | 5.49 (3.49, 8.55) | 7.5 (6.53, 9.65) | 7.88 (7, 9.91) | 8.45 (7.7, 10.22) |
| | CATE | | **2.01*** (0.36, 4.42) | **2.4*** (0.75, 4.7) | **2.96*** (1.31, 5.11) |
| | RR-CATE | | **136.67*** (104.35, 227.21) | **143.69*** (108.64, 234.71) | **154.01*** (115.91, 246.5) |
| $P$(N3-fragmentation) | % | 2.21 (1.18, 3.3) | 3.33 (2.77, 4.2) | 3.35 (2.84, 4.09) | 3.36 (2.86, 4) |
| | CATE | | **1.13*** (0.39, 2.38) | **1.15*** (0.43, 2.28) | **1.15*** (0.46, 2.09) |
| | RR-CATE | | **151.09*** (110.91, 297.74) | **152.13*** (115.04, 289.05) | **152.37*** (115.08, 273.77) |
| $P$(REM-fragmentation) | % | 1.68 (0.8, 2.41) | 2.41 (1.96, 3.12) | 2.55 (2.07, 3.2) | 2.76 (2.33, 3.3) |
| | CATE | | **0.73*** (0.23, 1.79) | **0.87*** (0.36, 1.86) | **1.07*** (0.53, 1.97) |
| | RR-CATE | | **143.11*** (110.63, 313.22) | **151.73*** (116.31, 328.02) | **163.67*** (123.05, 338.27) |

Table 7: Summary of expected probabilities (%) and estimated effects of OSA (CATE, RR-CATE) for 70-year-old males.

**Comparison based on derived Markovian matrices $\mathbf{P}^M$**

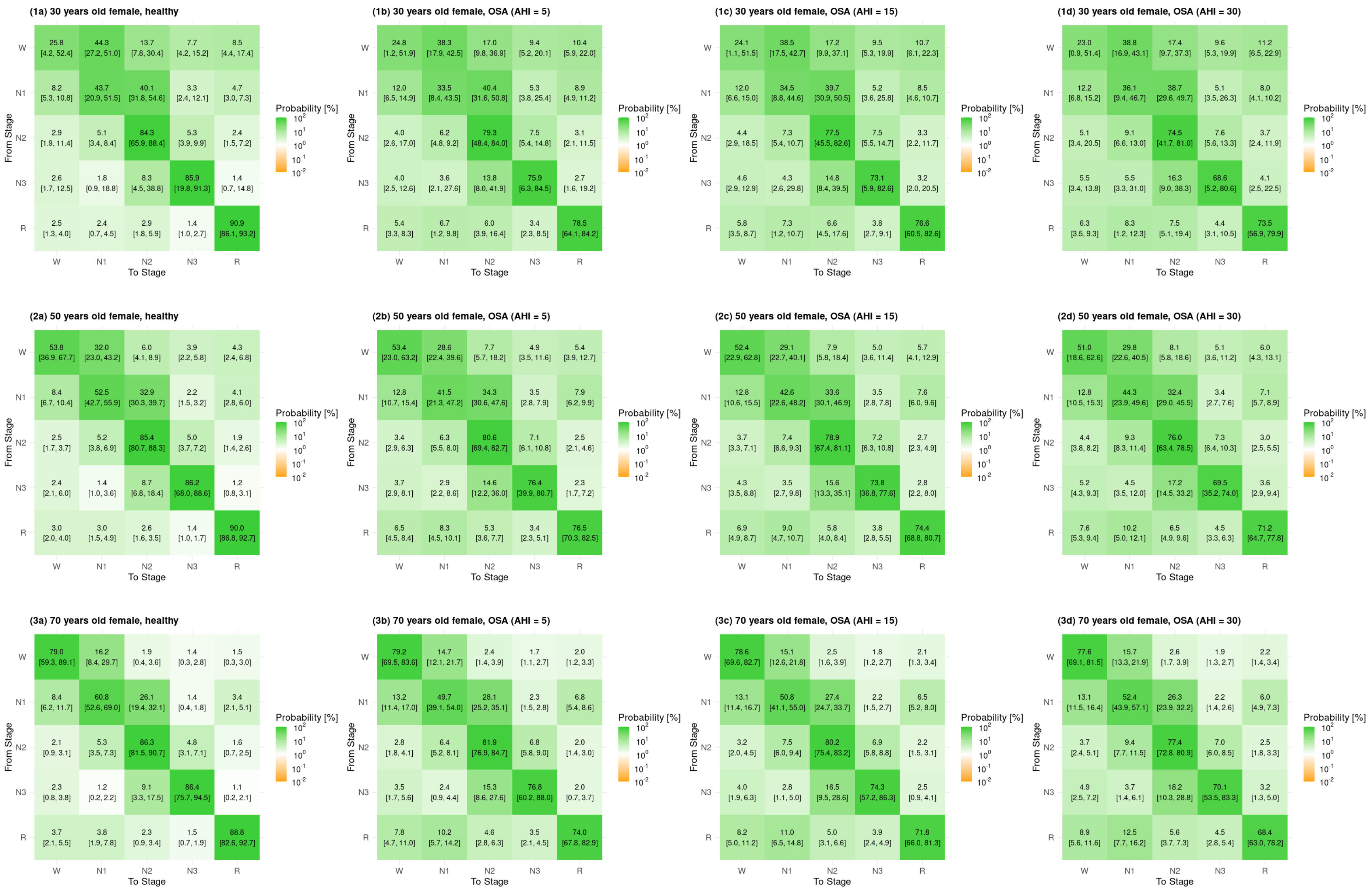


Figure 7: Derived Markovian transition matrices $\mathbf{P}^M$ for healthy females and females with OSA (AHI = 5, 15, 30) at different ages (30, 50, 70 years). Estimates are supplemented with 95% bootstrapped confidence intervals.

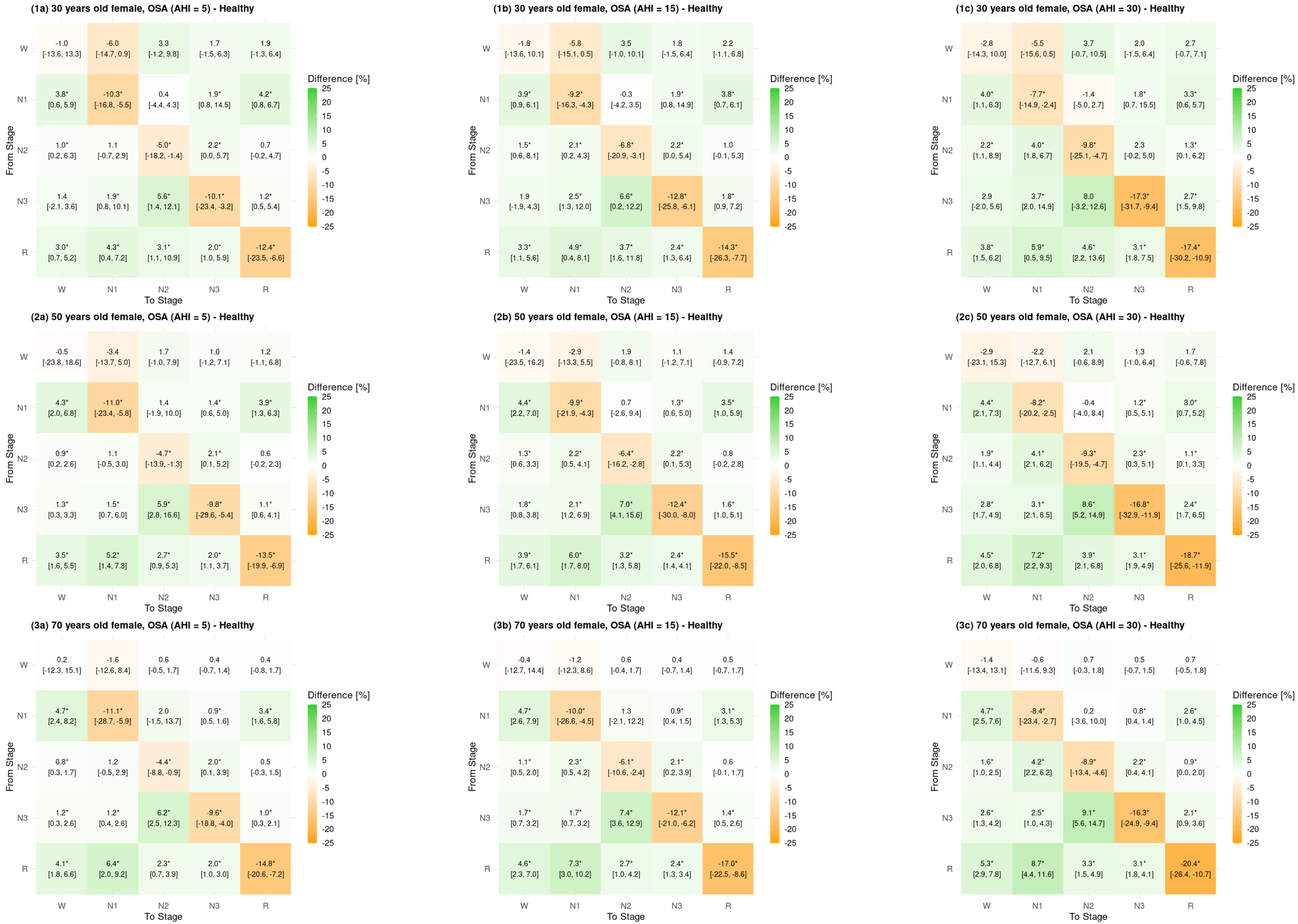


Figure 8: Difference in derived Markovian transition matrices $\mathbf{P}^M$ for healthy females versus females with OSA (AHI = 5, 15, 30) at different ages (30, 50, 70 years). Estimates are supplemented with 95% bootstrapped confidence intervals.

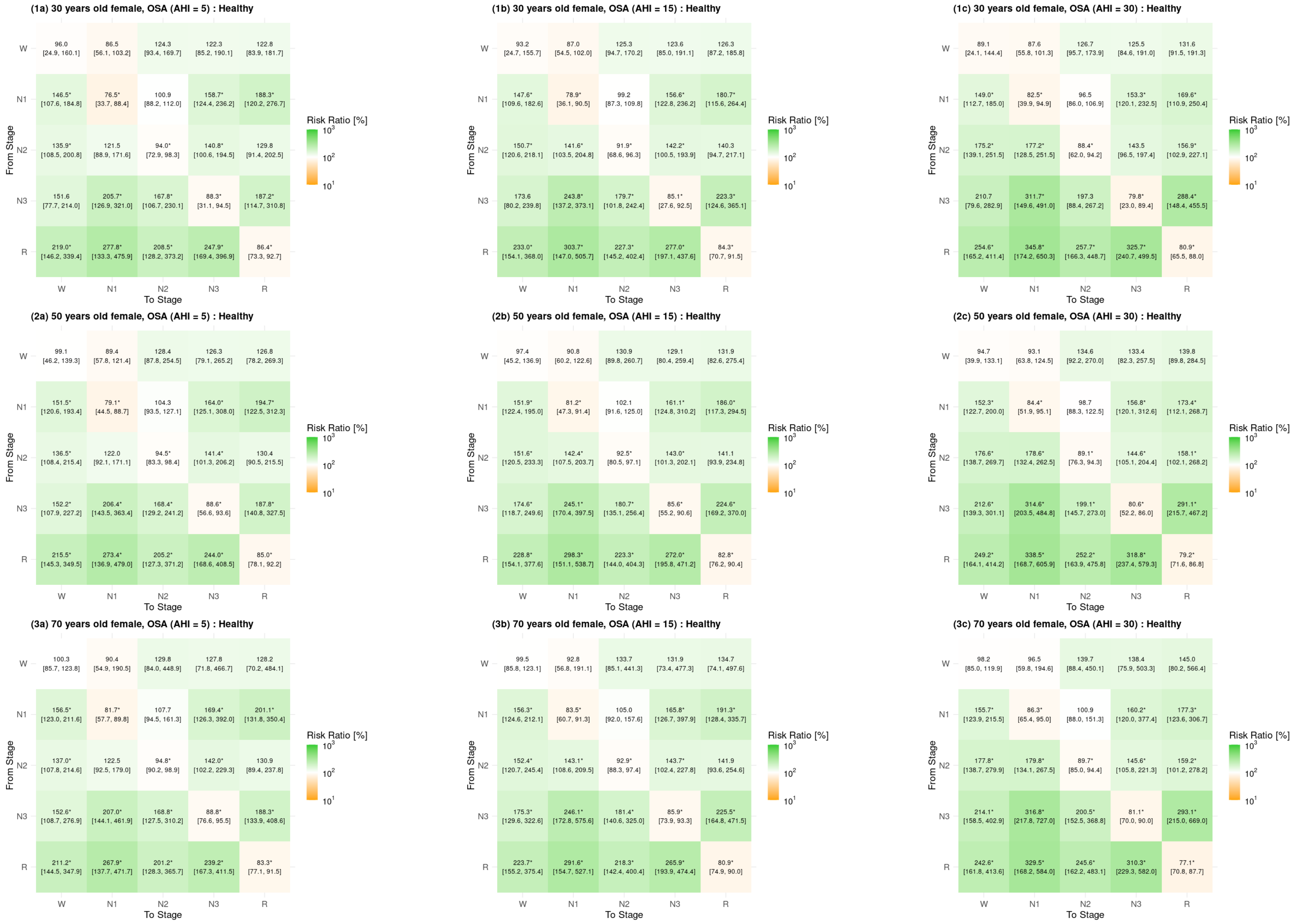


Figure 9: Risk ratio of derived Markovian transition matrices $\mathbf{P}^M$ for healthy females versus females with OSA (AHI = 5, 15, 30) at different ages (30, 50, 70 years). Estimates are supplemented with 95% bootstrapped confidence intervals.

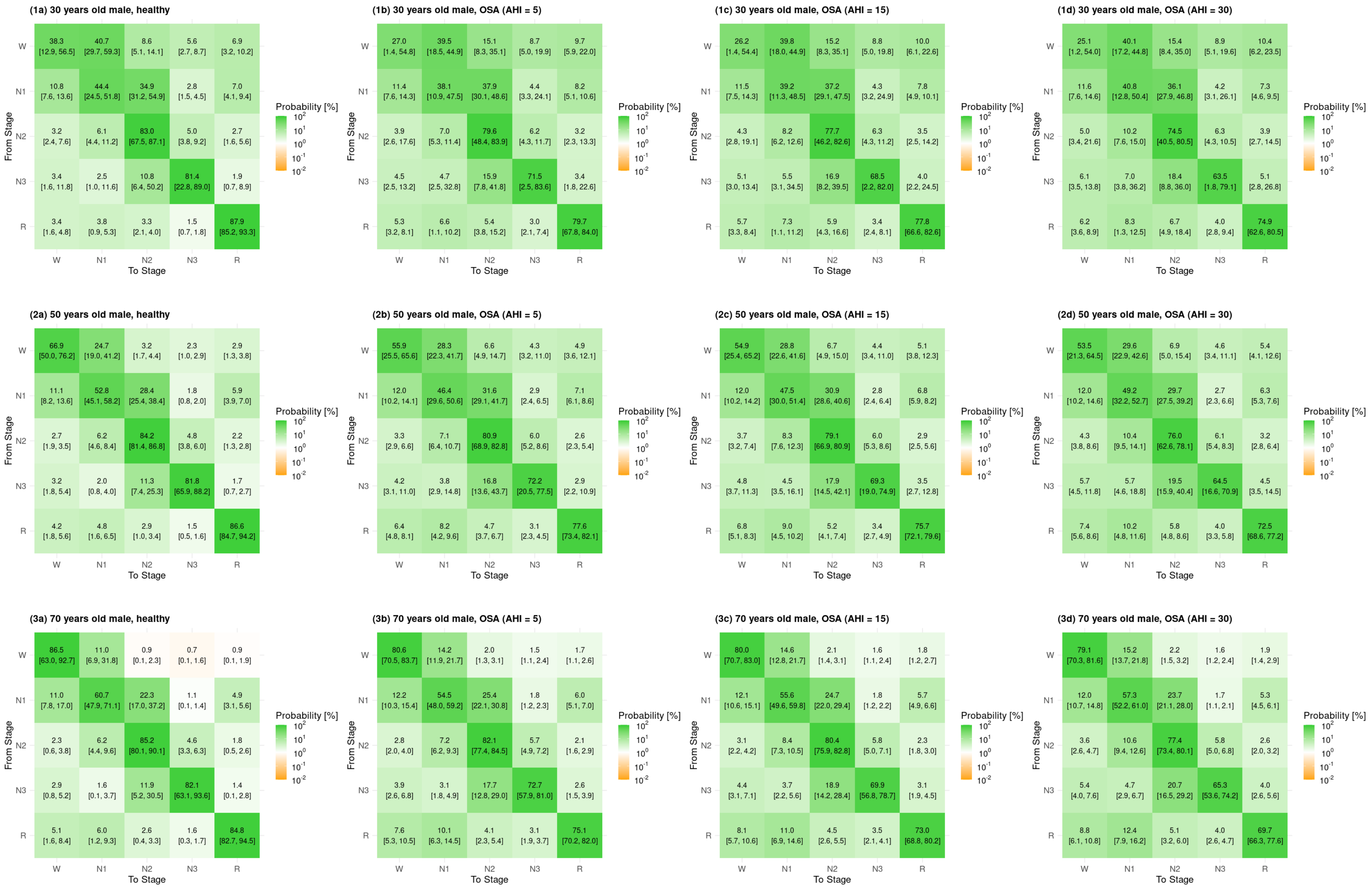


Figure 10: Derived Markovian transition matrices $\mathbf{P}^M$ for healthy males and males with OSA (AHI = 5, 15, 30) at different ages (30, 50, 70 years). Estimates are supplemented with 95% bootstrapped confidence intervals.

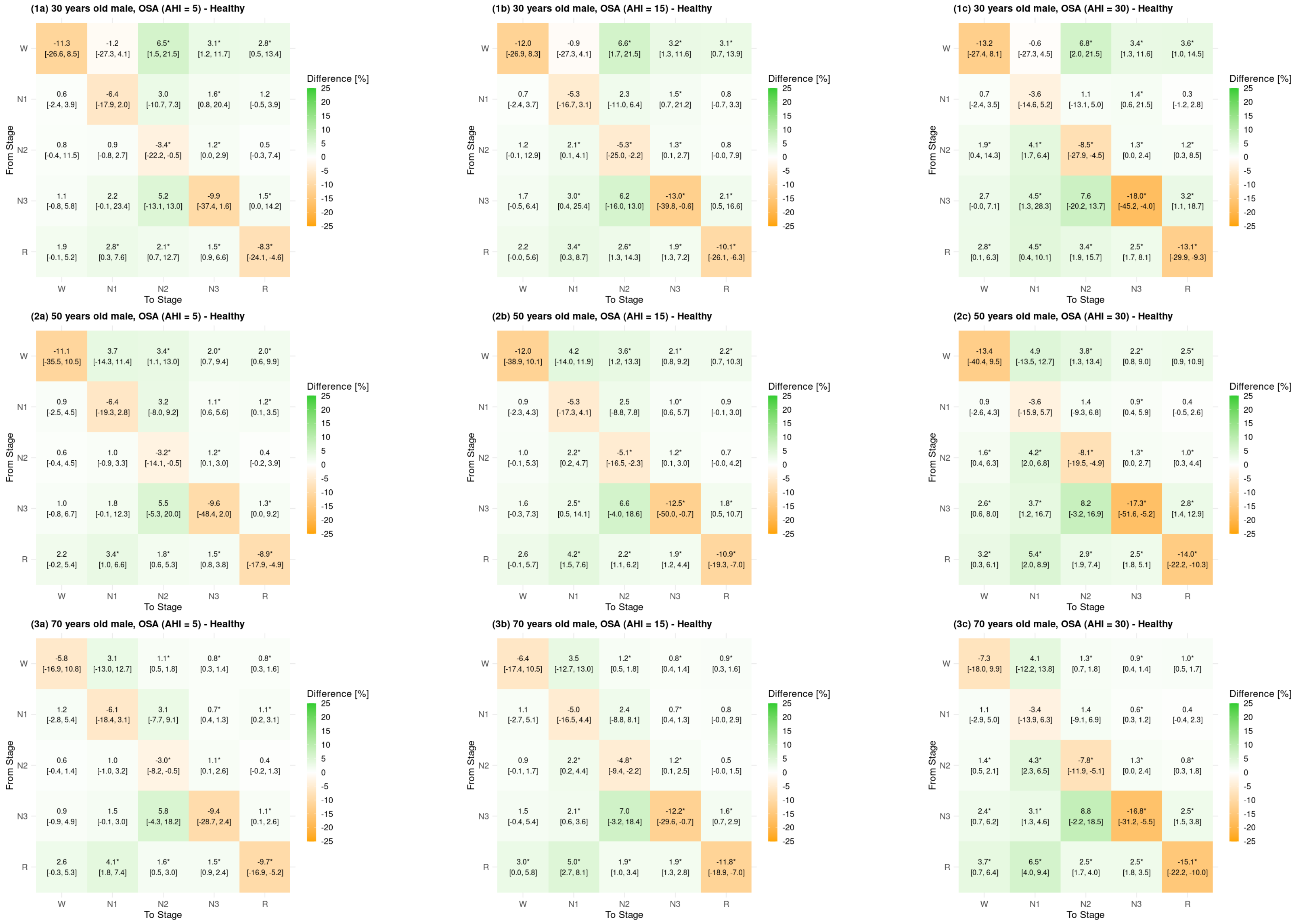


Figure 11: Difference in derived Markovian transition matrices $\mathbf{P}^M$ for healthy males versus males with OSA (AHI = 5, 15, 30) at different ages (30, 50, 70 years). Estimates are supplemented with 95% bootstrapped confidence intervals.

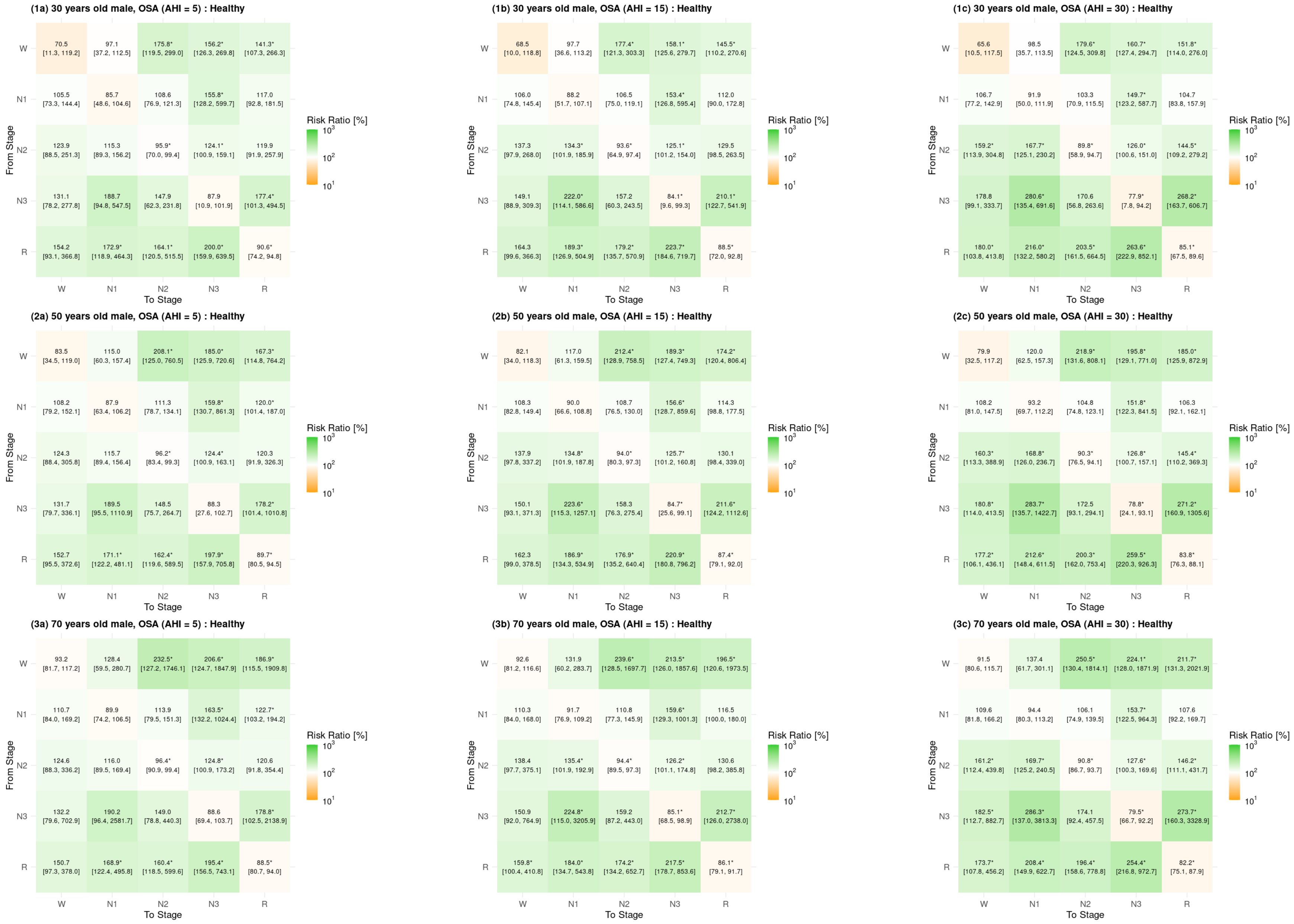


Figure 12: Risk ratio of derived Markovian transition matrices $\mathbf{P}^M$ for healthy males versus males with OSA (AHI = 5, 15, 30) at different ages (30, 50, 70 years). Estimates are supplemented with 95% bootstrapped confidence intervals.

### Effect plots

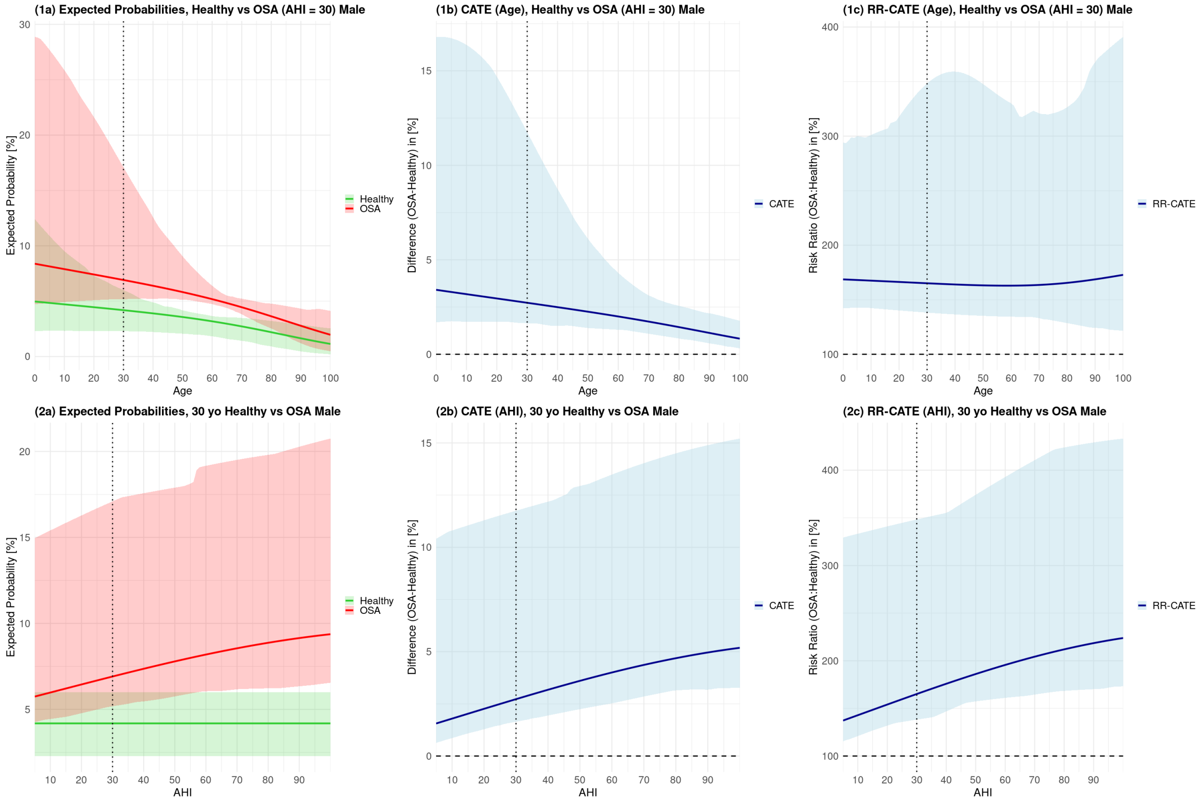


Figure 13: Effects of age and OSA-severities on NREM-REM oscillations, $P(\text{NREM} \rightleftarrows \text{REM})$, in males. The left plots (1a, 2a) depict expected probabilities for varying age with fixed AHI = 30, and for varying AHI with fixed age = 30. Based on that, the central (1b, 2b) and right (1c, 2c) plots depict age- and AHI-related CATE and RR-CATE.

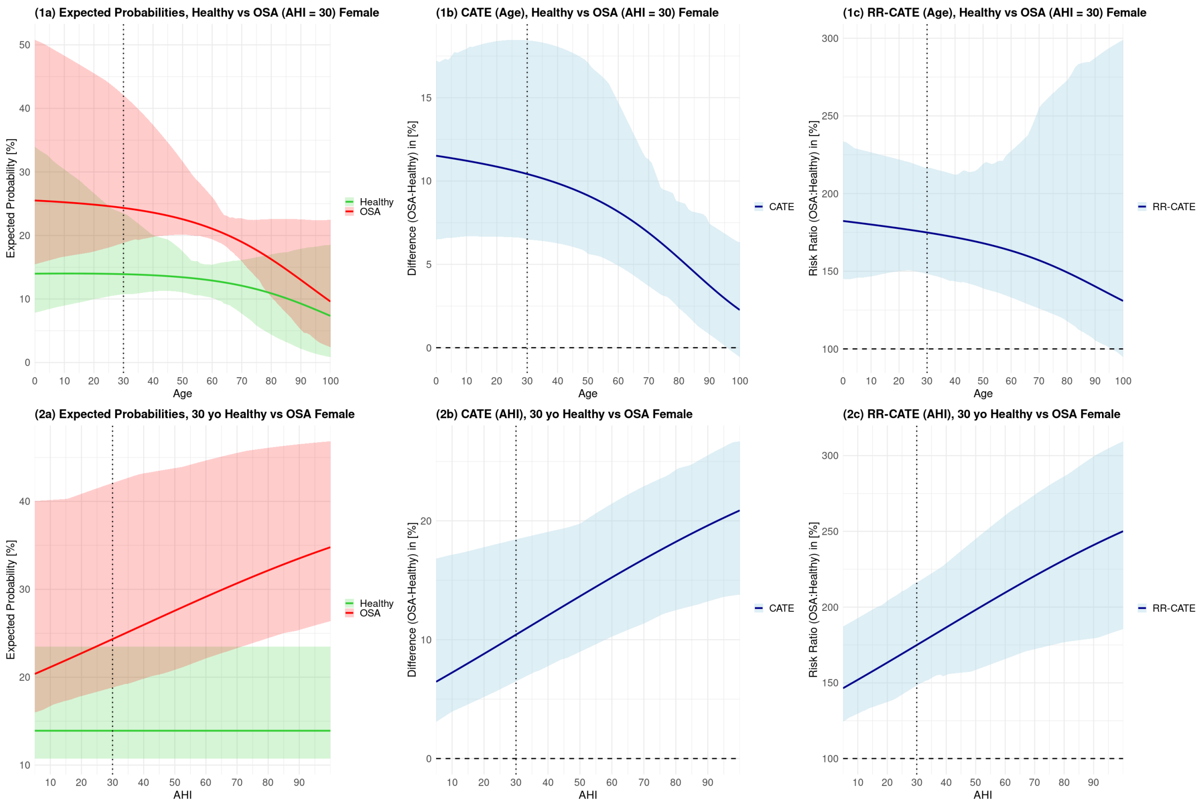


Figure 14: Effects of age and OSA-severities on sleep-stage fragmentation, i.e., the probability of transitioning from one (non-wake) sleep stage to a different one, in females. The left plots (1a, 2a) depict expected probabilities for varying age with fixed AHI = 30, and for varying AHI with fixed age = 30. Based on that, the central (1b, 2b) and right (1c, 2c) plots depict age- and AHI-related CATE and RR-CATE.

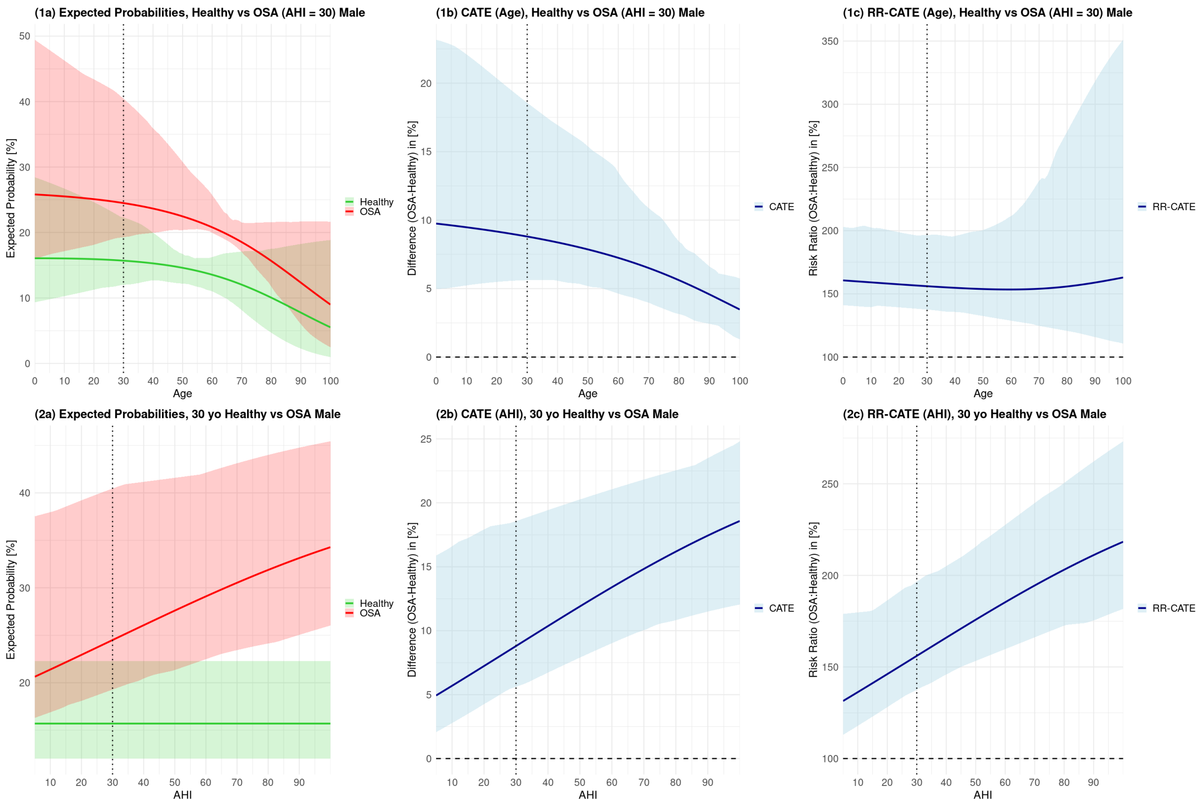


Figure 15: Effects of age and OSA-severities on sleep-stage fragmentation, i.e., the probability of transitioning from one (non-wake) sleep stage to a different one, in males. The left plots (1a, 2a) depict expected probabilities for varying age with fixed AHI = 30, and for varying AHI with fixed age = 30. Based on that, the central (1b, 2b) and right (1c, 2c) plots depict age- and AHI-related CATE and RR-CATE.
